# El Niño Southern Oscillation shapes cholera in Africa

**DOI:** 10.64898/2026.11.11.26367838

**Authors:** Javier Perez-Saez, Elizabeth C. Lee, Qulu Zheng, Raoul Kamadjeu, Benjamin Zaitchik, Andrew S. Azman

## Abstract

Cholera is considered a paradigm for climate-sensitive diseases, yet its relation with El Niño Southern Oscillation (ENSO), the dominant mode of interannual climate variability, remains poorly quantified in Africa where most cholera deaths occur. Using 15 years of surveillance data across 30 African countries and novel statistical and simulation frameworks, we identify five geographically coherent ENSO–cholera teleconnection groups, each characterized by distinct directions, strengths and timing of effects on outbreak occurrence, duration and incidence. For the unfolding major El Niño of 2026–27, we project two- to five-fold greater cholera burden in Eastern and Horn of Africa than under ENSO-neutral conditions. These findings identify immediate priorities for anticipatory action and establish a dynamic framework for attributing burden for outbreak-prone diseases to climate variability.

## Main text

Intensifying interannual climate variability linked to anthropogenic global warming poses major challenges for societal wellbeing and public health (*1–3*). At the center of this variability is the El Niño Southern Oscillation (ENSO), the dominant mode of natural interannual climate fluctuations on Earth, whose teleconnections affect temperature, precipitation, and extreme weather events across the globe (*4*, *5*). Years where equatorial Pacific Ocean temperatures are anomalously high or low, respectively defined as El Niño and La Niña events, have been consistently associated with severe social impacts, including disruption of food systems, rise in armed conflict, and the collapse of water, sanitation and hygiene (WASH) infrastructure (*6–9*). When compounded, these impacts can pose major public health challenges, particularly through the rise of climate-sensitive infectious disease outbreaks (*10*, *11*).

Cholera exemplifies this challenge. The ongoing seventh cholera pandemic kills far more than the roughly 7,000 deaths reported to the World Health Organization (WHO) in 2025, with the great majority of deaths in Africa (*12*). Yet Africa is also where ENSO effects on cholera remain most poorly understood. Despite its reputation as a model climate-sensitive disease, frequently invoked in the public health and climate change literature, the quantitative basis for cholera’s climate sensitivity is thin: rigorous evidence beyond the seminal studies linking cholera dynamics to ENSO in South Asia is scarce (*13–15*), and the evidence available for Africa does not provide testable predictions on how the timing and magnitude of a specific El Niño event affect cholera burden (*16*). In fact, studies of individual outbreaks have hinted that the global resurgence of cholera since 2022 (*17*) is linked to the 2020–23 La Niña event (*18*, *19*) without evaluating alternative hypotheses and considering the complexities of climate attribution.

The teleconnections between ENSO and weather differ sharply in direction, timing, and magnitude across the African continent (*20*), and because both floods and droughts can increase cholera outbreak risk (*11*, *21*), ENSO effects on cholera are likely region-specific. ENSO’s influence may also depend on the outbreak phase: the climatic conditions that allow outbreaks to ignite are not necessarily the same as those that sustain transmission or determine its final size.

Previous insights about this phenomenon were forestalled by limited availability of high-resolution cholera surveillance data at continental scale and the lack of analytical frameworks providing information on when, how and where ENSO matters most for igniting and fueling cholera outbreaks. Public health planning has thus been contained to qualitative intuitions to guide anticipatory action without a quantitative evidence base of ENSO-driven cholera risk. Closing this evidence gap is particularly urgent against the backdrop of the major El Niño predicted to reach record-breaking levels in 2026–27 (*22*), and governments and international agencies that are actively seeking preemptive approaches to limit cholera risk (*23*).

Here we leverage a unique 15-year surveillance database spanning 30 African countries to unravel the ENSO–cholera relationship using a Bayesian hierarchical framework that jointly estimates the direction, strength, and timing of ENSO effects on outbreak initiation, duration, and incidence at subnational scale, pooling information across the continent. We identify geographically coherent ENSO–cholera teleconnection groups, and assess the role of past major ENSO events on cholera burden in different regions of Africa. We finally turn these inferences into testable, region-specific forecasts for the unfolding 2026–27 El Niño event, identifying where anticipatory action may have the highest impact.

Our raw dataset consisted of 2,442 unique cholera outbreaks of at least 50 suspected cholera cases at subnational level, extracted following established methods from weekly and sub-weekly incidence time series spanning 2010 to 2024 and 1,400 unique locations in 30 countries in Africa (materials and methods; figs. S1 and S2). After harmonization of overlapping spatio-temporal data to 716 unique GADM administrative level (ADM) 1 or 2 units (*24*), the analysis dataset consisted of 1,404 canonical cholera outbreaks (henceforth “episodes”) covering 71,063 location-months and an at-risk population of 506.2M (2025 reference (*25*), 33% of Africa’s population) (materials and methods; fig. 1a,b; figs. S2 to S5; tables S1 to S3). Laboratory-confirmed case data were rarely available (138/1,404 episodes, 9.8%, 13/30 countries), but outbreaks with confirmed cases had similar attack rates to other episodes although they were slightly longer, and had similar onset, duration and incidence rates under El Niño/La Niña years as all episodes (table S4).

**Fig. 1.**
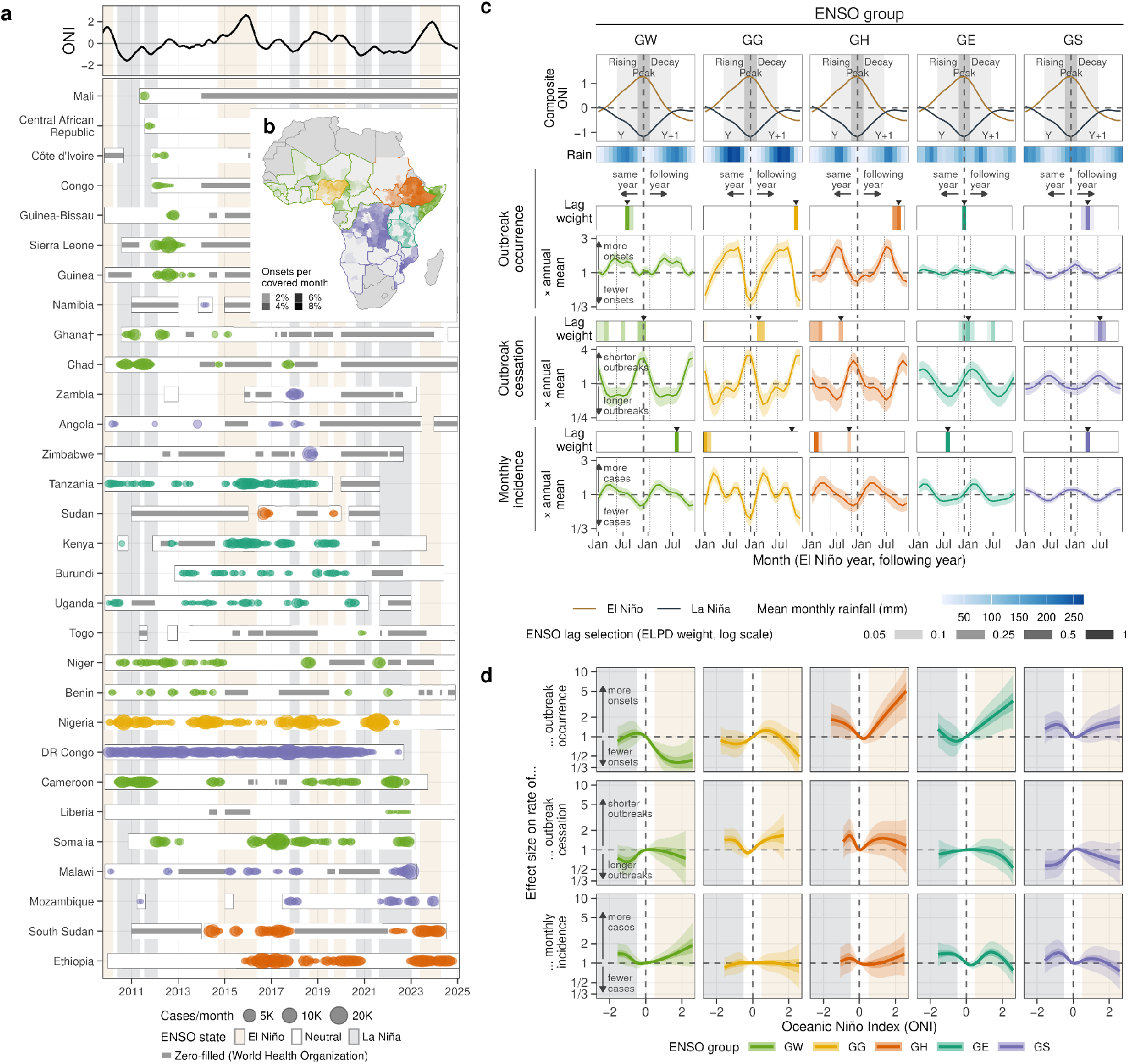
Cholera surveillance, seasonal timing and ENSO exposure–response in five African ENSO teleconnection groups. (**a**) Top: observed Oceanic Niño Index (ONI). Bottom: per country, periods covered by suspected-case surveillance (white boxes), including months zero-filled from World Health Organization national reports (grey bars), and monthly suspected cholera cases (circles, area proportional to cases), coloured by ENSO group (GW, West; GG, Guinean Gulf; GH, Horn; GE, East; GS, South). Countries are ordered by their last recorded case; daggers mark countries whose group membership is uncertain (maximum membership probability below 0.6; fig. S6). Shading marks El Niño (sand), La Niña (slate) and neutral (white) conditions (NOAA episodes: ONI ≥ +0.5 or ≤ −0.5 for at least five consecutive overlapping seasons). (**b**) Outbreak onsets per surveillance-covered month in each modelled subnational unit (opacity), coloured by ENSO group, with each group’s countries outlined in its colour; dark grey, countries not modelled; light grey, modelled countries without a subnational estimate. (**c**) Seasonal timing by ENSO group (columns), on a two-year calendar from January of the El Niño year (Y) to December of the following year (Y+1). Top row: composite ONI cycle, the mean ONI of the 22 El Niño (brown) and 19 La Niña (dark blue) episodes of 1950–2026, each aligned so that its peak falls in the Y/Y+1 winter; boxes mark the rising (June–October Y), peak (November Y – January Y+1) and decay (February–June Y+1) phases. Second row: mean monthly rainfall (CHIRPS 1990–2019 climatology of each first-level unit, population-weighted). Then, per response (outbreak occurrence, outbreak cessation, monthly incidence), two rows. The upper row is the weight of each single-lag ENSO model (pseudo-Bayesian model averaging; ELPD, expected log predictive density, by PSIS-LOO; opacity on a log scale, below 0.05 blank). Each lag is drawn at the outcome month that the December ONI informs: positive lags (following ENSO year) right of the dashed peak line, negative lags (leads; same ENSO year) left of it. Triangles mark the deployed lag, the best-ranked lag at or after −8 months (fig. S7). The lower row is the fitted seasonality relative to the annual mean: the cyclic seasonal spline for occurrence, and the annual plus semi-annual calendar harmonics for cessation (rate ratio > 1, shorter outbreaks) and incidence. Lines, posterior means; bands, 95% credible intervals. (**d**) Population-level exposure–response at the deployed lag: rate ratio relative to ONI = 0 for outbreak occurrence, outbreak cessation (as in (c)) and monthly incidence, by ENSO group. Lines, posterior medians; bands, 50% (dark) and 95% (light) credible intervals; curves span each group’s fitted ONI range; shading at ONI ±0.5 as in (a). Country-level curves: figs. S12 to S14.

Long-range associations between ENSO and cholera may be masked by population immunity or blurred by stochasticity in the location and timing of outbreak occurrence (*26–28*). We developed a Bayesian hierarchical statistical modeling framework to overcome these limitations—quantifying the direction, strength, and timing of ENSO effects on cholera outbreaks across Africa at the ADM1-2 level with spatial pooling of information (materials and methods; table S5). We independently assessed the effect of ENSO on outbreak occurrence (onset hazard), duration (cessation hazard), and monthly incidence rates (fig. S1). The framework explicitly allows non-linear ENSO effects to capture La Niña/El Niño asymmetries, and accounts for cholera seasonality (*29*), time since last outbreak as a proxy for immunity effects, as well as spatial coupling of outbreaks (materials and methods; table S6).

The outbreak occurrence model, which carried the strongest empirical signal, was used to identify spatial groups of countries in Africa with similar ENSO teleconnection profiles, with joint inference on the ENSO effect, the lag at which it acts, and group membership (materials and methods; fig. 1b,c,d; fig. S6). The best-fitting model identified five ENSO groups labelled by geographical centroids: West [GW], Guinean Gulf [GG], Horn [GH], East [GE] and South [GS] (fig. 1b; fig. S6; tables S7 and S8). We found close alignment between the seasonality of outbreak onsets, duration and monthly incidence rates, and the timing of each group’s rainy season (fig. 1c), consistent with evidence on the association between cholera excess risk and precipitation in Africa (*29*).

The Oceanic Niño Index (ONI), which quantifies ENSO, has an established yearly pattern, developing during the boreal summer (June–Oct) and peaking at the end of the year (Nov–Jan) (fig. 1c). The inferred time lag between ENSO and cholera outbreak characteristics consistently reflected the offset between the seasonal patterns of ONI, cholera and rainfall across groups, but differed in whether outbreaks responded to developing El Niño/La Niña events (same-ENSO-year response) or to events of the previous year (following-ENSO-year response). For outbreak occurrence, ENSO was linked to same-year responses in GW and GE and following-year responses in GG, GH and GS, with the best-supported lags matching the principal rainy season peaks in most groups, except in GE where it matched the short-rainfall peak in Nov–Feb (fig. 1c; fig. S7). Outbreak duration and monthly incidence rates followed similar ENSO timing patterns, with exceptions for following-year response of incidence in GW and same-year response of duration and incidence in GH (table S9; non-ENSO terms and sampler diagnostics: figs. S8 to S11).

We find that strong ENSO conditions have markedly different effects on cholera across Africa (fig. 1d; figs. S12 to S14). Our results demonstrate that during strong El Niño events (ONI at +1.5), outbreaks are twice as frequent in GH and GE, and 50% more frequent in GS (occurrence Incidence Rate Ratio [oIRR], GH: 2.3, 95% Credible Interval [CrI]: 1.1-4.2; GE: 2.1, 95% CrI: 1.1-3.9; GS: 1.5, 95% CrI: 0.9-2.4), with no statistically significant differences in duration (cessation IRR [cIRR], GH: 1.5, 95% CrI: 0.8-2.8; GE: 0.9, 95% CrI: 0.6-1.5; GS: 0.8, 95% CrI: 0.5-1.3) or incidence (monthly suspected case IRR [mIRR], GH: 1.1, 95% CrI: 0.6-1.9; GS: 1.1, 95% CrI: 0.7-1.8; GE: 1.5, 95% CrI: 0.95-2.2). El Niño has the opposite effect in GW (Western Africa excluding Nigeria plus Somalia), with around 60% lower onset risk (oIRR: 0.4, 95% CrI: 0.2-0.7), although the outbreaks that do occur tend to have 30% higher incidence rates (mIRR: 1.3, 95% CrI: 0.9-1.9); GG has a damped outbreak response. Strong La Niña (ONI at -1.5) had effects in the same direction as strong El Niño in GH and GS, although weaker (oIRR, GH: 1.8, 95% CrI: 1.0-3.2; GS: 1.3, 95% CrI: 0.8-2.2), with longer outbreaks in GS (cIRR: 0.6, 95% CrI: 0.35-0.95), while effects were close to null in GE (oIRR: 1.3, 95% CrI: 0.7-2.3; cIRR: 0.9, 95% CrI: 0.5-1.4; mIRR: 1.4, 95% CrI: 0.9-2.1). In GW, strong La Niña did not reduce outbreak onsets (oIRR: 0.9, 95% CrI: 0.5-1.5), but outbreaks tended to be more severe (mIRR: 1.4, 95% CrI: 0.96-2.0).

Within each ENSO teleconnection group, cholera outbreak associations with ONI were heterogeneous at national (figs. S12 to S14) and subnational levels (fig. 2a). When defining ENSO sensitivity as having statistically significant adverse effects greater than 25% on any of the outbreak dimensions (occurrence, duration, or incidence rates; materials and methods), we find that two-fifths of the covered population in Africa (40%, 201M of 506M) lives in ADM1 units sensitive to strong El Niño events (ONI at +1.5), covering most of the population in GE (72%) and GH (82%), with numbers rising to 43% (217M) under very strong El Niño (ONI at +2) (fig. 2b). A smaller share of the population (27%, 139M) is sensitive to strong La Niña conditions (ONI at -1.5), but in different ENSO groups: GH (82% of its population) and GW (20%).

**Fig. 2.**
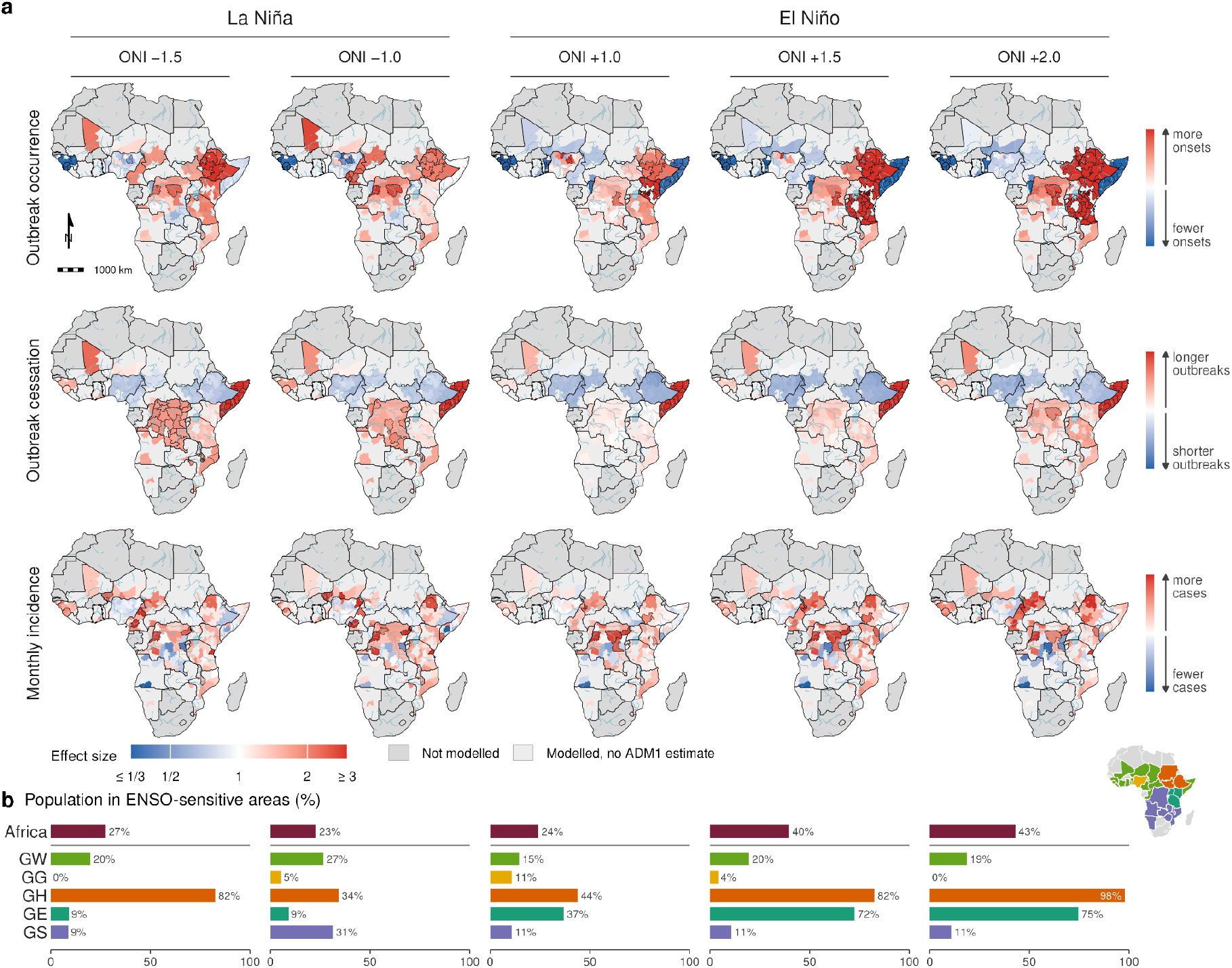
Subnational ENSO effects on cholera outbreak occurrence, duration and incidence, and the population living in ENSO-sensitive areas. (a) Rate ratio at five ONI values (La Niña: −1.5, −1.0; El Niño: +1.0, +1.5, +2.0) relative to ONI = 0, per first-level administrative unit (ADM1). Each ratio combines the population-level exposure–response with the country- and ADM1-level random deviations, each ENSO group at its deployed lag (Fig. 1c). Rows: outbreak occurrence, outbreak cessation (inverse rate ratio, so red marks longer outbreaks) and monthly incidence. Colours are capped at 1/3 and 3; black outlines, units whose 95% credible interval excludes 1; greys as in Fig. 1b. (**b**) Share of the population living in ENSO-sensitive ADM1s at each ONI value, for Africa and each ENSO group (inset; groups as in Fig. 1). An ADM1 is ENSO-sensitive when, for at least one response, the posterior probability of an adverse effect above 25% (more onsets, longer outbreaks or more cases) is at least 0.95.

To quantify how ENSO timing and strength affect key cholera outbreak dimensions, we developed a novel stochastic disease-agnostic outbreak simulation framework that tackles the persistent question of how to attribute changes in disease burden to climatic events like El Niño and La Niña. The simulator ingests the posterior distributions of our statistical models along with historical ONI data to produce cholera outbreak simulations (materials and methods). Using modern filtering methods for coupled dynamical systems (*30*), we reconstruct with high fidelity the historical time series of cholera outbreak states and incident cases across the continent (fig. 3a; figs. S15 to S17). This generative model is then used for counterfactual simulations with matched ENSO-neutral analogue ensemble trajectories (materials and methods). The difference between simulations driven by the observed ENSO and the ENSO-neutral counterfactuals enables us to attribute burden to major El Niño/La Niña events during our study period (materials and methods; fig. 3a; table S10). In-sample hindcast simulations (2012–2022) supported the use of the simulator for forecasting and counterfactual comparison, showing robust performance against climatology for outbreak onsets and monthly case counts in most ENSO groups (figs. S18 and S19).

**Fig. 3.**
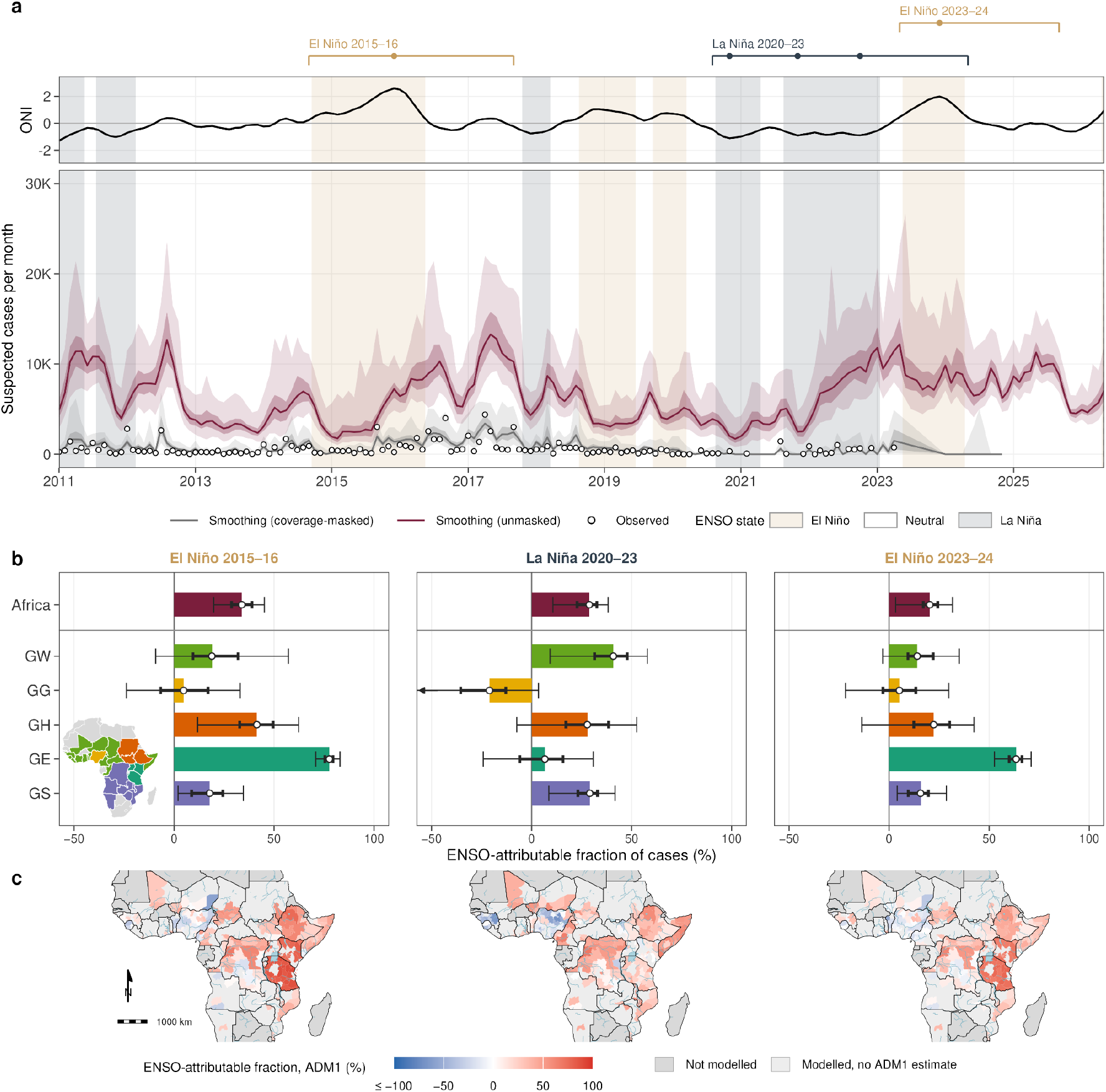
Attribution of cholera cases during past strong ENSO events. (a) Top: observed ONI, shaded as in Fig. 1a, with each event’s window (bracket, from launch to the last window end; dots, ONI peaks). The 2020–23 La Niña is one event with three winter peaks (event windows: materials and methods). Bottom: monthly suspected cases in Africa from the historical reconstruction (particle-filter smoothing; “smoothing” in the legend) over all modelled units (dark red) and over surveilled unit-months only (grey), with the observed cases in surveilled unit-months (circles). Lines, medians; bands, 50% and 95% intervals across 100 reconstruction replicates. (b) ENSO-attributable fraction of suspected cases (AF = 1 − CF/F) per event (columns), for Africa and each ENSO group. F is the factual simulation driven by the observed ONI from the launch; CF is the counterfactual in which the ONI is replaced by pre-industrial-control analogue trajectories that are ENSO-neutral in every December–February the window reads. Both start from the same reconstructed state; each replicate runs the posterior draw of its starting reconstruction replicate (100 draws × 100 replicates per arm), and every replicate is kept. Bars and circles, posterior medians; thick and thin bars, 50% and 95% intervals across posterior draws, noise-adjusted. (c) AF of the replicate means per ADM1, capped at −100%; greys as in Fig. 1b. Outbreak onsets and the other factual readings: fig. S20.

After accounting for stochasticity in outbreak dynamics, we find that 34% (95% CrI: 20-45) of suspected cholera cases occurring during outbreaks were attributable to the very strong 2015–16 El Niño event (peak ONI 2.6; fig. 3a) and primarily concentrated in GE and GH (fig. 3b; fig. S20). Similarly, 29% (95% CrI: 11-38) of cases were attributable to the multi-year 2020–23 La Niña event (minimum ONI -1.1), with highest attributable fraction in GW, GH and GS, consistent with previous reports (*18*). The very strong 2023–24 El Niño event (peak ONI 2.0) had a smaller impact, with only 20% (95% CrI: 3-32) of cases attributable to ENSO.

These historical events foreshadow an increase in cholera burden linked to the very strong record-breaking El Niño predicted to peak by the end of 2026 (*22*) (materials and methods; fig. 4a; fig. S21). Applying our generative simulation framework to available ONI forecasts (issued October 2026; materials and methods), we project burden through to October 2027, predicting an annualized cumulative incidence of 22.0 (95% prediction interval [PI]: 15.2-38.6) suspected cholera cases/100,000/year (173,600 cases, 95% PI: 119,700-305,000), close to double the 2011–2020 mean annual incidence rates in Africa (*31*), with the highest incidence rates in GH (26.2, 95% PI: 12.7-62.7), GW (25.1, 95% PI: 7.7-81.4) and GE (22.0, 95% PI: 14.9-31.0) (fig. 4a; table S11; figs. S22 and S23).

**Fig. 4.**
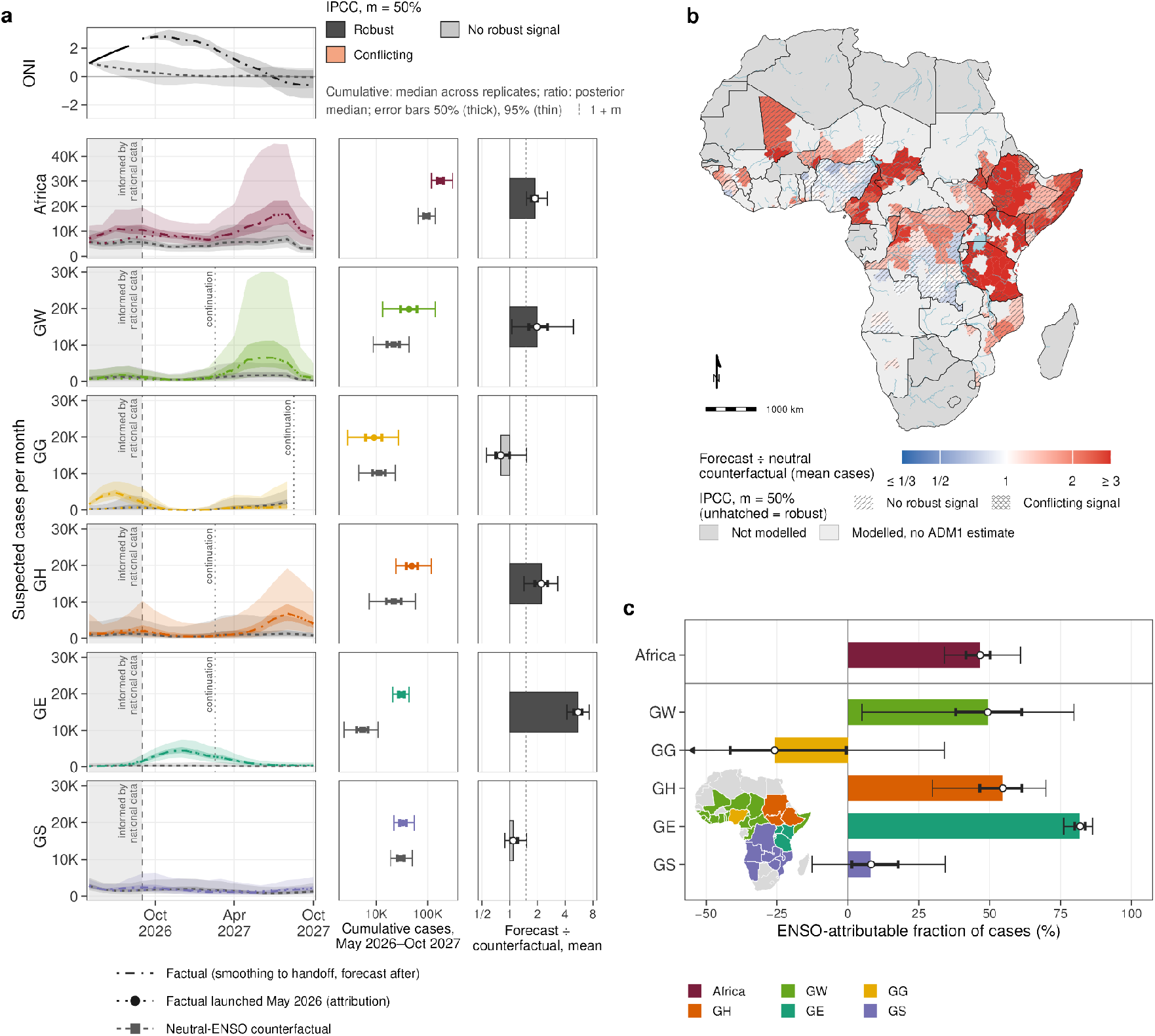
Cholera burden in Africa and the 2026–27 El Niño event. (a) Top: observed ONI (solid), CFSv2 ensemble forecast continued by pre-industrial-control analogues (dot-dashed) and the mean of the neutral-ENSO counterfactual analogues (dashed), with 95% bands (fig. S21). Left: monthly suspected cases from May 2026 to October 2027 for Africa and each ENSO group, in three arms started from the same reconstructed state. The factual run (dot-dashed) follows the historical reconstruction (as in Fig. 3a; shaded) up to the September 2026 handoff (dashed line) and the forecast after it. The attribution factual (F, dotted) and the neutral-ENSO counterfactual (CF, grey, dashed) are both launched in May 2026. A vertical dotted line marks each group’s firm horizon, after which its deployed lags read ENSO past the end of the CFSv2 forecast (June 2027) from the analogue continuation (veiled; “continuation”). Lines, medians; bands, 50% and 95% intervals across replicates. Middle: cumulative cases over the same period, F (coloured circles) and CF (grey squares); medians with 50% and 95% intervals across paired replicates. Right: ratio of mean F to mean CF cases; circles and intervals as in Fig. 3b; bars filled by the IPCC AR6 robustness class at m = 50% (robust: ≥ 66% of paired replicates exceed (1 + m) × CF and ≥ 80% exceed CF; conflicting: only the first holds); solid line, ratio 1; dotted line, 1 + m. (b) Ratio of mean F to mean CF cases per ADM1 over the window, capped at 1/3 and 3; diagonal hatching, no robust signal; cross-hatching, conflicting signal; unhatched, robust (classes as in (a)); greys as in Fig. 1b. (c) ENSO-attributable fraction of cases, for Africa and each ENSO group; bars, circles, intervals, draws, replicates and replicate rule as in Fig. 3b. Per-country series: figs. S22 (cases) and S23 (onsets); other analogue sets: figs. S24 and S25.

This translates to a burden twice that of ENSO-neutral counterfactual conditions (mean factual-to-counterfactual ratio: 1.9, 95% CrI: 1.5-2.6; other counterfactual analogue sets: figs. S24 and S25). Notably, the greatest increases are projected in ENSO groups GE and GH (mean factual-to-counterfactual ratio: GE: 5.5, 95% CrI: 4.2-7.3 and GH: 2.2, 95% CrI: 1.4-3.3). Following these predictions, 47% (95% CrI: 34-61) of all cholera cases through October 2027 will be attributable to the 2026–27 El Niño event, with strongest attributable fractions in GE (82% of the cases, 95% CrI: 76-86) and GH (55% of the cases, 95% CrI: 30-70). We find that 271M (95% CrI: 235-302, 54% [46–60]) people live in ADM1 units with statistical evidence for a 50% or more increase in cholera burden compared to ENSO-neutral conditions (table S11). Among these, 105M live in ADM1 units with a robust signal following IPCC attribution methods (materials and methods; fig. 4b), most of whom live in Ethiopia and Tanzania (table S11).

With over 170,000 suspected cholera cases projected across Africa through October 2027 and nearly half attributable to this year’s El Niño, the pressure on already-stretched global resources is acute. Oral cholera vaccine shortages forced the WHO to permit single-dose, as opposed to standard two-dose, campaigns in 2022 to extend population reach. While supply has since partially recovered, the scale of this projected ENSO-driven risk will likely test the stockpile and other control resources again (*32*). Nevertheless, thoughtful targeting can be achieved, as our results on the geography of ENSO-driven risk point to regions where El Niño anticipatory action has the largest expected benefit.

Prepositioning or earmarking limited resources like vaccines for countries in Eastern Africa should be a strong priority for late 2026, especially since ENSO precedes outbreak onsets by 0 (GE) to 10 months (GH). Further, enhancing the readiness to detect and respond to outbreaks in Western Africa and Somalia is critical, as this region may experience fewer outbreaks with higher incidence rates for those that do occur. Available data in Western Africa (GW) as of September 2026 strongly align with our ENSO-informed predictions, although with different levels of agreement at the country level (fig. S26). Importantly, although strong El Niño events typically hinder cholera outbreaks in GW, they are often followed by La Niña the next year (*33*), which in turn increases outbreak risk and severity for that cholera season, especially in Somalia. These dynamics thus call for an ENSO-informed dynamic allocation of regional-level resources for cholera control.

This study brings together multiple state-of-the-art approaches that advance on the recurring question of how to attribute changes in disease burden to climatic events in a dynamic modeling and simulation framework that may be extended to other outbreak-prone climate-sensitive diseases, particularly as access to high-resolution surveillance data expands. We however note several methodological limitations (supplementary text). Our predictions hinge on the stationarity of the inferred ENSO–cholera associations over the study period, which is supported by the consistency of in-sample hindcast skill of our simulation framework (figs. S18 and S19). More importantly, our predictions may very well be invalidated by timely and coordinated public health action to stem outbreak transmission, and we hope this study will contribute to that end.

Taking forward the legacy of the seminal single-location ENSO–cholera study in Bangladesh (*13*), we here resolve ENSO effects on subnational cholera outbreaks at the continental scale in Africa, identifying distinct ENSO–cholera regions and quantifying for the first time the timing, direction and magnitude of ENSO effects on outbreak onset, duration and size. The alignment of ENSO effects and cholera outbreaks with regional rainy seasons points to rainfall anomalies as a likely mediating pathway, though the specific hydroclimatic mechanisms remain to be fully resolved (*34*). As anthropogenic warming is projected to amplify ENSO-related rainfall variability and alter its teleconnections (*1*, *5*, *35*), anticipating how climate drives disease burden can only crescendo in public health importance—for cholera and beyond.

## Data Availability

The analysis code, the derived model inputs, the monthly WHO national series and the GitHub data folder cited in the supplementary materials are publicly available at https://github.com/GenevaIDD/enso_cholera_africa and archived at [Zenodo DOI upon acceptance]. Outbreaks were extracted from the cholera time-series database with the OutbreakExtractR package, available at https://github.com/HopkinsIDD/OutbreakExtractR. The cholera outbreak datasets contain public and confidential data and were derived from the Cholera Taxonomy database (https://cholera-taxonomy.middle-distance.com/). Public versions of the outbreak dataset are available on OSF at https://osf.io/2ncf7. The files of data S1 accompany this manuscript and are described in the supplementary materials. The ENSO index, the ENSO forecasts, the pre-industrial ENSO trajectories, the population estimates and the administrative boundaries are publicly available from the sources cited in the supplementary materials (materials and methods).

https://osf.io/2ncf7

## Acknowledgments

Claude Code (Anthropic; Claude Opus 4.6, Opus 4.8, Opus 5 and Opus 5.5, Sonnet 4, Sonnet 4.6, Sonnet 5 and Sonnet 5.5, and Fable 5.1 and 5.2; May–November 2026) was used throughout the study investigation as a coding aid, to draft parts of the supplementary text and figure captions, to transfer numerical results from the analysis outputs into the manuscript, to check consistency between text, tables and figures. OpenAI Astra 6 was used for a secondary code review. The authors reviewed and revised all such content and take full responsibility for the manuscript.

The authors would like to thank all the ministries of health, WHO offices, and operational partners including the UNICEF WCAR and ESAR offices for contributing data to the database over the past decade in addition to all previous team members who assisted with data validation and entry.

## Author contributions

Conceptualization: JPS, ASA

Methodology: JPS, BZ

Investigation: JPS, QZ

Visualization: JPS

Funding acquisition: ECL, ASA

Project administration: ECL, ASA

Supervision: ECL, ASA

Writing – original draft: JPS

Writing – review & editing: JPS, ECL, RK, QZ, BZ, ASA

## Supplementary Materials

### S1 Materials and Methods

The analysis has three parts (fig. S1). First, we extracted cholera outbreaks from sub-national surveillance records, mapped them to administrative units and built three analysis datasets, one each for outbreak occurrence, duration and monthly incidence (Section S1.1). Second, we grouped countries by their response to the El Niño–Southern Oscillation (ENSO) with a latent-class model, and fitted for each ENSO group three Bayesian hierarchical models of outbreak occurrence, duration and monthly case counts, each at an ENSO lag selected by cross-validation (Sections S1.2 to S1.5). Third, we used the fitted models in a stochastic simulator: a particle filter reconstructs the outbreak state of every unit up to August 2026, and forward runs under factual and counterfactual ENSO conditions attribute outbreaks to past ENSO events and to the current ENSO forecast, with hindcasts to assess forecast skill (Section S1.6).

**Fig. S1.**
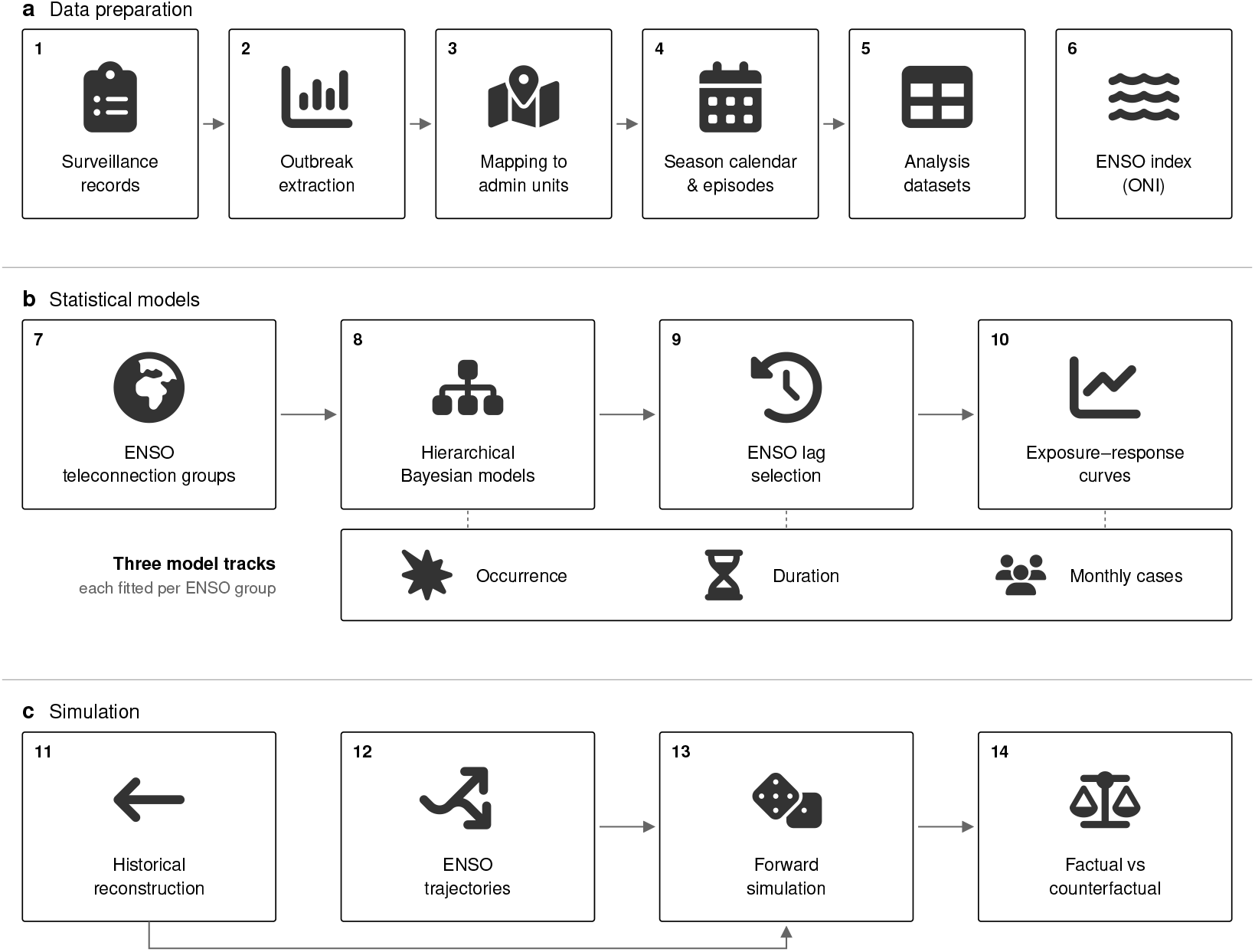
Analysis workflow. (**a**) Data preparation: sub-national surveillance records, complemented by WHO national data (1), give outbreaks (2) that are mapped to administrative units (3) and consolidated into one episode per unit and cholera year (4), forming the occurrence, duration and monthly case-count datasets (5); ENSO is measured by the Oceanic Niño Index at lags of *−*11 to +11 months (6). (**b**) Statistical models: a latent-class model partitions the countries into five ENSO groups (7); hierarchical Bayesian models are fitted for each group and analysis (8), at lags selected by cross-validation (9), giving exposure–response curves (10). (**c**) Simulation: a particle filter reconstructs the outbreak state of every unit up to August 2026 (11); forecast and counterfactual ENSO inputs (12) drive forward simulations (13), whose difference gives the outbreak burden attributable to ENSO (14). Icons: Font Awesome Free 6.5.2 (Fonticons, Inc.), CC BY 4.0.

#### S1.1 Cholera outbreak data

##### S1.1.1 Source surveillance data

We used outbreak data from the Global Cholera Taxonomy Database, a curated compilation of suspected cholera surveillance reports [36]. Each record gives the number of suspected cases, and where available confirmed cases and deaths, over a reporting interval in a reporting geography. We call a reporting geography a location period: a named area with a fixed boundary over a given period. We retrieved records for all countries of the World Health Organization (WHO) African and Eastern Mediterranean regions with reporting intervals between January 2010 and December 2024, together with the boundary of every location period.

##### S1.1.2 National surveillance data

We complemented the sub-national database with three national WHO sources. The first is the annual cholera report of the Weekly Epidemiological Record (WER), which gives country totals of suspected cases for 2010–2024 [37]. The second is the WHO global cholera dashboard [38], which gives weekly national counts of suspected cases from January 2023 and national annual totals for 2023–2024. The third is the WHO multi-country cholera situation reports [39], which give cumulative year-to-date national totals.

We used these sources in two ways. First, annual country totals identify country-years in which an outbreak cannot have occurred (Section S1.1.8). For 2023–2024 we took the larger of the WER and dashboard totals where both report. Second, we built a monthly national series of suspected cases. Weeks without a report are gaps, and only complete months are retained. Where the dashboard has no weekly report but consecutive situation reports give year-to-date totals at intervals of at most 45 days, we differenced the two cumulative totals and allocated the difference to the months of the interval between them. A month is read as without cases when WHO reports an explicit zero over the last 28 days. An unchanged cumulative total is read as a month without cases only for Tanzania, whose Ministry of Health declared the end of its national cholera outbreak in February 2026. The resulting monthly national series, complete from January 2023 to August 2026 with partial counts for September 2026, is released as GitHub data folder, file `D36_who_national_monthly`. These national-level observations enter the historical reconstruction as a country-level observation (Section S1.6.8).

##### S1.1.3 Cholera outbreak extraction

We identified outbreaks building on the operational outbreak definition of Zheng et al. [36], which we applied to each country separately. In short, an outbreak in a location is a period of weekly incidence above the location’s mean weekly incidence that totals at least 50 suspected cases.

We retained weekly and sub-weekly records at national to third administrative level, and removed records without a suspected case count. Sub-weekly records were aggregated to weeks, and duplicate records for the same location and week were averaged. Following our previous approach [36], we set weeks without a record within a location’s series to zero suspected cases, treating an unreported week as a week without cases, and extended each series by eight zero weeks before its first and after its last record.

Each location period was assigned the population inside its boundary from constrained 100 m WorldPop grids [25] for the median year of its records. The grids cover 2015–2030. For a year outside that range we used the nearest available year, rescaled by the change in the national total of the United Nations World Population Prospects 2024 (WPP) [40] while holding sub-national shares fixed. We rescaled gridded totals to the national WPP totals unless the rescaling factor fell outside 0.5–2.

We define the outbreak threshold of a location as the mean weekly incidence, in suspected cases per person, over its whole zero-filled series. A week is above threshold when its incidence is at least the threshold and at least one case was reported. An outbreak starts at the first of two consecutive above-threshold weeks, and ends at the last week before a run of six consecutive below-threshold weeks. The closing run decides that the outbreak is over but is not part of it, so episode end dates, durations and case totals exclude it. Two starts that are not separated by such a run belong to the same outbreak, and an outbreak without a closing run lasts until the end of the series. Finally, we discarded outbreaks with fewer than 50 suspected cases in total. An example of the outbreak definition is shown in fig. S2.

**Fig. S2.**
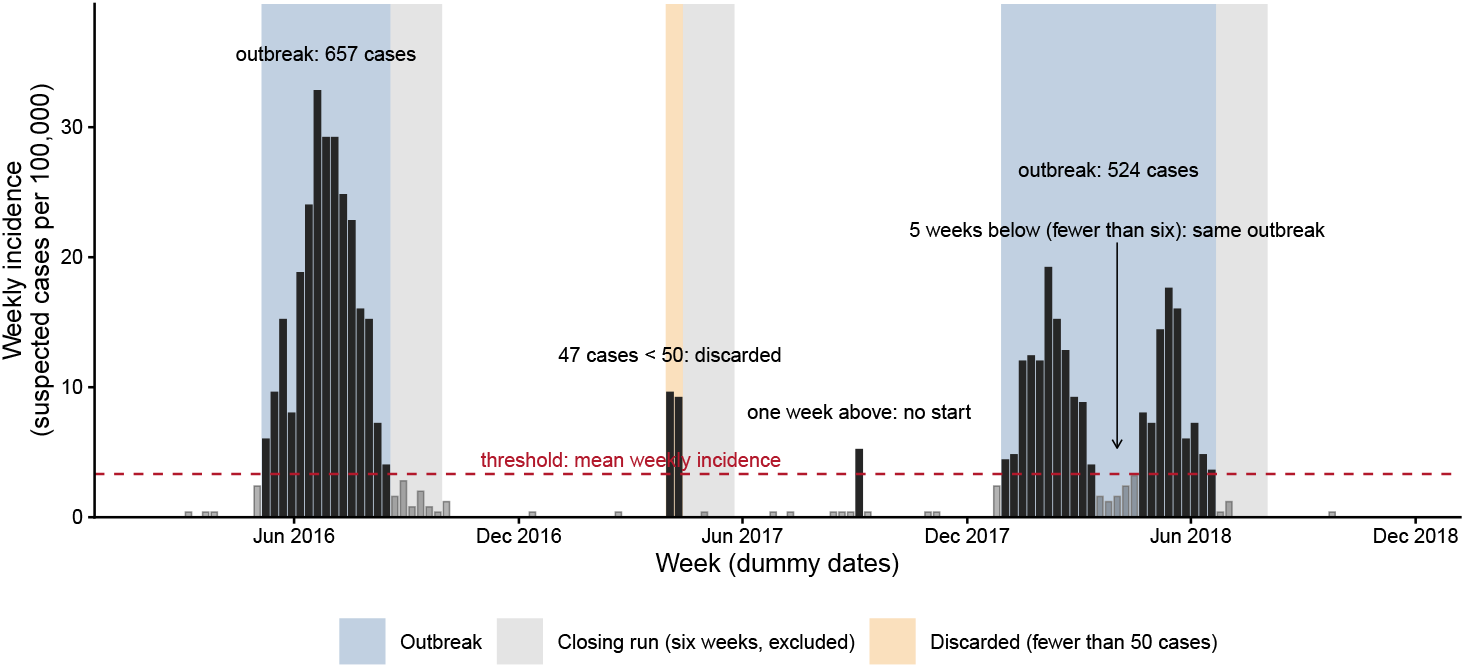
Outbreak extraction, worked example. Weekly suspected-cholera incidence of one location period, on a dummy series simulated for this figure (156 weeks, population 250,000; not surveillance data). Weeks without a record are zero, and eight zero weeks pad both ends of the series. The outbreak threshold (dashed red line) is the mean weekly incidence over the whole zero-filled series; weeks at or above it with at least one case (dark bars) are above threshold, the others (light bars) below. An outbreak starts at the first of two consecutive above-threshold weeks and ends at the last week before six consecutive below-threshold weeks; that closing run (grey) decides the end but is excluded from the outbreak’s dates, duration and case total. A run of fewer than six below-threshold weeks inside an outbreak does not end it, so the two waves of the second outbreak form one outbreak, and a single above-threshold week does not start one. Outbreaks with fewer than 50 suspected cases in total are discarded (orange): the two-week cluster in the middle of the series meets the start rule but stays below the 50-case floor.

##### S1.1.4 Episode table

We call an episode the record of one outbreak in one reporting geography: its start date, its end date and its suspected cases by month. We then aggregate these into one episode per administrative unit and cholera year (Section S1.1.7). We allocated case counts to calendar months by splitting the cases of each weekly record across the calendar months it overlaps.

##### S1.1.5 Mapping reporting geographies to administrative units

Location periods rarely coincide with a single administrative unit. We therefore intersected each location period with first- and second-level administrative units from the GADM database [24]. Throughout, a unit is a GADM administrative unit. We computed the population of each intersection from the same WorldPop surface. We write *H*[*l, g*] for the population living in the intersection of location period *l* and GADM unit *g*, *P_l_* for the population of *l*, and *Q_g_* for the population of *g*, and define two shares:

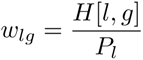 (share of *l*’s population living in *g*), 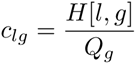 (share of *g*’s population covered by *l*).

We discarded intersections holding less than one person or less than 0.01% of *P_l_*, renormalising the weights *w_lg_*.

We derived the administrative level at which each location period is represented from its geometry. We first identified the best-covering level, and accepted a level one step coarser when it lost little coverage, using a coverage floor of 80% and a tolerance of 10%. We define the dominant coverage of a location period as max*_g_ w_lg_*, the share of its population living in its most populous unit, and call a location period composite when this is below 0.90. We excluded composite location periods from all analyses (55 of the 1,407 location periods with an extracted outbreak, 3.9%; fig. S3), and assigned every other location period to its dominant unit.

Population denominators in the models are unit populations by year from the same WorldPop surface and WPP rescaling, with the same nearest-year rule outside 2015–2030.

##### S1.1.6 Cholera-season calendar

Cholera transmission in much of Africa follows an annual cycle whose phase varies by location [29]. We accounted for this seasonality by assigning each location period a trough month, the calendar month of lowest mean seasonal transmission in Perez-Saez et al. [29]. We matched estimates at the location’s own unit, then at its first-level unit, and then at the nearest unit of the same country, and set the trough month to January for location periods without any match (57 of the 2,435 sub-national location-period episodes, before the exclusions of Section S1.1.7). Each administrative unit then receives a single calendar, given by the most frequent trough month among the location periods reporting on it. We then define each unit’s cholera year as running from trough to trough, and label a cholera year by the calendar year in which it starts, so that episodes of early 2010 can belong to cholera year 2009.

##### S1.1.7 Canonical episodes

A unit can be reported on by several location periods at once, for example by a district and a province report, or by successive versions of a boundary. We therefore aggregate location-period episodes into one episode per unit and cholera year. The episode starts at the earliest member start and ends at the latest member end. Its size is the total at the finest reporting level, choosing the level with the largest total, so that cases reported at two levels are not counted twice. A unit therefore has at most one onset per cholera year. We define the months at risk of a unit as the months covered by the surveillance series of at least one location period reporting on it.

National-level records are removed before aggregation, together with later outbreaks of a location period in the same cholera year and composite location periods (fig. S3). Aggregation then reduced the remaining 2,195 location-period episodes to the final analysis dataset of 1,404 episodes, of which 365 combine several location periods, cumulating 1,109,769 suspected cases in 716 GADM units of 30 countries, from January 2010 (first episode start) to November 2024 (last episode end) (fig. S3, tables S1 and S2, figs. S4 and S5).

**Fig. S3.**
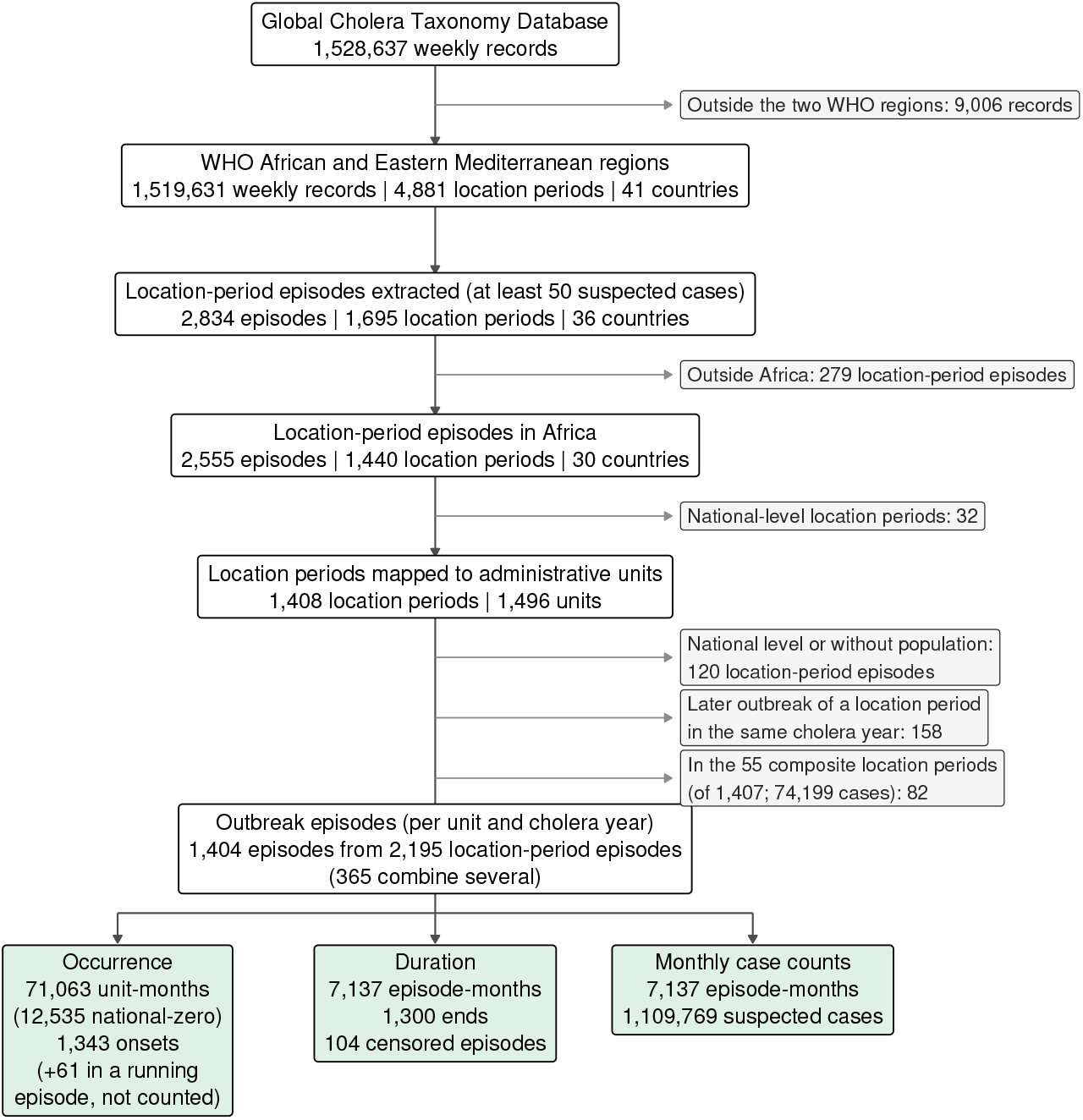
Data processing cascade. Records, outbreaks and episodes retained by each processing step, from the weekly records of the Global Cholera Taxonomy Database to the three analysis datasets (Sections S1.1.3 to S1.1.8). Side boxes give what leaves the cascade at a step: records outside the WHO African and Eastern Mediterranean regions; location-period episodes outside Africa; national-level location periods; and, before consolidation, national-level location-period episodes and those without a population estimate, later outbreaks of a location period in the same cholera year (left out of the occurrence dataset) and the episodes of the composite location periods. One mapped location period loses all its episodes at the step that removes national-level episodes and those without a population estimate, so the composite location periods are counted out of one fewer location period than are mapped. The cascade ends in canonical outbreak episodes and forks into the occurrence, duration and monthly case-count datasets, whose counts are those of the prepared datasets, before the duration and monthly models drop first-level episodes that overlap a second-level episode. Onsets that fall inside a running episode of an earlier cholera year of the same unit are not occurrence events, but their episodes remain in the duration and monthly datasets.

##### S1.1.8 Analysis datasets of outbreak occurrence, duration and monthly incidence

We derived three datasets from the episode dataset, one for each of the three dimensions of a cholera outbreak: when it starts (onset timing), how long it lasts (duration) and how many cases it produces each month (monthly incidence rate). The occurrence dataset has one observation per unit and calendar month while the unit is at risk, with an onset indicator as outcome and the number of days at risk in the month as exposure. It includes every second-level unit with at least one recorded episode in 2010–2024, and the first-level units with no reporting second-level unit. Units without a recorded episode never enter the dataset or the simulations (fig. S5).

As months at risk follow sub-national surveillance coverage, a country-month without an active sub-national series contributes nothing, even when national surveillance (Section S1.1.2) shows that the country had no cholera. We therefore added national-zero months, country-months in which the whole country had no suspected cholera, as no-onset unit-months for every unit of the country. We used national-level data (Section S1.1.2) to impute zeros, accounting for both monthly-level national zeros when available and annual-level observations of fewer than 60 cases (the outbreak definition limit plus 20% tolerance), as well as our sub-national data. Country-years in which WHO reported fewer than 60 suspected cases also qualify, with two edge guards. The last four months of such a year are claimed only if the next year is also below the threshold, and the first four months only if the previous year is also below it or has no WHO figure, so that an epidemic counted by WHO but not recorded in the database cannot run into a zero January to April. A WHO year whose database episode cases exceed 1.2 times the WHO total is treated as under-reported, and its months are not claimed unless they rest on the national series alone. Ethiopia 2010–2016, whose WHO figures reflect reporting of acute watery diarrhoea rather than an absence of cholera, is excluded [41]. The window ends in December 2024, the last complete WHO annual-report year, and a national-zero month is never added inside a recorded episode of its unit. This adds 12,535 unit-months without onset (17.6% of the 71,063 unit-months of the occurrence dataset).

We exclude data under the following conditions: (i) a unit is not at risk during the remaining months of an outbreak that began in an earlier cholera year; an onset recorded in those months is an episode for the duration and monthly models but not an occurrence event; (ii) where a first-level unit and its second-level units both report, the first-level unit-months are dropped, and the months of the second-level units inside a first-level episode with no overlapping second-level episode are censored, so that these units are not left at risk during a first-level outbreak; (iii) later onsets of the same location period in the same cholera year; (iv) re-dating duplicates; and (v) members absorbed into a canonical episode. Exclusion counts by country are given in GitHub data folder, file `D31_occ_panel_exclusions`. Over all countries, 61 onsets and 856 unit-months were removed from the risk set inside running episodes, and 670 second-level unit-months were censored under 96 first-level episodes without an overlapping second-level episode.

The duration dataset has one observation per episode and calendar month while the episode is active, with an indicator that the episode ended in that month as outcome and the number of active days in the month, floored at one day, as exposure. We consider episodes that end within 73 days of the end of their reporting series as right-censored: 31 days, plus the six weeks of the closing run, which is not part of the outbreak. This applies to 104 of the 1,404 episodes (7.4%). The monthly case-count dataset uses the same episode-months, with the number of suspected cases in the month as outcome and the unit population multiplied by the number of active days as exposure. Months without a report within an episode count as zero cases. As in the occurrence dataset, the duration and monthly models drop the 323 first-level episodes that overlap a second-level episode, and are fitted to the remaining 1,081 episodes and 5,273 episode-months.

##### S1.1.9 Descriptive summaries by region and ENSO state

#Tables S1 to S3 describe the canonical episodes of the analysis set without a model. They group countries by World Health Organization African Region (WHO-AFRO) subregion: Western, Central, Eastern and Southern Africa. Somalia and Sudan, members of the WHO Eastern Mediterranean Region, are placed in Eastern Africa.

Each episode also takes the ENSO state of its cholera year (Section S1.1.6): the Oceanic Niño Index (Section S1.2.1) of the December–February season whose December lies in that cholera year, El Niño at or above +0.5 and La Niña at or below *−*0.5, with no persistence requirement. The record spans 16 cholera years, from 2009 (Section S1.1.6).

At the continental level, outbreaks were more common during La Niña (96.2 outbreaks/yr) than during El Niño (82.3 outbreaks/yr) and ENSO-neutral years (83.2 outbreaks/yr), with strong differences between WHO-AFRO subregions: in La Niña years, Western and Southern Africa saw more outbreaks (Western: 34.8/yr vs. 18.8/yr neutral; Southern: 5.5/yr vs. 0.2/yr neutral), while Eastern Africa (including Somalia and Sudan) had fewer (17.2/yr vs. 40.0/yr neutral) (table S3). Overall, cholera outbreaks had a median cumulative attack rate of 78 cases/100,000 (interquartile range [IQR]: 30–195), and lasted 14 weeks (median, IQR: 8–24), with first-level episodes being slightly longer and with lower burden (table S2). Overall, attack rates and duration differed across ENSO conditions (lower attack rates in El Niño and La Niña than in neutral years), but without a consistent pattern across subregions (table S3).

#### S1.2 El Niño–Southern Oscillation

##### S1.2.1 ENSO index and cholera outbreak-association lags

We measure ENSO with the official Oceanic Niño Index (ONI) of the NOAA Climate Prediction Center [42], the three-month running mean of ERSST sea-surface-temperature anomalies in the Niño 3.4 region (5°N–5°S, 120°–170°W) [43, 44]. Writing ONI*_t_* for the index of the three-month season centred on calendar month *t* (the December–February value is assigned to January), the lagged exposures are ONI*_t_*_−_*_ℓ_* for candidate lags *ℓ* = *−*11*, … ,* 11 months, of which lags below *−*8 months are not selected (Section S1.4.5). A negative lag *ℓ*reads the index *|ℓ|* months after the outcome month. As ENSO events develop over several months, the index after the outcome month can describe a developing event better than the index before it, and we therefore let the data choose between positive and negative lags. Each model uses a single lag *L*, which we select as described in Section S1.4.5.

We summarise the selected lag of each model by its sign. A positive lag *L* relates the outcome to the ONI *L* months earlier, so that cholera follows an event that has already developed (a *following-year* response, as events peak in December–February and cholera responds after the ENSO winter); a zero or negative lag (a lead) relates it to the ONI at or after the outcome, so that cholera responds as the event develops and peaks (a *same-year* response). The class is assigned per model, so a group can be same-year for one outcome and following-year for another.

#### S1.3 Statistical modelling framework

The statistical model aims to quantify how ENSO, at a group-specific lag, changes the rate at which outbreaks start, the rate at which they end and the number of cases they produce each month, accounting for seasonality, outbreak history, contagion and persistent differences between places. Every model is fitted separately for each ENSO teleconnection group (Section S1.4.1). We model outbreak occurrence, outbreak duration and monthly case counts with three Bayesian hierarchical models that share the same nested random effects, the same ENSO exposure basis and the same approach to anchoring the intercept on an empirical baseline rate. We first describe this shared structure, then each model, the anchoring, the priors and the estimation settings.

We use one set of symbols in the three models. Index *i* is an administrative unit (first- or second-level), *t* a calendar month, *k* an outbreak episode and *s* a posterior draw. For a unit *i*, *j*(*i*) and *c*(*i*) are the first-level unit and the country containing it; for an episode *k*, *i*(*k*) is the unit in which it was recorded, *t*_0_(*k*) its onset month and *d_kt_* = *t − t*_0_(*k*) + 1 its age in month *t*. *τ* (*t*) *∈ {*1*, … ,* 12*}* is the calendar month of the year of month *t*, *ℓ* a candidate ENSO lag and *L* the selected one, *h* = 1*, … , J* indexes the columns of the ENSO spline basis, *u* denotes a random intercept and *v* a random slope. We write *a_i_* for the empirical baseline rate of the cell (country and administrative level) containing unit *i* (Section S1.3.7); each model has its own baseline rate, written 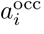, 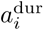 and 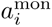 where the model is not clear from the context. Observations of the occurrence model are unit-months (*i, t*); observations of the duration and monthly case-count models are episode-months (*k, t*).

##### S1.3.1 Discrete-time hazard as a Poisson model

We estimate the occurrence and duration models as Poisson mixed models fitted to a unit-month expansion of the data, the standard approach for piecewise-constant discrete-time hazard models [45, 46].

For an interval (*i, t*) of length Δ*t_it_*days (a unit-month, or an episode-month for duration) with event indicator *y_it_* ∈ {0, 1*}* and a hazard *λ_it_* = exp(*η_it_*) constant within the interval, the event probability is

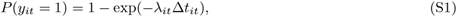

and we fit the log-linear Poisson form

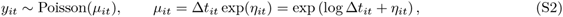

with log Δ*t_it_* as an offset. With at most one event per interval and Δ*t_it_* the time at risk, this likelihood is proportional to the piecewise-exponential survival likelihood with right-censoring [45, 46], and it agrees with the grouped-time likelihood (S1) [47] to first order in *λ_it_*Δ*t_it_*, as monthly onset and cessation probabilities are small. The simulator draws events from (S1) (Section S1.6.3).

##### S1.3.2 Hierarchical structure

All three models include nested random intercepts, with random slopes on selected covariates. The nesting reflects the expectation that two administrative units in the same country are more alike than two units in different countries, and the random slopes that the effect of a covariate may also vary geographically.

We fit every model separately for each ENSO group. As the group already stratifies the continent, no model includes a regional level. The occurrence model nests random intercepts at country, first-level unit and second-level unit. The duration and monthly case-count models, fitted like the occurrence model to first- and second-level units, nest them at country and first-level unit, with an additional second-level intercept for observations at second-level units. We estimate random effects as mutually independent variance components.

##### S1.3.3 ENSO exposure–response

We evaluate the ENSO exposure at a single lag *L*, selected for each model and ENSO group (Section S1.4.5), and model it as a smooth non-linear function of the index:

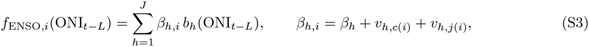

where *b*_1_*, … , b_J_* are the basis functions of a natural cubic spline with *J* degrees of freedom, *β_h_* the population coefficients and *v_h,c_*_(_*_i_*_)_ and *v_h,j_*_(_*_i_*_)_ the country and first-level-unit random slopes of basis column *h*. We write *f*_ENSO_ for the population curve (*v* = 0). The natural spline is linear beyond its boundary knots, which limits the curvature of the fitted curve at extreme ONI values. We chose *J* together with the grouping, from the latent-class models with four to five degrees of freedom (Section S1.4.2); the chosen basis, *J* = 4, is used in all three models and all groups.

The random slopes on every basis column let the shape of the exposure–response curve vary geographically within a group. The boundary knots are the extremes of the ONI values at lag *L* over the observations of the group’s dataset, so the fit itself involves no extrapolation. The simulations read index values beyond this range, chiefly in strong El Niño events, and clamp the index (Section S1.6.11).

##### S1.3.4 Occurrence model

We model outbreak occurrence with a Poisson mixed model of the form (S2) fitted to the occurrence dataset, with the baseline rate as a second offset (Section S1.3.7), 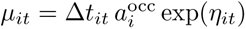, and linear predictor

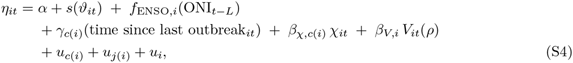

where *ϑ_it_* is the day of the unit’s cholera year on which month *t* starts (0 to 365; Section S1.1.6), *s*(*·*) is a cyclic cubic regression spline with eight knots, giving a smooth seasonal baseline hazard that joins continuously across the cholera-year boundary, *f*_ENSO_*_,i_* is the single-lag exposure–response function (S3) at the group-specific lag *L*, and *u_c_*_(_*_i_*_)_, *u_j_*_(_*_i_*_)_ and *u_i_* are the nested random intercepts at country, first-level unit and second-level unit. The coefficients of the outbreak-history and contagion terms vary by unit through random slopes: *γ_c_*_(_*_i_*_)_ and *β_χ,c_*_(_*_i_*_)_ = *β_χ_* + *v_χ,c_*_(_*_i_*_)_ add a country slope to the population effects, and *β_V,i_* = *β_V_* + *v_V,c_*_(_*_i_*_)_ + *v_V,j_*_(_*_i_*_)_ a country and a first-level-unit slope.

Time since last outbreak at the unit enters as a categorical variable with seven levels: less than one year, one to two years, and so on up to six years or more. Units without an earlier recorded outbreak are assigned to the last level. The categorical form allows a strongly non-linear effect, large in the first year or two and flat thereafter.

The within-unit contagion indicator *χ_it_* equals one when an outbreak episode was ongoing anywhere in the first-level unit *j*(*i*) at any time during month *t −* 1.

We define the neighbour pressure *V_it_*(*ρ*) as the distance-weighted share of other first-level units of the same transmission unit with an ongoing outbreak one month earlier:

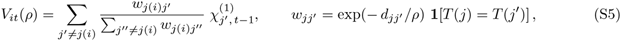

where 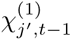 is the ongoing-outbreak indicator of first-level unit *j*^′^ in month *t −* 1, defined as for *χ*, *d_jj_′* is the distance between the centroids of units *j* and *j*^′^, *T* (*·*) is the transmission unit, and *ρ* is a bandwidth that we fix at 500 km. Transmission units are two pre-specified clusters of countries, Central–East and West, within which cholera activity is connected by movement and shared epidemic waves [48] (table S6); they are independent of the ENSO groups, which describe the climate signal rather than the spread of infection. The normalising sum runs over every first-level unit of the transmission unit, including units never affected, so that the covariate is bounded between 0 and 1. As it excludes the focal first-level unit *j*(*i*), it measures pressure from neighbours only and remains distinct from *χ*. Neighbours are coupled across national borders within a transmission unit but not across transmission units, and second-level units take the value of their first-level unit.

Time since last outbreak and the within-unit contagion indicator have country random slopes, and the neighbour pressure and each of the *J* ENSO basis columns have country and first-level-unit random slopes. The seasonal spline does not vary by unit.

##### S1.3.5 Duration model

We model outbreak duration with a Poisson cessation-hazard model of the same form (S2), fitted to the duration dataset, in which the outcome indicates that an ongoing episode ended in a given month. Episodes enter at the first- or second-level unit in which they were recorded.

Episode *k* contributes one interval for each calendar month *t* during which it is active, and we denote this set of months by *A_k_*. Each interval carries the indicator *y_kt_ ∈ {*0, 1*}* that the episode ended in month *t*, the number of days *e_kt_ >* 0 of month *t* during which the episode was active, floored at one day, and the episode’s age *d_kt_*, which serves as the baseline clock.

Writing *ν_kt_*for the cessation hazard, assumed constant within the month, the likelihood contribution of each interval is

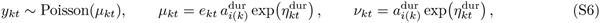

where 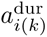 is the empirical baseline cessation rate (see Section S1.3.7) and log *e_kt_* enters as an offset, so that exp(*η*^dur^) is a hazard ratio relative to that baseline. The cessation log-hazard is

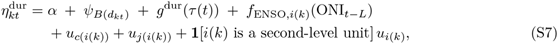

where *B*(*·*) maps episode age *d* to one of six bands (months 1, 2 and 3 each form a band, followed by months 4–5, 6–8 and 9 or more), *ψ*_1_*, … , ψ*_6_ are the band effects with *ψ*_1_ *≡* 0 for identifiability, so that the intercept *α* carries the hazard of the first month, *g*^dur^ is a seasonal term with two harmonics in the calendar month *τ* = *τ* (*t*) of the active month, capturing an annual and a semi-annual cycle,

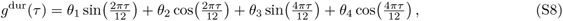

with four fixed effects *θ*_1_*, … , θ*_4_ without random slopes, *f*_ENSO_*_,i_*_(_*_k_*_)_ is the single-lag exposure–response function (S3) with country and first-level-unit random slopes on its *J* basis columns, and *u*_(·)_ are nested random intercepts at country and first-level unit, with a second-level intercept for episodes recorded at a second-level unit. The band effects have their own prior, *N* (0, 1.5) (table S5). As for the occurrence model, this Poisson likelihood is the survival likelihood with right-censoring [45, 46]: censored episodes contribute months in which the cessation indicator is zero and no terminating month. The forward simulator draws cessation events from the monthly probability 1 *−* exp(*−ν_kt_e_kt_*) (Section S1.6.4).

##### S1.3.6 Monthly case-count model

We model the monthly case count *C_kt_* of episode *k* in month *t* with a negative binomial mixed model:

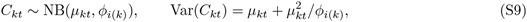

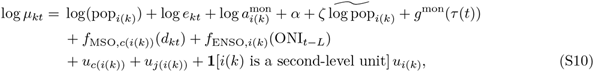

where *e_kt_* is the number of active days as in the duration model, 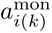 the empirical baseline rate of the episode’s unit, and *ζ* a population elasticity on the log population 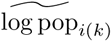, centred within administrative level. Population thus enters both as an offset and through *ζ*, so that case counts need not scale proportionally with population. The seasonal term *g*^mon^ has the two-harmonic form (S8) of *g*^dur^, with its own coefficients. The within-episode epidemic curve *f*_MSO_*_,c_* is a cubic B-spline with four degrees of freedom in the episode’s age *d_kt_* (months since onset), with country random slopes on its four columns, and *f*_ENSO_*_,i_*_(_*_k_*_)_ is the single-lag exposure–response function (S3) with country and first-level-unit random slopes. Random intercepts are nested at country and first-level unit, and observations at second-level units have an additional second-level intercept.

We also model the negative binomial dispersion hierarchically, with country and first-level-unit random intercepts on log *ϕ_i_*_(_*_k_*_)_. This allows overdispersion to vary by geography, as excess variability in reported case counts reflects local surveillance practice as much as local epidemiology.

##### S1.3.7 Empirical baseline-rate anchoring

In all three models the conditional mean is a product of offsets and an exponentiated linear predictor, *μ* = (Π*_n_ O_n_*) exp(*η*). We include in each model an additional offset, the empirical baseline rate *a*, whose denominator equals the product of the model’s other offsets:

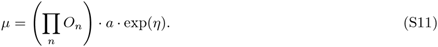

The intercept then measures the log ratio of the fitted rate to the observed baseline rate, a dimensionless quantity of order one, so that exp(*α*) *≈* 1 is the appropriate prior centre in every model. The numerator and denominator of *a* are:

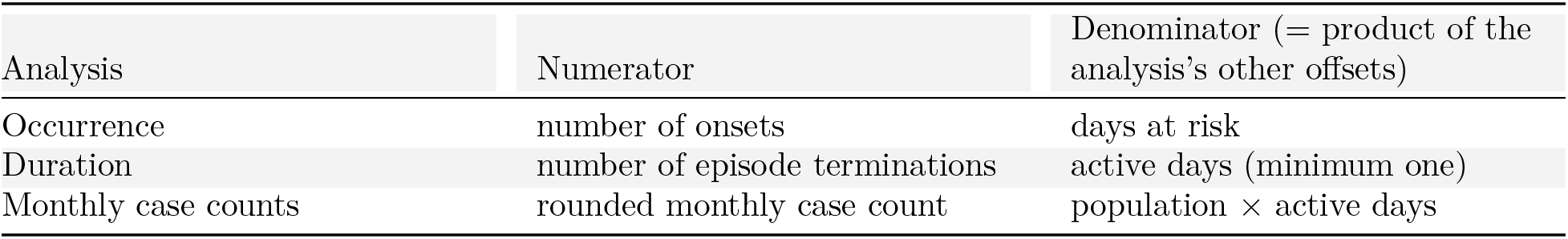

| Analysis | Numerator | Denominator (= product of the analysis’s other offsets) |
|---|---|---|
| Occurrence | number of onsets | days at risk |
| Duration | number of episode terminations | active days (minimum one) |
| Monthly case counts | rounded monthly case count | population × active days |

We estimate baseline rates in cells defined by country and administrative level. For cell *m* with numerator total *Y_m_* and denominator total *E_m_*, the raw estimate is log(*Y_m_/E_m_*). Under *Y_m_ ∼* Poisson(*E_m_a_m_*), the delta-method variance of log(*Y_m_/E_m_*) is 1*/Y_m_*, and we shrink each cell towards its country parent, the rate of its country pooled over administrative levels after adjustment to a common level (below), with the empirical-Bayes weight *w_m_* = *ξ*^2^*/*(*ξ*^2^ + 1*/Y_m_*) = *Y_m_/*(*Y_m_* + 1*/ξ*^2^), where *ξ*^2^ is the between-cell variance in excess of Poisson sampling noise, which we estimate by moments over cells with at least one event as 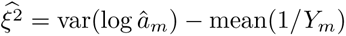, floored at 10^−6^.

Baseline rates also differ systematically by administrative level, as a finer unit has a smaller population and therefore a different event rate per unit time. We estimate this difference between levels within countries (the level contrast), and use it to adjust a cell to a common level before pooling it into a coarser parent and to re-express a borrowed parent rate at the target level. The contrast is the level coefficient of a Poisson regression of the numerator on administrative level and country with the log denominator as offset, in which the country term performs the demeaning. A contrast whose confidence interval includes zero is set to zero.

We resolve the rate of each cell in order of specificity: (i) for a cell with observations, its own shrunken estimate, which for a cell without events is its country parent; (ii) for a cell without observations in a country observed only at other administrative levels, that country’s rate adjusted to the target level; (iii) the ENSO-group rate; and (iv) the continent-wide rate. No cell needed (iii) or (iv).

Of the 60 country-by-level cells at the first and second administrative levels, 51 have observations of their own in the occurrence model, 51 in the monthly model and 46 in the duration model, in which 5 further cells have observations but no termination and take the value of their parent. The remaining 9 cells borrow a level-adjusted country rate. In the duration model the between-cell variance is at its floor, indicating no heterogeneity in cessation rates beyond Poisson noise once country and level are accounted for; the shrinkage weights are then close to zero, and every duration cell takes, in effect, the rate of its country parent. All three models retain a second-level contrast.

We compute one baseline-rate table for the whole continent, used by every group and model and by the simulator.

##### S1.3.8 Priors

#Table S5 lists the priors. The intercept *α* has a normal prior, and every fixed-effect coefficient *β*, including the ENSO basis coefficients, a normal prior centred at zero. The standard deviations of random intercepts and slopes at the country and administrative-unit levels, the smoothing standard deviation of the seasonal spline and the random-effect standard deviation of the log dispersion have half-normal priors. In the single-country group GG (Nigeria), the country-level random effects are kept, so that the simulator draws them like those of the other groups, under a tight *N* ^+^(0, 0.05) prior on their standard deviations, as one country cannot inform a between-country variance.

As the intercept measures a log ratio to the empirical baseline rate, its prior puts 95% probability on fitted rates within roughly a factor of seven of the observed baseline rate. The prior on the other fixed effects (all but the duration band effects, whose prior is wider; table S5) puts 95% probability within roughly a factor of 2.7 per unit change in a covariate on the rate scale. We chose the random-effect standard deviation priors based on prior-predictive simulations, which showed that wider scales imply between-unit rate ratios of implausible magnitude. For the seasonal spline of the occurrence model we used *N* ^+^(0, 0.35) on the smoothing standard deviation, as the heavy-tailed half-*t*_3_ prior with scale 2.5 allows seasonal amplitudes far larger than observed and dominated the residual prior-predictive over-prediction even with the intercept anchored.

##### S1.3.9 Estimation

We fitted all models with Hamiltonian Monte Carlo as implemented in the Stan programming language [49], with four chains of 350 warm-up and 1,000 sampling iterations, giving 4,000 posterior draws per model. We assessed convergence for every model using the potential scale reduction factor *R̂*, bulk and tail effective sample sizes, the number of divergent transitions after warm-up, and the energy-based Bayesian fraction of missing information (GitHub data folder, file D27 convergence, fig. S11).

#### S1.4 Data-informed ENSO grouping and lag selection

Two structural choices are made from the data: which countries share an ENSO exposure–response (the grouping) and at which lag the index enters each model. Both encode the timing and sign of the ENSO teleconnection, which differ across Africa and are not known *a priori* precisely enough to fix per country, so we make both by predictive comparison, in two steps. We first estimate the partition of countries with a latent-class extension of the occurrence model, in which every class has its own ENSO lag and exposure– response curve, choosing the number of classes and the spline basis from a country-block cross-validation comparison and the agreement of the chains (Section S1.4.2). Within this partition, we then select the lag of each model and group by approximate leave-one-out cross-validation over the lag grid, with *L ≥ −*8 (Section S1.4.5). The class lags of the latent-class model only define the partition. As the same occurrence data choose the partition and fit the group models, we report each country’s membership uncertainty beside its effects.

##### S1.4.1 ENSO teleconnection grouping

The influence of ENSO on African rainfall and temperature differs in sign, strength and timing between regions [20]. We therefore fitted every model separately for each of five ENSO groups of the 30 countries with outbreak data (table S7, fig. S6a).

The groups are data-informed: they are the classes of a latent-class model of outbreak occurrence, in which countries that share an ENSO lag and exposure–response form one class (Section S1.4). They are classes of countries, not necessarily contiguous geographic regions. We name each group after the part of the continent in which most of its members lie, and list them from west to east and south: GW (West), GG (Guinean Gulf), GH (Horn), GE (East) and GS (South). GG holds Nigeria alone; most other countries of the Gulf of Guinea coast are in GW. Table S7 lists the members of each group. Released data files label each group by the *a priori* regional group whose seed countries fix its class, which keeps the class labels identifiable (Section S1.4.2; table S7).

##### S1.4.2 Latent-class grouping model

In the latent-class model each country *c* belongs to one of *K* classes *q* = 1*, … , K*, and the ENSO term of a unit-month (*i, t*) of a country in class *q* is

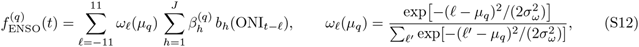

a Gaussian kernel over the lag grid centred at the class lag *µ_q_*, with a fixed width *σ_ω_* = 1.5 months and a uniform prior on *µ_q_* over [*−*11, 11], applied to a natural cubic spline of the index with *J* degrees of freedom. The rest of the linear predictor is that of the occurrence model (S4) without a regional level, and the class coefficients *β*^(^*^q^*^)^ have country and first-level-unit random slopes. Class membership is marginalised at country level: the likelihood of a country is the mixture over classes of the likelihood of all its unit-months, with membership probabilities softmax*_q_*(log *π_q_* + *κA_cq_*), where *A* is the one-hot encoding of an *a priori* partition into East Africa, the Horn of Africa, Southern Africa, West Africa and Nigeria (table S7) and *κ ≥* 0, with a half-normal prior, measures how strongly the data retain it. Classes are identified by a core of seed countries fixed to their *a priori* group (Tanzania, Uganda and Burundi; Ethiopia and Sudan; Malawi, Zimbabwe and Zambia; Niger, Liberia and Togo; Nigeria). The seeds identify the classes: without them the mixture is unchanged by a relabelling of its classes, and in exploratory fits the chains swapped class labels or left classes empty. Fixing a core of countries to each class keeps the labels fixed within a chain, so that chains can be compared and averaged class by class. The core of each of the four regional classes is made of the countries that fell in the class of their *a priori* group in about 85% or more of the chains of exploratory four-class fits seeded with one country per class, which had left some classes with a single member; Uganda and Burundi are fixed with Tanzania, and Nigeria, its own *a priori* group, seeds the fifth class alone. All other countries are free.

We fitted the model over a grid of natural-spline bases with four to five degrees of freedom and two to five classes, with *a priori*partitions for fewer than five classes that merge whole groups of the five-group partition, and compared the fits by country-block approximate leave-one-out cross-validation, one block per country (table S8). Each fit ran eight chains of 400 warm-up and 300 sampling iterations. The latent-class model is multimodal, so we also required the chains of a fit to agree: we retained the three chains whose class lags agree best (the smallest maximum difference between their class lags), and summarised every fit on these chains. A mode is a set of chains whose class lags agree within 1.5 months; the mode sizes of every fit are given in table S8.

We selected the final fit, with four degrees of freedom and *K* = 5 classes, because its three agreeing chains form its dominant mode and its difference in expected log predictive density to the best fit of the comparison, *−*16.4 (standard error 14.2), is about one standard error. The best fit, with five degrees of freedom and five classes, has no mode of three chains. Each country is assigned to its class of highest mean posterior membership probability over the agreeing chains, and we write *p*_max_ for that probability.

Fig. S6c shows how the classes change as *K* rises from 2 to 4 at the selected basis, and panel b the final fit with *K* = 5. We describe each class by the final groups of its members: a class written GW + GG holds the countries of both groups, and “part of” gives the share of a group’s countries that the class holds. The most probable classes are, at *K* = 2: GW + GG + GH, GE + GS; at *K* = 3: part of GW (10 of 15) + GH + GS, part of GW (5 of 15) + GG, GE; at *K* = 4: part of GW (10 of 15) + GS, part of GW (5 of 15) + GG, GH, GE; at *K* = 5: GW, GG, GH, GE, GS. The final partition keeps part of the regional structure of the *a priori* partition, and two of the 30 countries are assigned to a class other than the one their *a priori* group seeds (table S7). The countries of the WHO African region without outbreak data are not assigned to any group.

One of the 30 countries, Ghana, has *p*_max_ *<* 0.6, and the class sizes summed over the membership probabilities differ from the hard counts (fig. S6b). The seed countries have *p*_max_ = 1 by construction. We report *p*_max_ beside every group and country effect (Figs. 1 and 2; table S9; fig. S6b; data S1, files `T3a`_`effects_country_occurrence, T3b_effects_country_duration` and `T3c_effects_country_monthly`), the stability of the partition under sampling from the membership probabilities, and its sensitivity to the number of classes, the exposure basis and the chains used: adjusted Rand index and countries that change class after an optimal matching of class labels, with the country-block difference in expected log predictive density to the final fit (GitHub data folder, files `D5_membership_stability` and `D5_membership_sensitivity`; fig. S6d).

##### S1.4.3 Rainy seasons of the teleconnection groups

Panel c of Fig. 1 in the main text outlines the main rainy season or seasons of each teleconnection group. For each first-level administrative unit we computed the monthly rainfall climatology, the mean CHIRPS rainfall [50] of each calendar month over 1990–2019, from the admin-level climate dataset of the VIMC climate project [51] (for units without their own series, the mean over their second-level units). A calendar month is rainy in a unit when its climatological rainfall exceeds the unit’s average month, one twelfth of its annual total [52]; units receiving less than 100 mm a year have no rainy season. A month belongs to a group’s main season when at least half of the group’s 2025 population, from the population estimates described in Section S1.1.5, lives in units in which that month is rainy. Weighting by population rather than counting units prevents sparsely populated arid units from shaping a group’s season. Each run of consecutive such months, allowed to cross the year end, is outlined as one season, so a group with two rainy seasons shows two outlines. Months close to the 50% threshold could fall on either side of it under a different population year or climatology period.

##### S1.4.4 Seasonality of outbreaks, cessation and cases

We compared the rainy seasons of Section S1.4.3 with the monthly pattern of each outcome, by ENSO group and calendar month (the fitted seasonal patterns are shown in panel c of Fig. 1 in the main text). For occurrence we computed the number of outbreak onsets per 1,000 unit-months at risk in the occurrence dataset, by the calendar month of the onset. For cessation we computed the number of episodes ending per 100 active episode-months in the duration dataset. For case counts we took the median and interquartile range of suspected cases per active episode-month in the monthly dataset, leaving out months with a reporting gap. All three are computed on the units the models are fitted to (Section S1.1.8). Onset and cessation rates carry exact Poisson 95% intervals. A month with fewer than five events is not used to locate a peak. Observed onsets peak in a rainy-season month in all five groups, cessation in one of the five groups and cases per active month in three of the five groups.

##### S1.4.5 ENSO lag selection

Once the country partition was established, for each model and ENSO group, we fitted the single-lag model (S3) separately at every candidate lag *L ∈ {−*11*, … ,* 11*}* months, and compared the fits by approximate leave-one-out cross-validation with Pareto-smoothed importance sampling [53]. For each lag we computed the expected log pointwise predictive density relative to the best lag, together with the standard error of the paired difference.

A negative lag is admissible because the index after the outcome month is a function of the developing ENSO event, which can precede its own peak by several months.

We select for each model and group the lag with the highest expected log predictive density among the lags allowed by a floor, *L ≥ −*8. The floor keeps the index a model reads within the forecast’s reach: the firm forecast horizon of a group is the last month of the ENSO forecast plus its smallest lag (Section S1.6.12), so a lag of *−*9 to *−*11 months would end it within a few months of the launch. The duration and monthly case-count lags are selected independently of the occurrence lag. Selected lags are given in table S9, and full rankings, with the Pareto *k* diagnostics of every lag, in the GitHub data folder, file `D7_lag_loo_ranking` and fig. S7. We call two lags distinguishable only when their difference exceeds two standard errors of the paired difference; the rankings give, for every model and group, the number of lags within two standard errors of the best, and in several of them the selected lag is a point choice among nearly equivalent lags.

The floor binds in three of the 15 combinations of model and group (duration model of GH, best lag *−*9; monthly case-count model of GH, best lag *−*10; monthly case-count model of GG, best lag *−*11). Table S9 reports for every model and group the floor, whether the best lag lies below it and the paired difference in expected log predictive density that this costs, the second-best lag and the best non-negative lag, each with its difference to the best lag. Fig. S7 shades the lags below the floor.

#### S1.5 Derived quantities

##### S1.5.1 Exposure–response curves

We reconstruct posterior exposure–response curves draw by draw. For each posterior draw *s*, we combine the fitted basis coefficients 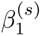*, … ,* 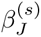 with the spline basis evaluated on a grid of ONI values, and express the result as an incidence rate ratio relative to the reference ONI_0_ = 0, the centre of ENSO-neutral conditions:

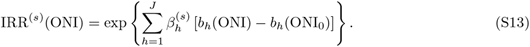

We summarise curves by the pointwise median and the equal-tailed 50% and 95% credible intervals of IRR^(^*^s^*^)^(ONI) across draws. Group-level curves use the fixed-effect coefficients, and country- and first-level- unit curves add the corresponding random slopes to 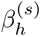 (figs. S12 to S14). Posterior summaries of the other model terms are shown in figs. S8 to S10.

##### S1.5.2 Population living in units with an ENSO effect

For an ENSO state ONI_1_ *∈ {−*1.5*, −*1, 1, 1.5, 2*}*, a first-level unit has an effect of magnitude *m* in a model when its unit-level rate ratio (S13) at ONI_1_ lies above 1 + *m* or below 1*/*(1 + *m*). The effect is adverse when it means more cholera: onset and case rate ratios above 1 + *m*, and a cessation hazard ratio below 1*/*(1 + *m*), hence longer outbreaks. It is protective in the opposite direction, and we also count effects in either direction. We evaluate magnitudes *m ∈ {*0, 0.10, 0.25, 0.50*}* and posterior probability cut-offs *p ∈ {*0.5, 0.8, 0.95*}*, for each of the three models alone and for any of them.

A unit is counted at (*m, p*) when the model, taken on its own, gives a posterior probability of at least *p* to an effect of magnitude *m* in the chosen direction; for “any model”, when at least one of the three models does. The headline count is the population of the units with an adverse effect in any of the three models at *m* = 0.25 and *p* = 0.95. It is complemented by the expected population with an effect, the posterior mean and 95% interval over draws of the population of the units whose drawn rate ratio lies beyond the same bound, and by the counts at every other magnitude, cut-off and direction (data S1, files D10 enso exposure exceedance and D12 enso exposure by country). Populations are the 2025 estimates of each unit of the dataset (Section S1.1.5).

#### S1.6 Simulation

We use the fitted models to simulate outbreak histories forward in time under factual and counterfactual ENSO trajectories. This section describes the simulator, the particle filter that reconstructs the historical state, and the hindcast, forecast and attribution designs.

##### S1.6.1 Notation

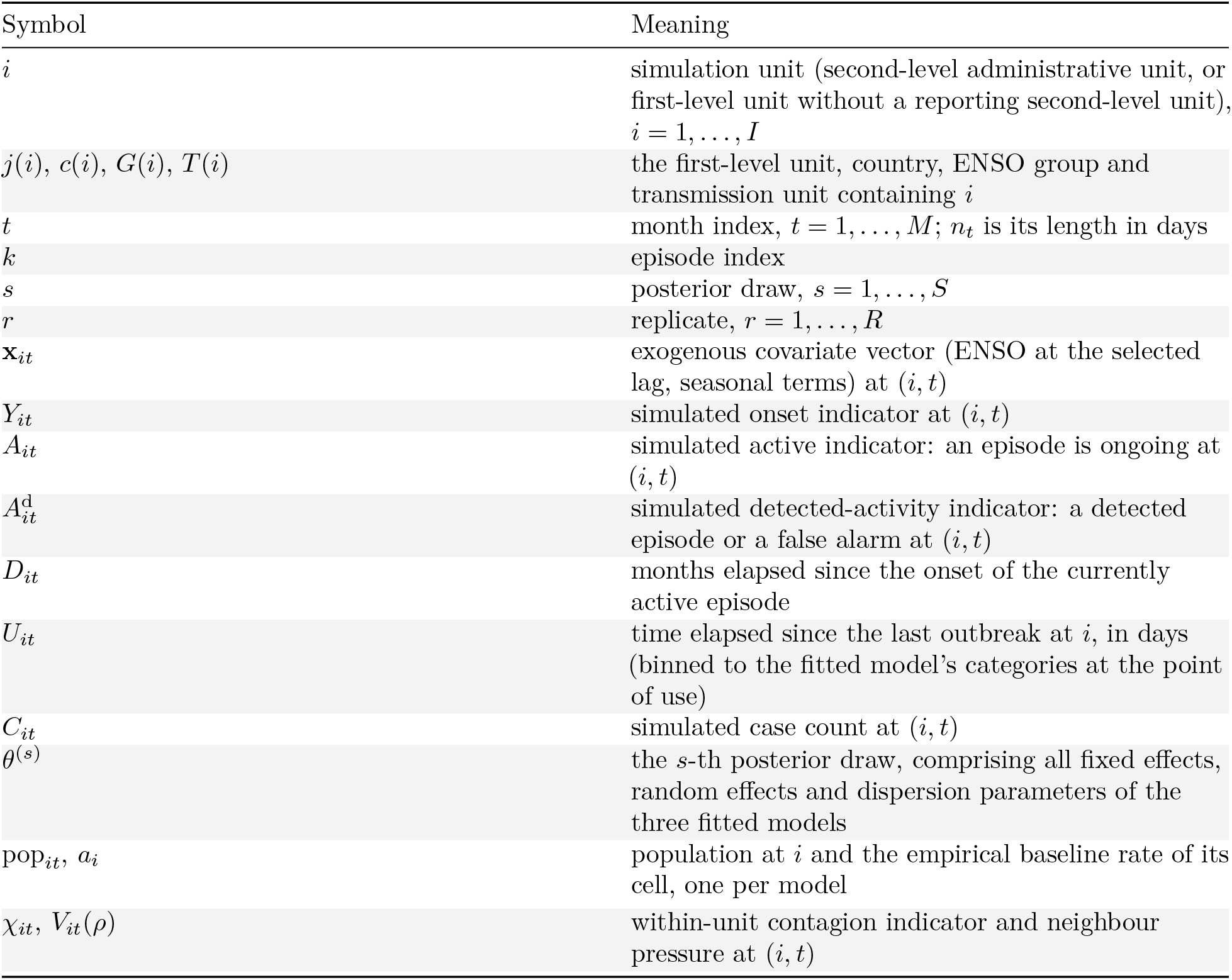

| Symbol | Meaning |
|---|---|
| *i* | simulation unit (second-level administrative unit, or first-level unit without a reporting second-level unit), *i* = 1, ..., *I* |
| *j*(*i*), *c*(*i*), *G*(*i*), *T*(*i*) | the first-level unit, country, ENSO group and transmission unit containing *i* |
| *t* | month index, *t* = 1, ..., *M*; *n*<sub>*t*</sub> is its length in days |
| *k* | episode index |
| *s* | posterior draw, *s* = 1, ..., *S* |
| *r* | replicate, *r* = 1, ..., *R* |
| **x**<sub>*it*</sub> | exogenous covariate vector (ENSO at the selected lag, seasonal terms) at (*i*, *t*) |
| *Y*<sub>*it*</sub> | simulated onset indicator at (*i*, *t*) |
| *A*<sub>*it*</sub> | simulated active indicator: an episode is ongoing at (*i*, *t*) |
| *A*<sup>d</sup><sub>*it*</sub> | simulated detected-activity indicator: a detected episode or a false alarm at (*i*, *t*) |
| *D*<sub>*it*</sub> | months elapsed since the onset of the currently active episode |
| *U*<sub>*it*</sub> | time elapsed since the last outbreak at *i*, in days (binned to the fitted model's categories at the point of use) |
| *C*<sub>*it*</sub> | simulated case count at (*i*, *t*) |
| *θ*<sup>(*s*)</sup> | the *s*-th posterior draw, comprising all fixed effects, random effects and dispersion parameters of the three fitted models |
| pop<sub>*it*</sub>, *a*<sub>*i*</sub> | population at *i* and the empirical baseline rate of its cell, one per model |
| *χ*<sub>*it*</sub>, *V*<sub>*it*</sub>(*ρ*) | within-unit contagion indicator and neighbour pressure at (*i*, *t*) |

##### S1.6.2 State vector and its dynamics

The simulator is a discrete-time Markov chain with state at the end of month *t*

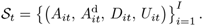

All other quantities needed by the models at month *t* + 1 are deterministic functions of *S_t_* and of the exogenous covariates. As the models were fitted to recorded episodes, the contagion covariates and the time since last outbreak are built from detected rather than latent activity. Each simulated episode is detected at onset with probability 1 *− δ*_fn_ and keeps this status while it lasts, and an inactive unit records a false alarm with monthly probability *δ*_fp_, with *δ*_fn_ and *δ*_fp_ as in the measurement model of Section S1.6.8; 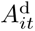 = 1 for a detected episode or a false alarm. We compute the two contagion covariates from the detected indicators as

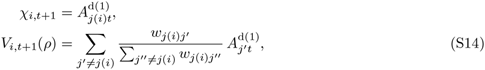

where 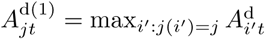 is the detected state of first-level unit *j* and *w* is the distance kernel of (S5), restricted to the transmission unit. As in the fitted model, *χ* includes the unit’s own activity and *V* excludes it.

In the fitted data, the neighbour covariate counts recorded activity only, so a neighbour without surveillance in the previous month counts as inactive. We measured this coverage without changing the covariate. A unit was under surveillance in a month if one of its location periods had a reporting window covering it (including first-level series), the month was a national-zero month, or the unit had a recorded episode then. For each unit-month we computed the kernel-weighted share of its neighbours under surveillance in the previous month, over the same kernel and transmission-unit restriction as the covariate. In the simulations every unit is simulated, so the implicit coverage there is one, and the simulator applies the contagion effect at a neighbour activity that the fit may not have observed where coverage was low. Coverage by country, group and year is given in GitHub data folder, file `D33_contagion_coverage`.

Time since last outbreak advances at each month without a detected onset, and resets at a detected onset; false alarms feed the contagion covariates but do not reset it:

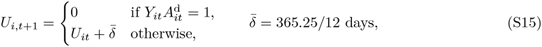

where we advance the counter by the mean month length 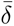 rather than by the calendar length *n_t_*, as it only feeds a coarse categorical coding. A unit with no recorded onset is coded in the oldest category (six years or more), as in the fitted model. Months since onset advance while an episode is active and are undefined otherwise.

The within-year clock of the seasonal baseline hazard resets at each location’s trough month, but the state variables do not: recent outbreak history and ongoing outbreaks carry over across the cholera-year boundary in calendar time.

As the distance kernel is zero between transmission units (Section S1.3.4), no state variable couples them: transmission units evolve independently given a posterior draw and the covariates, and are simulated in parallel. Each holds 15 modelled countries of several ENSO groups (table S6). Each country is simulated with the models, selected lags and posterior draws of its own ENSO group, while the contagion terms couple it to its neighbours.

##### S1.6.3 Occurrence step

At each month, we evaluate the occurrence model for every unit not currently experiencing an episode, given the current state and the covariates:

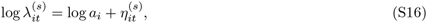

where 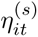 is given by (S4) evaluated at draw *s*. Following (S1), the probability of an onset during the month is

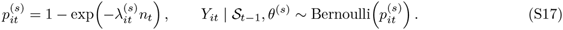

The simulator follows the fitted risk set, from which a unit leaves after an onset until the next cholera year (Section S1.1.7): the hazard (S16) is that of the first onset of a cholera year, so we draw an onset only for a unit that is inactive and has had no detected onset yet in its current cholera year. With 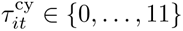 the month of the unit’s cholera year, counted from its first month, this holds when 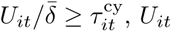 being read before its update (S15), or when the unit has no detected onset at all. A unit whose episode ends within the cholera year is therefore not at risk again before the next one, and simulated detected onsets, like the fitted data and the observations the hindcasts are scored against, number at most one per unit and cholera year. An undetected episode does not close the risk set, as an unrecorded episode does not in the fitted data.

##### S1.6.4 Duration step

For an episode *k* active at (*i, t*) with *D_it_* months since onset, we evaluate the cessation hazard from the duration model at draw *s*,

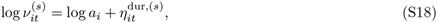

where 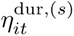 is given by (S7) with its band term evaluated at the simulated episode age *D_it_* and its seasonal term at the calendar month of *t*. The episode ends during the month with probability

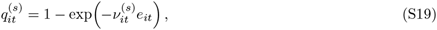

where *e_it_* is the number of days of month *t* during which the episode is active. As the simulator does not resolve days within a month, it sets *e_it_* to the length of the month, halved in the onset month (the expected active fraction under a uniformly distributed day of onset) and, in the case-count step (S21), also halved in the month the episode ends, whose cessation is drawn before the case count, as in the fitted dataset, where an episode’s first and last months are partial. The cessation probability itself carries only the onset correction, as whether the month is the last is what it draws. Episode durations therefore follow a discrete-time survival distribution, in which the probability that an episode starting in month *t*_0_ is still active after *d* months is

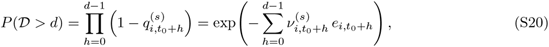

and the expected duration in months is 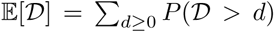. As the cessation hazard combines a categorical function of months since onset with the calendar-month harmonics (see Section S1.3.5), (S20) reproduces the non-geometric empirical distribution of durations, in which few episodes end in their first month and the monthly probability of ending levels off from the third, and the expected duration depends on the month of onset.

##### S1.6.5 Case-count step

For each month in which an episode is active, we draw a case count from the monthly model,

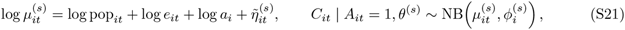

where 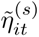 collects the non-offset terms of (S10) (intercept, population elasticity, seasonality, within-episode curve, ENSO and random effects) evaluated at draw *s*, with the within-episode curve *f*_MSO_ evaluated at the simulated *D_it_*. The negative binomial is parametrised such that Var(*C*) = *µ* + *µ*^2^*/ϕ*. The mean and the drawn count are both capped at 15% of the unit’s population per month.

In the simulator and the historical reconstruction, the population pop*_it_* of a unit is resolved for every simulated month at that month’s cholera year, the calendar year of the month *m_i_ −* 1 months before *t*, with *m_i_* the unit’s trough month (see Section S1.1.6). This is the convention under which the monthly case-count model was fitted. Populations come from the unit population table (2015–2030), and years outside it are rescaled by the national totals of the UN World Population Prospects 2024. The population offset, the population elasticity term and the incidence cap therefore follow population growth over the simulated period. Per-capita rates (annualised incidence and attributable cases per 100,000) divide by the month-weighted mean population over the months each estimate covers, and counts of people living in affected areas use the 2025 population.

##### S1.6.6 Algorithm

We simulate each replicate *r* independently for each transmission unit, such that replicate-unit pairs can be distributed across processors. Each replicate uses a single posterior draw *s_r_* for all three models of every ENSO group in the transmission unit, so that occurrence, duration and case-count parameters come from a coherent posterior state. The initial state *S*_0_ is a terminal state of the particle filter (see Section S1.6.8). At each month *t* = 1*, … , M* , we compute the covariates **x***_it_* and the contagion covariates from *S_t_*_−1_ using (S14). We then draw onsets with (S17) for units in the risk set (Section S1.6.3), cessation for active units with (S19), and case counts for units active during the month with (S21), and update *A*, *A*^d^, *D* and *U* to obtain *S_t_*. Each replicate returns its event table, with the onset month, duration and total cases of every episode, and its unit-by-month state array.

Detection only feeds the covariates. Every reported simulated quantity, in the reconstruction, the hindcasts, the forecasts and the attribution, counts latent onsets and cases, detected or not.

Random number streams are seeded by replicate, so results do not depend on parallelisation. With 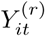 and 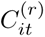 the onsets and cases of replicate *r* and *R* the number of replicates (Section S1.6.7), the marginal onset probability at (*i, t*) is 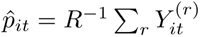 the expected number of outbreaks over a window 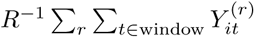, and the expected total number of cases 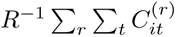, with credible intervals given by empirical quantiles across replicates.

##### S1.6.7 Uncertainty decomposition

For a scalar summary *Z* of a simulated trajectory, for instance the total number of outbreaks in a country over a horizon, the total variance decomposes as

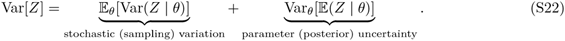

Replicates at a fixed posterior draw resolve the first term, and replicates across posterior draws the second. The forward simulations nest replicates within draws in a fixed cycle: replicate *r* runs posterior draw *s_r_* = ((*r −* 1) mod *S*) + 1 of a fixed set of *S* draws, so that every draw is run by *R/S* replicates. The forecast, the counterfactual, the attribution factual and the past-event arms run *R* = 10,000 replicates per transmission unit and arm, *S* = 100 draws of *R/S* = 100 replicates each (GitHub data folder, file `D34_run_design`), and the factual and counterfactual arms of a replicate share its draw. The hindcasts run 5,000 replicates per run in the same cycle, 50 replicates per draw. Intervals over replicates reflect the sum (S22); the intervals of the attributable fraction separate the second term (Section S1.6.14).

##### S1.6.8 Particle filtering for historical state reconstruction

Forecasts and counterfactuals require a starting state consistent with the observed record. We therefore reconstruct the latent state of the simulator over the observed record, from January 2011 to August 2026, by sequential Monte Carlo before any forward simulation, with the sub-national database as observation to December 2024 and the WHO monthly national series from January 2023 to August 2026. The reconstruction ends in the last month with a published ONI season, so that the forecasts start from the latest observed ENSO state. The ENSO covariates of the reconstruction are the observed ONI throughout; for models with a negative lag, the months after August 2026 are taken from the calibrated ENSO forecast described below.

We write *X_t_* = (*X*_1_*_t_, … , X_It_*) for the latent state of all units at month *t*, where *X_it_* comprises the active and detected-activity indicators, the months since onset, the time since last outbreak and the current case-count mean. The transition density *f* (*X_t_ | X_t_*_−1_; *θ*^(^*^s^*^)^) is the one-step simulator described above, evaluated at a fixed posterior draw. The observations at (*i, t*) are the reported onset indicator *Ỹ_it_* and, where available, the reported case count *C̃_it_*.

As reporting is imperfect, we allow for false negatives, onsets that are not recorded, with probability *δ*_fn_, and false positives, recorded onsets without a latent onset, with probability *δ*_fp_. The onset term of the measurement model is

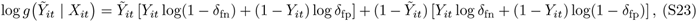

where *Y_it_* is the latent onset indicator. The term is evaluated in the unit-months of the occurrence dataset; that dataset, with the months removed inside running episodes and its national-zero months (Section S1.1.8), sets the risk set of the filter.

The case term is evaluated in every unit-month with a recorded count, including months after an onset, with zeros recorded in at-risk months without a case report. Writing *µ_it_* and *ϕ_i_* for the mean and dispersion of the monthly model, it is

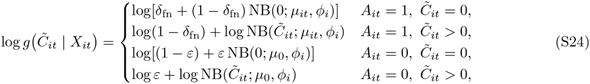

where *A_it_* is the latent active indicator. In an inactive unit a count is reported with probability *ε*, drawn from a negative binomial with a floor mean *µ*_0_ = 0.1, so that the term is a probability distribution over the reported count in both states. The first case penalises latent episodes that continue through months without recorded cases, and the fourth particles whose episode has ended while cases are still reported. We set *ε* = 1, so that the inactive term is the floor negative binomial at every count; *ε* cannot be estimated, as every at-risk month outside the recorded episodes is recorded as zero. Missing observations do not contribute to the likelihood. We set *δ*_fn_ = 0.10 and *δ*_fp_ = 0.001, reflecting the assumption that under-ascertainment is possible and that spurious outbreak reports are rare.

The WHO monthly national series enters as a third, country-level term. Writing *W_ct_* for the national count of country *c* in month *t* and 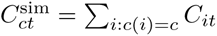 for the particle’s simulated cases summed over the modelled units of the country, the term is

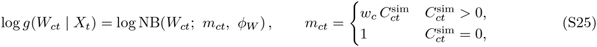

where *w_c_* is a country multiplier and *ϕ_W_* = 5 is a reporting overdispersion distinct from the dispersion of the monthly model, as the process variation is already in *C*^sim^. With *φ_c_* the share of the national population living in the modelled units of the country, the multiplier is by default *w_c_* = 1*/φ_c_*, which scales the simulated count to the national total. Where WHO and the database both observe a country over enough months in 2023–2024 (at least 100 database cases over at least 6 overlap months), we estimate its own scale instead: with *W_c_* the WHO total and DB*_c_* the database total over the simulated units in those months, *ς_c_* = *W_c_φ_c_/*DB*_c_*, clamped to [0.1, 10], and *w_c_* = *ς_c_/φ_c_*, which equals *W_c_/*DB*_c_* unless clamped. This identified 2 of 30 countries; the others keep the fixed factor 1*/φ_c_* (GitHub data folder, file `D30_who_scale`). To avoid scoring a country-month twice, the WHO count is not scored in country-months the database observes. The floor mean of one case gives a particle without simulated activity in the country a finite, graded penalty when cases are reported. This term is evaluated from January 2023; from January 2025, after the end of the sub-national database, it is the only observation, and countries without WHO data run unconstrained from then on. Scoring the simulated count rather than its expectation lets the filter also condition on the sampling variation of the case-count model.

We use the block particle filter [54, 55] implemented in the spatPomp package [30], with one block per country, so that the national term enters its block exactly once, one independent filter run per transmission unit, 30,000 particles, 100 replicates and a 24-month burn-in (January 2009 to December 2010) before the filtering window.

Smoothing replicate *j* runs the filter at draw *j* of the 100 posterior draws in every transmission unit, and returns one state path over the whole record. At the last month, the filter samples one particle per block and traces its ancestry back in time, so that the path is a draw from the smoothing distribution, stitched across the countries of the transmission unit, and its state in any month is conditioned on the whole record, later months included. We call these paths the historical reconstruction. Their last states are the initial states *S*_0_ of the forecasts; the hindcasts and the past-event arms start from an earlier month of the same paths. Forward replicate *r* starts from the state of smoothing replicate *s_r_* (Section S1.6.7), one smoothing replicate per posterior draw, and runs the draw *s_r_* under which that state was reconstructed, so that state and parameters are a joint posterior draw. The pairing depends only on the replicate number, and is the same in every transmission unit and arm.

##### S1.6.9 Validation statistics

We compare simulated and observed outbreak histories over their common period, restricted to unit-months with documented surveillance coverage. The same scores serve the hindcasts (Section S1.6.10).

We assess the calibration of the binary monthly onset outcome with the randomised probability integral transform for discrete outcomes [56], which is uniform on (0, 1) when the predictive distribution is calibrated. A U-shaped histogram indicates under-dispersed predictions, a hump-shaped one over-dispersed predictions, and a sloped one a systematic bias.

We measure predictive accuracy for onset counts and case totals with the continuous ranked probability score (CRPS) [57], estimated from the simulated ensemble with the fair estimator, which does not favour ensembles of a particular size [58]. Lower scores are better, and the score rewards sharpness subject to calibration.

We also perform four structural checks comparing simulated and observed month-of-year onset frequencies across the modelled units, the distribution of episode durations, the fraction of unit-months in the active state, and the probability of an onset given an ongoing outbreak in the previous month in the same first-level unit, in another first-level unit of the same country, or in another country of the same transmission unit (fig. S15). The reconstruction of each country is shown in figs. S16 and S17.

##### S1.6.10 Hindcast skill

We assess retrospective forecast skill with hindcasts. The hindcast years lie inside the fitting period, and the grouping, the lags and the parameters were estimated with them, whereas the reference forecasts leave the scored season out (below). Each hindcast also starts from the historical reconstruction, which has seen the observations of the season it scores (Section S1.6.8), and it reads the observed ENSO index over the scored season rather than a forecast available at launch. The scores are therefore in-sample upper bounds on the skill of the model given perfect knowledge of ENSO; they check consistency and do not establish the skill of a forecast issued in real time (Section S2.1).

Hindcasts follow the cholera season of each country, from trough to trough. A country’s trough is the calendar month with the lowest onset rate (onsets per at-risk unit-month, three-month circular smoothing) in the occurrence dataset over 2010–2024, where the country has at least 30 onsets and the trough is stable (within one month in at least 80% of 500 year-bootstrap resamples), and otherwise the trough month shared by most of its units in the cholera-season calendar (Section S1.1.6). This trough, estimated from the onsets themselves, can differ from the calendar trough that sets the cholera years of the models and the past-event windows. The season window runs from the month after the trough to the next trough month inclusive, one month later than a cholera year that starts at the same trough. Its start is floored at May, after the spring predictability barrier of ENSO, and the months cut by the floor and their share of onsets are reported (GitHub data folder, file `D3_season_ends`). Each country is scored to the end of its season window, extended by the smallest of the three selected lags of its group when that lag is positive; the past-event windows are extended by the largest of the three instead (Section S1.6.16).

For each cholera year from 2012 to 2022, we run one hindcast per transmission unit and launch month. These are the first and last cholera years that fit entirely inside the January 2010 to December 2024 data window with their margins: the launch state needs the reconstructed months and the lagged ONI before it, and the season window, extended by the lag, must close before the data end. In each run, the countries whose season starts in that month are launched together from the state of the preceding month in the historical reconstruction, the whole transmission unit is simulated, and only those countries are scored in that run. The observed ONI is used both over the window and in the months before launch that enter its initial state.

We score occurrence on the number of onsets per unit over each season window, in units and months at risk. Its reference is a climatology of the unit itself: a binomial distribution over the unit’s at-risk months in the window, with the unit’s own onset rate over its at-risk months outside the scored season, so that the scored season does not enter its own reference; its CRPS is computed exactly. We score case counts per month and per season total on the at-risk units, against a pooled monthly climatology: for each scored month, the cases of the same calendar month and its two neighbours in the other cholera years, leaving out the scored season and every season overlapping it, scaled to the population of the scored year. The model ensemble is scored with the fair CRPS (Section S1.6.9) on raw counts and on log(1 + *y*), and with a threshold-weighted CRPS that counts errors above the 50-case outbreak threshold. Skill is CRPSS = 1 *−* ΣCRPS*/* ΣCRPS_ref_, with both sums over the same units, months and seasons, pooled by ENSO group, for Africa and by country. Leads count from each country’s own launch and are grouped into early (0–3 months), peak-season (4–8; labelled “peak” in the figures, not to be confused with the peak of an ENSO event) and late (9 or more) classes. Intervals come from 1,000 bootstrap resamples of whole cholera years (figs. S18 and S19). Fig. S19a shows the skill against the two climatologies by ENSO group and lead class, for onsets and for monthly case counts on both scales. The hindcasts tend to over-predict case totals: the observed total of a group window (one ENSO group and launch year) falls below the 95% prediction interval in 11 of the 55 group windows and above it in two (fig. S19b).

##### S1.6.11 Forecast design

Forecasts start in September 2026 from the filtered state at the end of the observed record (August 2026), with the replicates and draws of the forward arms (Section S1.6.7). This is the forecast shown in the figures; the attribution statistics use a second factual arm launched with the counterfactual (Section S1.6.13).

Over the forecast horizon, we draw the ENSO input from the 40 member trajectories of the latest NCEP Climate Forecast System version 2 (CFSv2) monthly Niño 3.4 forecast issued by the NOAA Climate Prediction Center (CPC) [59], here the forecast issued in October 2026 with target months to July 2027, assigning one ensemble member to each replicate by sampling with replacement.

Raw CFSv2 Niño 3.4 forecasts are biased, and in strong El Niño events they overshoot the observed amplitude. We therefore calibrate them with the probability-distribution and spread correction CPC applies operationally to CFSv2 Niño 3.4 [60], which we rebuild from the CFSv2 hindcasts because CPC publishes the corrected forecast only as a figure. The CFSv2 hindcasts that train the correction are taken from the North American Multi-Model Ensemble archive [61], averaged over the Niño 3.4 box: 360 start months from 1991 to 2020, leads of 0 to 9 months and 24 to 28 members per start; the real-time forecast is the 40-member CPC forecast above. As in CPC’s procedure, the 1991–2020 sample pools hindcasts initialised before and after the 1999 change in the CFSv2 ocean initial conditions [62, 63]. For initialisation month *o* and lead *ℓ*_f_ (written *ℓ* in the subscripts below), let *F_oℓ_* denote the empirical distribution function of all member anomalies of these forecasts (each relative to its own start-month and lead mean) and *G_oℓ_* that of the observed Niño 3.4 anomalies in the corresponding target months. A member anomaly *x_j_* of the real-time forecast is mapped to

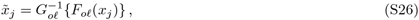

with values beyond the range of the reference sample held at its extremes. The observations are the monthly ERSST Niño 3.4 anomalies whose three-month running mean is the ONI the statistical models were fitted on, so the corrected forecast is on the scale of the fitted ENSO exposure–response. The ensemble spread is then set to the error of the corrected ensemble mean in the same 1991–2020 sample, 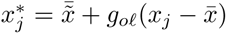 with 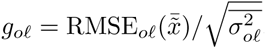, where bars denote ensemble means, RMSE*_oℓ_* is the root mean squared error of the mapped ensemble mean against observations and 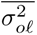 the mean raw ensemble variance. We scale the raw member deviations rather than the mapped ones because, in an event outside the range of the reference sample, the range limit of (S26) maps all members to the same value and would remove the spread entirely; CPC avoids this with a larger real-time ensemble. Only the calibrated ensemble mean is therefore held within the range of the reference sample; the members spread around it without a limit. Mapped instead to OISST v2.1 observations [64], this procedure reproduces CPC’s own corrected ensemble means for the January to June 2026 forecasts to within 0.05–0.13 °C at leads of 0 to 6 months, where CPC’s correction itself moves the forecast by 0.2–0.7 °C. For the forecast issued on 8 October 2026, the ERSST-calibrated seasonal means are within 0.08 °C (root mean squared difference) of CPC’s corrected forecast from October–December 2026 to April–June 2027, whereas the raw forecast exceeds it by up to 0.8 °C.

We join the calibrated members to the observed record. Months already observed take the monthly ERSST value; the CFSv2 analysis months that precede the first forecast month are used only where the monthly ERSST record has not yet been released, after shifting them so that the three-month mean of the latest published ONI season is reproduced. The ONI of each trajectory is computed as a three-month running mean, using the published ONI wherever available.

In the forward simulations, the ONI entering a model is clamped to an interval around its boundary knots. With lo and hi the lower and upper boundary knots of the model’s ENSO basis (the observed ONI range of its group) and *ɛ* the clamp margin, the ONI is held within [lo *− ϵ |*lo*|,* hi + *ϵ |*hi*|*], with *ϵ* = 0.25 in the forecast, counterfactual, hindcast and past-event arms and in the analytic cross-check. Within this interval the natural spline extrapolates linearly beyond the knots; outside it the effect is held at its value at the interval bound. The calibrated forecast of the 2026–27 El Niño reaches or exceeds the upper knot of several fitted curves, where an unbounded extrapolation would be unsupported by the data.

ENSO phases follow one rule set. The NOAA CPC episode rule (ONI at or beyond *±*0.5 °C for at least five consecutive overlapping three-month seasons) is used for the ENSO-state shading of the figures and for the summary of the reconstruction by phase. The cholera-year state of an outbreak and the classification of analogue winters use a threshold phase at *±*0.5 °C without a persistence requirement. The launch of a past event requires the ONI to stay beyond *±*0.5 °C from the month after the launch to the event’s first peak (Section S1.6.16).

##### S1.6.12 Analogue continuation of the ENSO forecast

Beyond the last month whose ONI is a full three-month CFSv2 mean (the month before the last CFSv2 target month), each replicate’s trajectory is continued with an analogue drawn from unforced climate-model simulations. We built a library from the Niño 3.4 series of the pre-industrial control runs published with Lenssen et al. [65] (CMIP6 and earlier NMME and CMIP5-generation models), each rescaled to the standard deviation of the observed ONI over 1991–2020, and characterised every month of every run by its calendar month, its ONI, its three-month tendency and its ONI six months earlier. For a replicate whose CFSv2 trajectory ends in state (*v,* Δ*v, v*_−6_), we took the library months of the same calendar month, ranked them by the standardised Euclidean distance over these three features, keeping at most one start per two years within a run, and drew one of the 30 nearest. The continuation follows the analogue’s subsequent months, with the offset between the replicate’s last value and the analogue’s start fading with a three-month time constant. In a validation on 468 observed seams between 1982 and 2020, this continuation scored better (fair CRPS 0.22, 0.40 and 0.43 at leads of 1–6, 7–12 and 13–24 months) than persistence with autoregressive decay (0.44, 0.75, 0.80), climatology (0.47, 0.47, 0.46) and a neutral continuation (0.67, 0.66, 0.65). One run (GFDL CM2.1-NMME) was excluded from the library because it is the longest run, supplying a third of the starts, and has the strongest cold bias after strong El Niño events; its exclusion improved the score after such events from 0.42 to 0.38 at leads of 7–12 months. The library therefore holds 10 runs, and the same runs supply the counterfactual analogues (Section S1.6.13).

A forward simulation covers every month from its launch to the end of its horizon, both included, and starts from the reconstructed state of the month before the launch. As each model uses ENSO at its selected lag, a group’s firm horizon is the last month whose ONI is a full three-month CFSv2 mean plus the smallest lag selected for that group across the three models; with a negative lag it falls before that month, and a group whose firm horizon equals the launch month keeps one firm month (table S9, fig. S21, GitHub data folder, file `D6_firm_horizons`). Months past a group’s firm horizon read the analogue continuation of ENSO. A transmission unit mixes ENSO groups and is simulated to the latest firm horizon among its countries, with the analogue continuation supplying the ENSO input where needed. Summaries by group, and the country panels of figs. S22 and S23, censor each country at the firm horizon of its own group; the attribution of the 2026 forecast reads every group over one calendar window for Africa (see Section S1.6.13). Each replicate thus combines one ENSO trajectory, one posterior draw and one filtered terminal state.

##### S1.6.13 Counterfactual design

In the counterfactual arm we replace the ENSO trajectory over the forecast horizon with trajectories of unforced internal ENSO variability, drawn from the pre-industrial control archive of Lenssen et al. [65]. With the neutral analogue set of the main analysis, the difference between factual and counterfactual forecasts gives the cholera outbreak burden attributable to the observed and forecast ENSO state relative to ENSO-neutral conditions; with the full pool of analogues, it is the burden relative to unforced internal ENSO variability of either sign (below).

The counterfactual trajectories are anchored in May 2026, the last month before the 2026–27 El Niño developed, and the counterfactual arm is launched from the filtered state at the end of April 2026, so that the counterfactual ENSO departs from the observed ONI in the same month in which the epidemic state is released from the observations. Every attribution statistic compares this arm with an *attribution factual*: the factual arm launched from the same end-of-April state, with the same smoothing replicate, random number seed and posterior draw for each replicate, so that the two arms differ only in their ENSO trajectory. The forecast shown in the figures is a third arm, launched from the August 2026 state, which retains the national counts observed from May to August 2026; it enters no attributable quantity. The difference between the displayed forecast and the counterfactual decomposes per posterior draw into an ENSO component (attribution factual minus counterfactual) and an information component (displayed forecast minus attribution factual), the latter being the effect of the months observed between the two launches (GitHub data folder, files `D22_attrib_information_scope` and `D22_attrib_information_decomposition`). The end-of-April state is itself a smoothed state, reconstructed from observations to August 2026, so the information component compares the realised May to August record with its re-simulation, not with a forecast made in April.

The counterfactual analogues come from the same library as the factual continuation (Section S1.6.12), with the same rescaling, features and excluded run. From the last observed month before the launch, we select the 100 analogues per model nearest in ONI level, three-month tendency and ONI six months earlier, standardised by the library’s feature standard deviations and at least 12 months apart within a model, over 10 models. Each path is joined to the observed ONI of the launch month, with the offset between analogue and observation decaying with an e-folding time of three months, so that both arms start from the observed ONI and differ only through the analogues’ evolution.

We classify every December to February (DJF) winter read by a reported month (the month minus the group’s ENSO lag) on the mean of the monthly ONI over December to February, with thresholds of *−*0.5 and +0.5 °C and no persistence requirement. A trajectory belongs to the La Niña, neutral or El Niño set if its first winter has that phase and every later winter it reads is neutral; otherwise it is labelled mixed and kept only in the full pool. For the 2026 forecast, the pool of 1,000 trajectories gives 30 La Niña, 151 neutral and 208 El Niño analogues; set sizes, model shares, seam errors and the mean ONI of every classified winter are given in GitHub data folder, files `D29_analog_sets`, `D29_analog_sets_winters` and `D29_analog_sets_models`. The main analysis draws counterfactuals from the neutral set, which asks what the coming season would hold without an ENSO event of either sign; the full pool and the two other conditional sets are reported in the supplementary figures.

We write 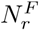 and 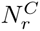 for a burden summary, either the number of outbreaks or total cases, under the factual and counterfactual arms in replicate *r*, using the same posterior draw and random number stream in both arms, and report two estimands. The first is the ratio of expectations,

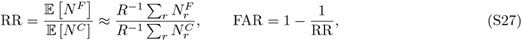

which we map on a log_10_ scale. We use the ratio of expectations rather than the expectation of ratios because most administrative units have no outbreak in most replicates, so that per-replicate ratios are mostly undefined or infinite. The second is the paired difference,

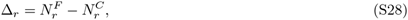

which is defined in every replicate and provides the uncertainty statement: we report a shift as credible when a central interval of the empirical distribution of *{*Δ*_r_}* excludes zero. Pairing removes the stochastic and parameter variation shared by the two arms, such that Var[Δ] is much smaller than Var[*N ^F^*] + Var[*N ^C^*]. Probability-of-increase maps are not paired. Results for each analogue set are shown in figs. S24 and S25, and the forecast and neutral-ENSO counterfactual of each country in figs. S22 and S23.

The attribution covers one calendar window for every ENSO group, from the attribution start to October 2027 (the firm horizon of group GS, ENSO lag +4 in the occurrence model), and summaries for Africa as a whole sum the groups over that window. Groups whose firm horizon ends earlier are read on into the analogue continuation of ENSO, both arms alike, and groups forecast further are cut at the window end; GitHub data folder, file `D23_africa_scope` gives each group’s longest continuation lead per model and the skill of the analogue continuation against climatology at that lead. The counterfactual constrains every winter read by the reported months to be neutral.

##### S1.6.14 Attribution metrics

We summarise the paired factual and counterfactual simulations with five metrics, each answering a different question. All are computed from the same replicates, per first-level administrative unit and for each country, ENSO group and Africa as a whole. For a first-level unit *j* and replicate 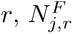 and 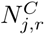 are the total cases or the number of outbreak onsets over the comparison window in the factual and counterfactual arms. Scope totals sum their units within each replicate, 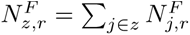; countries and ENSO groups use the replicates paired within their group, and Africa uses the replicates present in every group, so that no total mixes draws from different replicates. The main results keep every replicate. As a sensitivity analysis, a replicate pair is removed from every unit of an ENSO group (and from Africa) when, in either arm, any unit of the group reaches 2 *×* 10^5^ cases in a month of the window or the cumulative cases of the group over the whole window reach 10^6^; in the posterior intervals below, the pair is removed from every group of its transmission unit, so that the groups of a unit keep one set of replicates. As this conditions on the outcome, and the factual arm reaches these thresholds more often where ENSO raises incidence, it pulls the attributable fraction towards zero. The pairs removed because of each arm and the resulting change in the attributable fraction are given in GitHub data folder, file `D24_explosive_pairs`. We report two comparison windows: the main window, from the common launch in May 2026, and, as a sensitivity analysis, a window restricted to the months after the launch of the displayed forecast in September 2026. In both, the attribution factual and the counterfactual are free simulations from the same state.

The intervals of the attributable number and fraction, and of the exposed population of the ratio-of-means rule below, are posterior intervals of the ENSO effect. For posterior draw 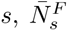 and 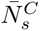 are the mean window totals over the *R/S* replicates that ran *s* (Section S1.6.7), summed over the units of a scope at the same draw (the draw indexes the fit of the unit’s ENSO group, so that a scope within one group sees one parameter vector), and 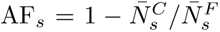. These per-draw means still carry the epidemic variation of their replicates, so the variance of AF*_s_* across draws overstates the posterior variance by the within-draw noise, as in a nested simulation whose inner samples are the replicates of one draw [66], or in the analysis of variance that separates internal variability from the spread between climate-model ensemble members [67]. We write *v̂*_noise_ for the mean over draws of the delta-method variance of AF*_s_*, computed from the within-draw variances and covariance of the replicate totals, and *v̂*_draws_ for the variance of AF*_s_* across draws, so that *v̂*_draws_ *− v̂*_noise_ is the method-of-moments estimate of the posterior variance. Rather than report that variance alone, we shrink AF*_s_* towards the pooled value by 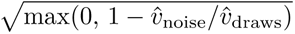 before taking the 2.5% and 97.5% quantiles over draws; this rescaling keeps the centre and shape of the ensemble of draws and matches its variance to the estimate, the constrained Bayes estimator of a population of parameter values [68, 69], in place of the conditional-mean shrinkage that would narrow the ensemble by the full noise share. When the estimated noise share *v̂*_noise_*/v̂*_draws_ of a scope or first-level unit reaches 0.8, the replicates cannot separate its posterior spread from their epidemic variation. We then report its pooled value with the range of its replicates and no posterior interval; the figures draw that range dashed. The interval of the exposed population of the ratio-of-means rule covers the first-level units whose spread is identified, and the population of the others is reported separately, with the part whose pooled estimate passes the rule. Africa is computed like an ENSO group: the replicate totals of every simulation unit are summed within each replicate before the per-draw means, so that groups simulated together in one transmission unit keep their dependence. Draw *s* indexes the fit of each group and transmission units are simulated independently, so the sum at one draw index is a joint draw. Replicate quantiles of 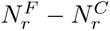 (below) describe the spread of single paired simulations, epidemic noise included; so do the cumulative window totals of the main-text forecast figure, shown as the median and 50% and 95% quantiles across paired replicates, whereas its attributable fractions are the noise-adjusted posterior intervals above. Population counts use the 2025 population of the observed reporting units of each simulated first-level unit, and shares are of the population of the simulated first-level units in the scope.

###### Attributable burden

The attributable number and fraction of cases or onsets are

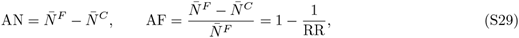

where 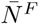 and 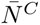are means over replicates and RR is the ratio of expectations of eq. (S27), so that AF coincides with the FAR of that equation. These are the attributable number and fraction of health impact attribution [70, 71, 72]. We also report the replicate quantiles of 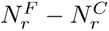 and of 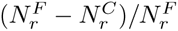, and the attributable number per 100,000 population.

###### Probability ratio

For an outbreak event *E* we compare its probability in the two arms, 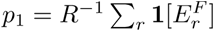 and 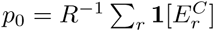, through

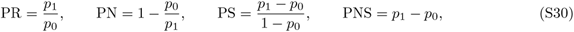

the probability ratio of event attribution [73] and the probabilities that the ENSO forecast was a necessary, a sufficient, and a necessary and sufficient cause of the event [74]; PN is the fraction of attributable risk of event attribution. We keep the sign of PN, PS and PNS, so that negative values indicate an event made less likely. We use two events: at least one outbreak onset in the window, and window cases of at least 50, the outbreak-size threshold of the episode database. When no counterfactual replicate has the event while some factual replicates do, PR is infinite; we then report as lower bound 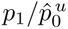, where 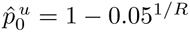 is the exact one-sided 95% upper bound on *p*_0_ given zero events in *R* replicates [75, 76]. PS is undefined when *p*_0_ = 1. In large scopes both arms almost always contain the event and PR tends to one, so we interpret the probability ratio at the first-level-unit and country level.

###### Population with a larger expected burden

Following Childs et al. [72], a first-level unit counts towards the exposed population at magnitude *m ∈ {*0.25, 0.5, 1*}* when its mean factual total is positive and at least (1 + *m*) times its mean counterfactual total; a positive factual mean against a zero counterfactual mean counts, and two zero means do not. We report the posterior mean and 95% credible interval of this population, applying the rule to each draw’s shrunk AF*_s_*, using 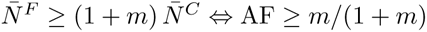. We also report the rule on replicate medians, which is not driven by the few replicates with very large outbreaks; it has no interval.

###### Population with a robust change

Following the robustness assessment of the IPCC AR6 Atlas [77], with *m* as the variability threshold, a unit has a *robust* increase when at least 66% of the paired replicates have 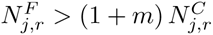 and at least 80% have 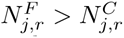 (ties counting as disagreement), a *conflicting* signal when only the first condition holds, and *no robust signal* otherwise.

###### Emergence of the signal

With the paired difference 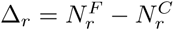 of (S28), the signal-to-noise ratio

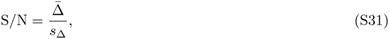

where *s*_Δ_ is the standard deviation of Δ*_r_* across replicates, measures the change relative to the epidemic variability that remains after pairing [78]. We report the population of the units in which the signal has emerged, S*/*N *≥ γ* for an increase, S*/*N *≤ −γ* for a decrease, at *γ* = 1 and *γ* = 2. Unlike the robustness classes, this measure needs no magnitude threshold; for a normally distributed difference, S*/*N = 1 corresponds to an increase in 84% of the replicates.

The first two metrics quantify how much burden, and how much more likely an outbreak, the forecast ENSO state implies; the last three locate the population where the expected burden is higher by a margin, where the replicates agree on the change, and where the change stands out from the epidemic noise.

##### S1.6.15 Testing of predictions for 2026 cholera incidence (West)

For the West group we compare the modelled ENSO effect on 2026 with the WHO national counts (fig. S26). The modelled ratio is the mean of the historical reconstruction over the mean of its neutral-ENSO counter-factual, over the only months both arms cover, May to August 2026 (the counterfactual is launched from the reconstruction in May, and the reconstruction ends in August). Each counterfactual replicate is paired with the reconstruction replicate it was launched from, and the interval is the spread of the ratio across the reconstruction replicates. The observed ratio is the 2026 WHO count over the mean of the ENSO-neutral years since 2010, standardised to the 2026 population, with one pseudo-case on each side; a year is neutral when its mean ONI lies within *±*0.5 °C. It has two estimators: the same calendar months (May to August), for which the WHO monthly series covers only the neutral years 2024 and 2025, and the 2026 year to date against full neutral years, a lower bound. Countries without a WHO series are left out rather than counted as zero, and Somalia, whose 2026 series stops in January, is censored. Over the countries with an observed ratio, the modelled West ratio is 7.0 (95% interval over replicates 3.9 to 14.4), against an observed 7.4 for the same months (Cameroon and Congo) and a lower bound of 7.0 for the year to date (Central African Republic, Cameroon, Congo and Chad; modelled 6.3 for these countries).

##### S1.6.16 Attribution of past ENSO events

We apply the paired design of the forecast attribution to three strong ENSO events of the study period: the El Niño events of 2015–16 and 2023–24 and the La Niña of 2020–23, attributed as one event (table S10). For each event we launch both arms in the month before the ONI crossed +0.5 (El Niño) or *−*0.5 (La Niña) and stayed beyond it until the event’s first peak: September 2014, August 2020 and May 2023. For the 2020–23 La Niña, whose ONI returned above *−*0.5 in May to August 2021, this persistence applies to the first of its three winters only. Both arms start from the reconstruction state of the month before the launch (Section S1.6.8), which has seen the observations of the event, and are paired as in the forecast: same reconstructed state, posterior draw and random number stream, with the replicates and draws of the forward arms (Section S1.6.7).

The factual arm uses the observed ONI throughout, as the hindcasts do (Section S1.6.10), but runs over the whole event window from a single launch and is not scored. The counterfactual arm replaces the ONI from the launch month onwards with analogue trajectories from the pre-industrial control archive of Lenssen et al. [65], built as in Section S1.6.13 and joined to the observed ONI of the launch month. We draw from the neutral set, so that the counterfactual describes the seasons had the event not developed. The ONI of the months before the launch is the observed one in both arms, and, as in the forecasts, the ONI entering each model is clamped to its widened boundary knots (Section S1.6.11).

The 2020–23 La Niña is attributed as one event: one launch (August 2020, the month before the ONI crossed *−*0.5), one pair of factual and counterfactual runs per transmission unit, and counterfactual analogues neutral in each of the event’s three La Niña winters (DJF 2020/21, 2021/22 and 2022/23). Later winters read by the reported months (DJF 2023/24) are not constrained. The event’s counterfactual has 83 neutral analogues. Results are reported for the whole event and for consecutive per-winter sub-windows.

Windows are set per country. The comparison window runs from the launch to the end of the country’s cholera year containing the peak of the event (see Section S1.1.6), extended by the largest positive lag selected for its group across the three models, so that the lagged response of every model to the peak falls within the window (the hindcasts use the smallest lag instead, Section S1.6.10). For the La Niña, the *k*-th sub-window ends with the cholera year containing the *k*-th winter’s ONI peak plus the same lag, and the next sub-window starts the month after; the sub-windows partition the event window, so their attributable numbers add up to the event’s, but their attributable fractions do not. A transmission unit is simulated to the latest window end among its countries, and each country is censored at its own window end. As windows are set per event, the later La Niña sub-windows may overlap the window of the 2023–24 El Niño, and the burdens of the two events are then not additive.

We read the factual arm in two ways. The first is the free simulation with observed ENSO. The second replaces, over the whole window, the factual series of replicate *r* with the historical reconstruction of the filtered replicate it starts from (see Section S1.6.8). This anchors the factual arm to the observations, but it conditions that arm on data the counterfactual arm does not see, so that the contrast also contains the misfit of the model to the observations. The two arms of this reading also treat the index differently: the reconstruction does not clamp the ONI, whereas the counterfactual arm holds it within the widened knots. As the reconstruction has 100 replicates, this reading has at most as many distinct factual series.

From each pair of arms we derive the attributable number and fraction (S29), the probability ratio and probabilities of causation (S30), the signal-to-noise ratio (S31) and the two population metrics, as in Section S1.6.14, with every replicate kept, the same scope totals and, for the free simulation, the same posterior intervals over draws (fig. S20); the reading with the reconstruction as factual is a point estimate. In the main-text figure, the bars of Africa and each ENSO group show the median of this noise-adjusted posterior with its interval, whereas the per-unit maps show the attributable fraction of the replicate means, a point value without interval. As both arms launch in the same month, there is a single comparison window per country.

To relate the simulated fraction to the observed burden, we also report the observed-anchored attributable number

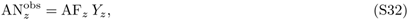

where *Y_z_* is the observed total of scope *z* over its simulated unit-months, with its interval from that of AF*_z_* and *Y_z_* held fixed. Observed onsets are taken from the occurrence dataset and observed cases from the monthly case-count dataset, using for each unit-month the largest total among its reporting levels (see Section S1.1.7). We report alongside the share of simulated unit-months in the occurrence dataset, as a measure of surveillance coverage, and the ratio of *Y_z_* to the mean factual total, as a check of the factual arm.

As a cross-check without epidemic dynamics, we compute the burden attributable to each event directly from the fitted models on the observed data [70]. For an observed unit-month (*i, t*) in the window with outcome *y_it_* (an onset in the occurrence model, cases in the monthly case-count model), posterior draw *s* and a counterfactual trajectory 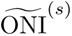 drawn for that draw from the event’s neutral analogue set,

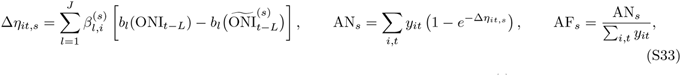

where *b_l_* and *L* are the basis columns and selected lag of the model (S3), 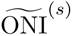 equals the observed ONI before the launch, and 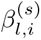 combines the population coefficients with the country and first-level random slopes of unit *i* (the country-level coefficients for a unit without a random effect). Both ONI series are clamped to the widened knots as in the simulations. We report the mean and 2.5% and 97.5% quantiles over 400 posterior draws of the fitted models (the simulations use 100 per model), which carry both parameter uncertainty and the spread of the counterfactual ENSO trajectories. This calculation uses every observed unit-month of a group’s countries, including reports at coarser levels that the simulations do not represent, and it ignores the effect of ENSO on contagion and on the immunity carried between seasons.

#### S1.7 Software and reproducibility

We fitted all models in R [79] using Stan [49] through the `brms` interface [80] and the `cmdstanr` backend [81]. We constructed exposure bases with the dlnm package [82], computed approximate leave-one-out cross-validation with the loo package [53, 83], and ran the block particle filter with the `spatPomp` package [30].

##### S1.7.1 Use of AI-assisted tools

The analysis code (data processing, model fitting, simulation, postprocessing and figure code) was developed with the assistance of the AI coding agent Claude Code (Anthropic) between May and November 2026, using the large language models Claude Opus 4.6, Opus 4.8, Opus 5 and Opus 5.5, Claude Sonnet 4, Sonnet 4.6, Sonnet 5 and Sonnet 5.5, Claude Fable 5.1 and 5.2, and Claude Haiku 4.5 and 5.5 for auxiliary agent tasks. Working in the authors’ code repository under their direction, the agent wrote and edited code and ran analysis scripts on the authors’ workstations and computing clusters. The authors made all decisions on study design, data inclusion, model specification, priors and interpretation. Code changes were reviewed by the authors and checked with an automated test suite; code restructurings were required to reproduce prior outputs exactly. OpenAI Astra 6 was used for a secondary code review. AI tools did not generate, alter or impute any data: all reported estimates come from the statistical models and simulations described here. The analysis runs without AI tools from the publicly available code (Section S1.7.2).

##### S1.7.2 Data and code availability

The analysis code, the canonical outbreak episode tables, the derived model inputs, the monthly WHO national series and the GitHub data folder cited in this supplement are publicly available at https://github.com/GenevaIDD/enso cholera africa and archived at [Zenodo DOI upon acceptance]. Outbreaks were extracted from the Global Cholera Taxonomy Database with the OutbreakExtractR package, available at https://github.com/HopkinsIDD/OutbreakExtractR. The raw outbreak database is available from the authors on request, under its terms of use. The files of data S1 accompany this manuscript and are listed with their content under Data S1 after the references. The ENSO index, the ENSO forecasts, the pre-industrial ENSO trajectories, the population estimates and the administrative boundaries are publicly available from the sources cited in Materials and Methods.

### S2 Supplementary Text

#### S2.1 Limitations

Although our analysis builds on the most comprehensive continent-wide database of cholera, it has several limitations. First, our data are suspected cases, with case definitions and reporting that vary between countries and over time. Where confirmation is reported, episodes with a confirmed case have onset-rate contrasts by ENSO phase broadly consistent with those of all episodes, where the data allow the comparison (table S4), which suggests that the suspected-case signal is not driven by non-cholera diarrhoea. Reporting that intensifies around outbreaks independently of ENSO would tend to dilute an ENSO effect rather than create one.

Second, the ENSO teleconnection groups and the lag of each model are selected on the data the grouped models are fitted to. The partition is the dominant mode of one of several nearly equivalent fits (Section S1.4.2), and the selected lags are often within two standard errors of others (Section S1.4.5). Because we select the lags on a fixed partition after the grouping pass, we do not propagate the grouping uncertainty to our ENSO effect estimates, although the overall patterns were consistent across the numbers of groups we tested, despite multimodality across MCMC chains. We also did not account for the Indian Ocean Dipole, which is known to act synergistically with ENSO on climate patterns in Africa; it could explain some of the remaining uncertainty in the ENSO effects and is a subject for further study.

Third, neither our inferential models nor our outbreak simulator account for external drivers such as conflict or displacement, nor for cholera prevention and control measures such as vaccination or water, sanitation and hygiene (WASH) interventions. There is evidence that ENSO increases the risk of conflict, so we believe that our estimates represent a total effect of ENSO through these pathways; in the occurrence model, which conditions on recorded outbreak history and contagion, they are direct effects with respect to those terms. Reactive cholera control may have shortened outbreaks, reduced their size or prevented large outbreaks from igniting, possibly diluting the association between ENSO and cholera. Accounting for these interventions is challenging because unified continent-level data on their implementation over the study period are scarce, but including them in future analyses would improve our estimates of how ENSO is associated with cholera outbreaks in Africa. More technically, we use a single source of ENSO forecasts, one calibrated CFSv2 issuance, without multi-model or issuance-to-issuance uncertainty, continued past each group’s firm horizon by analogues whose skill falls with the lead (Section S1.6.12). Larger ensembles would make the ENSO forecast more robust, but we believe this uncertainty is marginal: ONI values were already high at the time of writing, and there is little doubt that 2026–27 will be a very strong El Niño event, although the peak ONI value it will reach remains uncertain.

Finally, our hindcasts are in-sample: we initialise the simulator from a smoothing state that has seen the scored season, and we use the observed ONI rather than the forecast values available when the simulation is launched. We therefore consider the reported hindcast skill an upper bound, with real-time skill probably lower. We nevertheless know of no other systematic quantification of seasonal cholera predictions, and the fact that these simulations outperform climatology across several regions and lead times points to a promising avenue for ENSO- or weather-informed early warning systems for cholera.

## Supplementary Figures

**Fig. S4.**
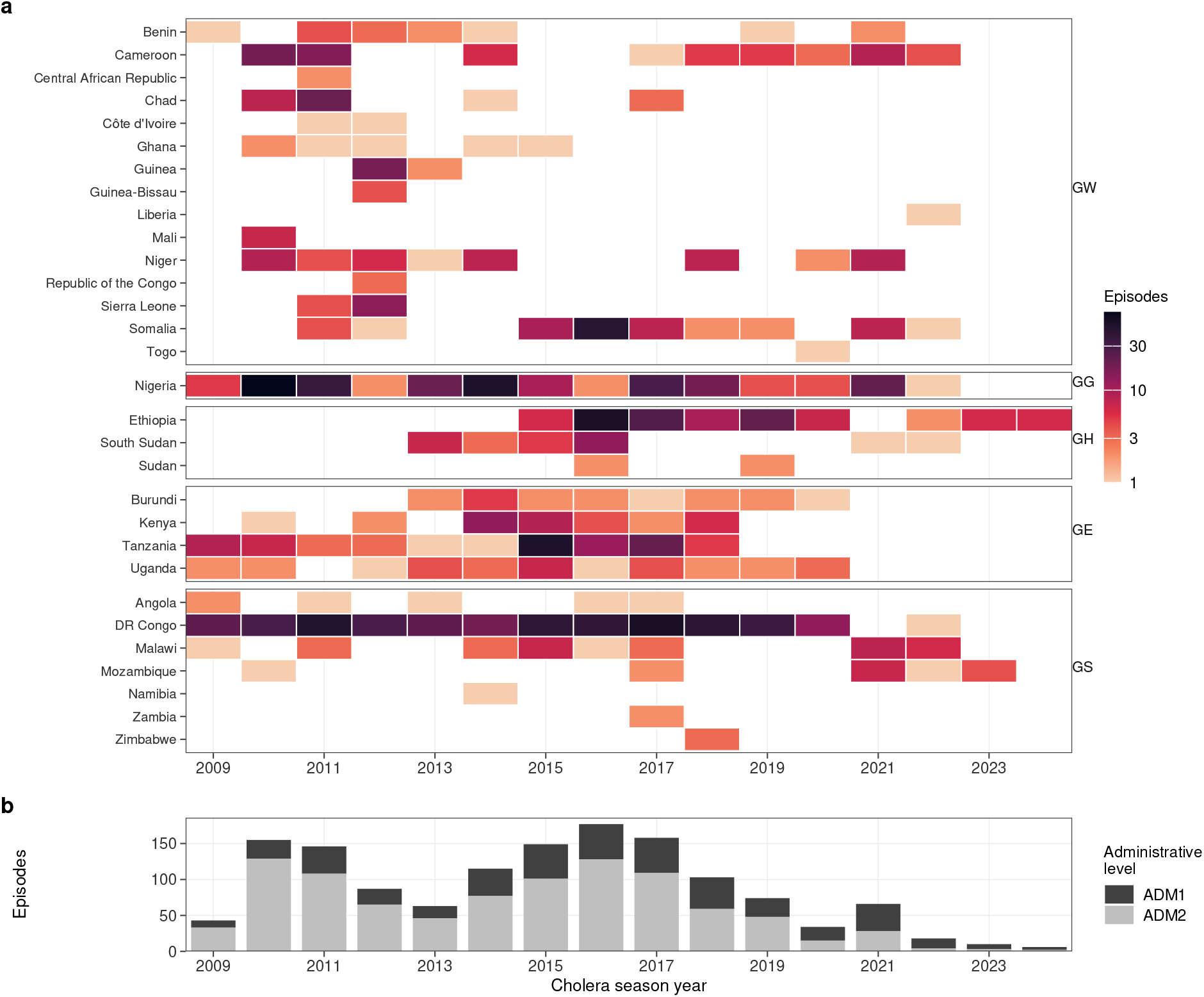
Outbreak data coverage. (**a**) Number of canonical episodes of the analysis set per country and cholera year (colour, log scale; blank: no episode), countries grouped by ENSO group and ordered by name within a group. Cholera years are labelled by the calendar year in which they start, so that the first episodes of 2010 can fall in cholera year 2009. (**b**) Number of canonical episodes per cholera year, stacked by the administrative level at which the episode is recorded. Units of analysis are mapped in fig. S5.

**Fig. S5.**
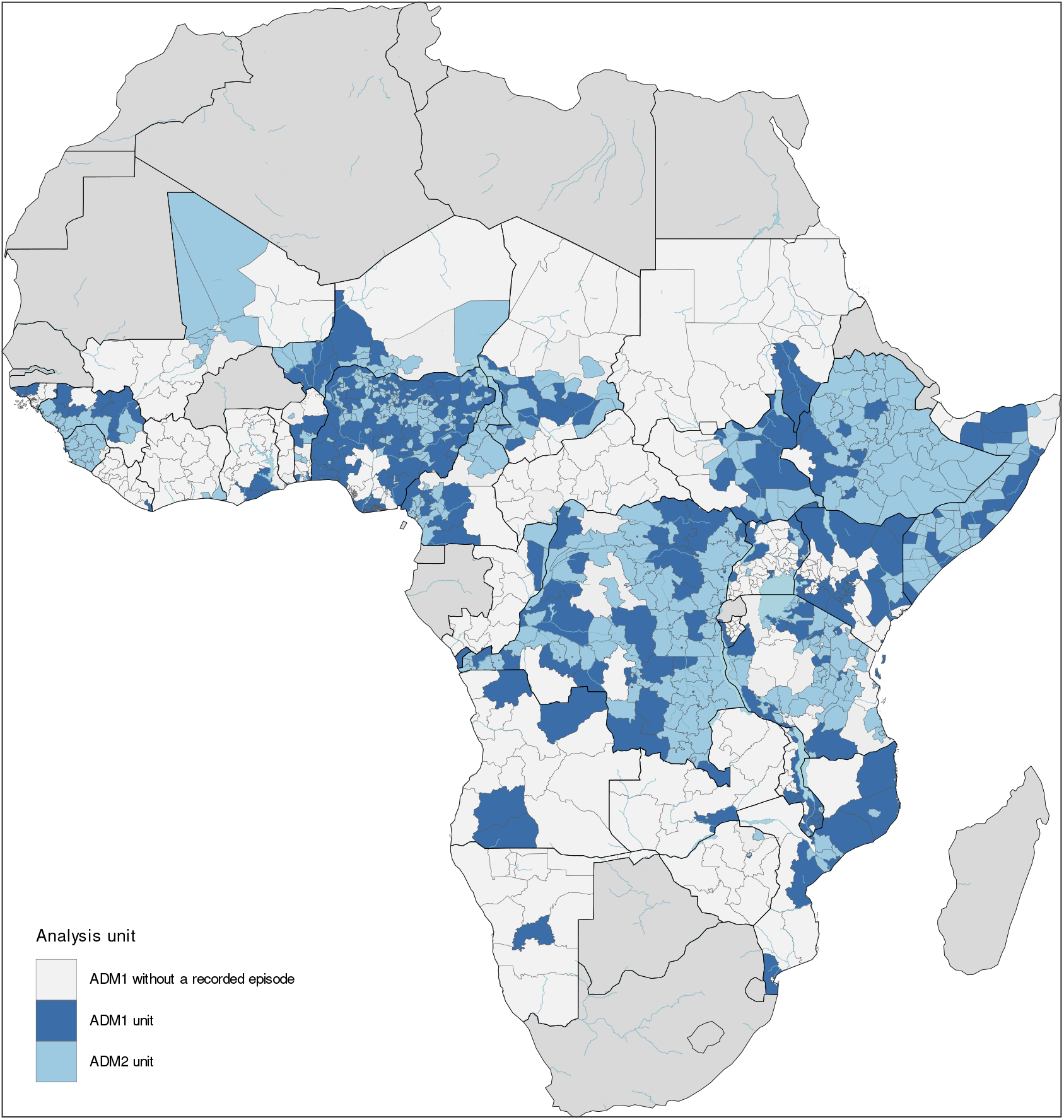
Modelled administrative units. Administrative units of the modelled countries that enter the analyses, coloured by the administrative level at which they are modelled (first or second level), and the first-level units of the same countries without a recorded episode (lightest grey). Countries outside the analysis are a darker grey. Boundaries: GADM.

**Fig. S6.**
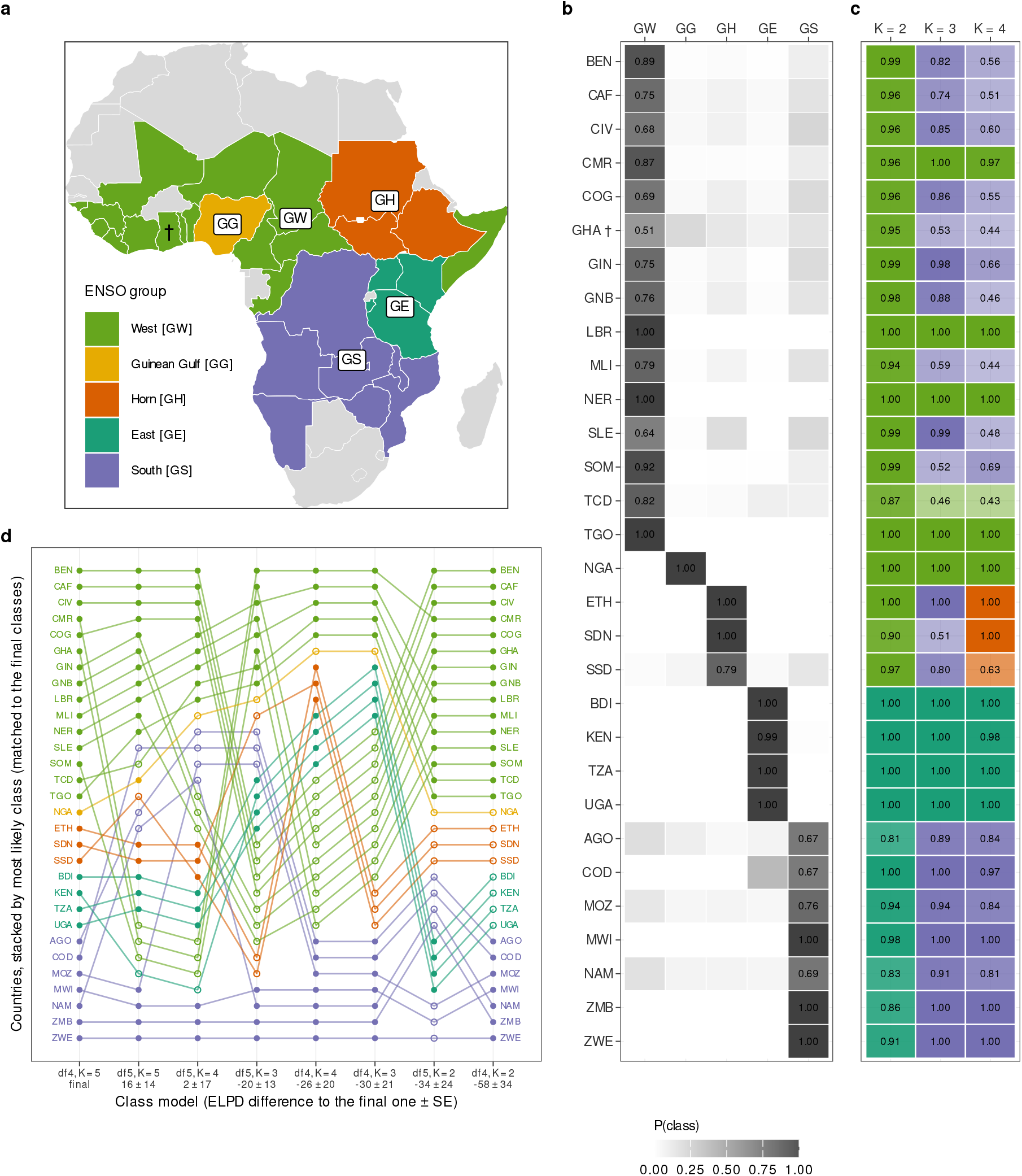
ENSO teleconnection groups. (**a**) Partition of the countries with outbreak data into the five ENSO groups used in all grouped analyses, estimated with the latent-class model (eq. S12); groups are named after the part of the continent in which most of their members lie (Section S1.4.1). Countries without outbreak data carry no group. (**b**) Posterior probability of each country (rows, grouped by assigned class) belonging to each class of the final fit (*K* = 5), with each country’s largest probability *p*_max_ printed. (**c**) Most probable class of each country, and its posterior probability, in the fits with *K* = 2 to 4 classes at the selected ENSO basis, classes matched to the final class labels and coloured as in panel a. (**d**) Class of each country in every fit of the latent-class model comparison (columns: the final fit first, then the other fits by their difference in expected log predictive density to it), classes matched to the final class labels; lines are coloured by the assigned class, and a hollow point marks a country outside its assigned class. †: countries with *p*_max_ *<* 0.6. The three agreeing chains of each fit only.

**Fig. S7.**
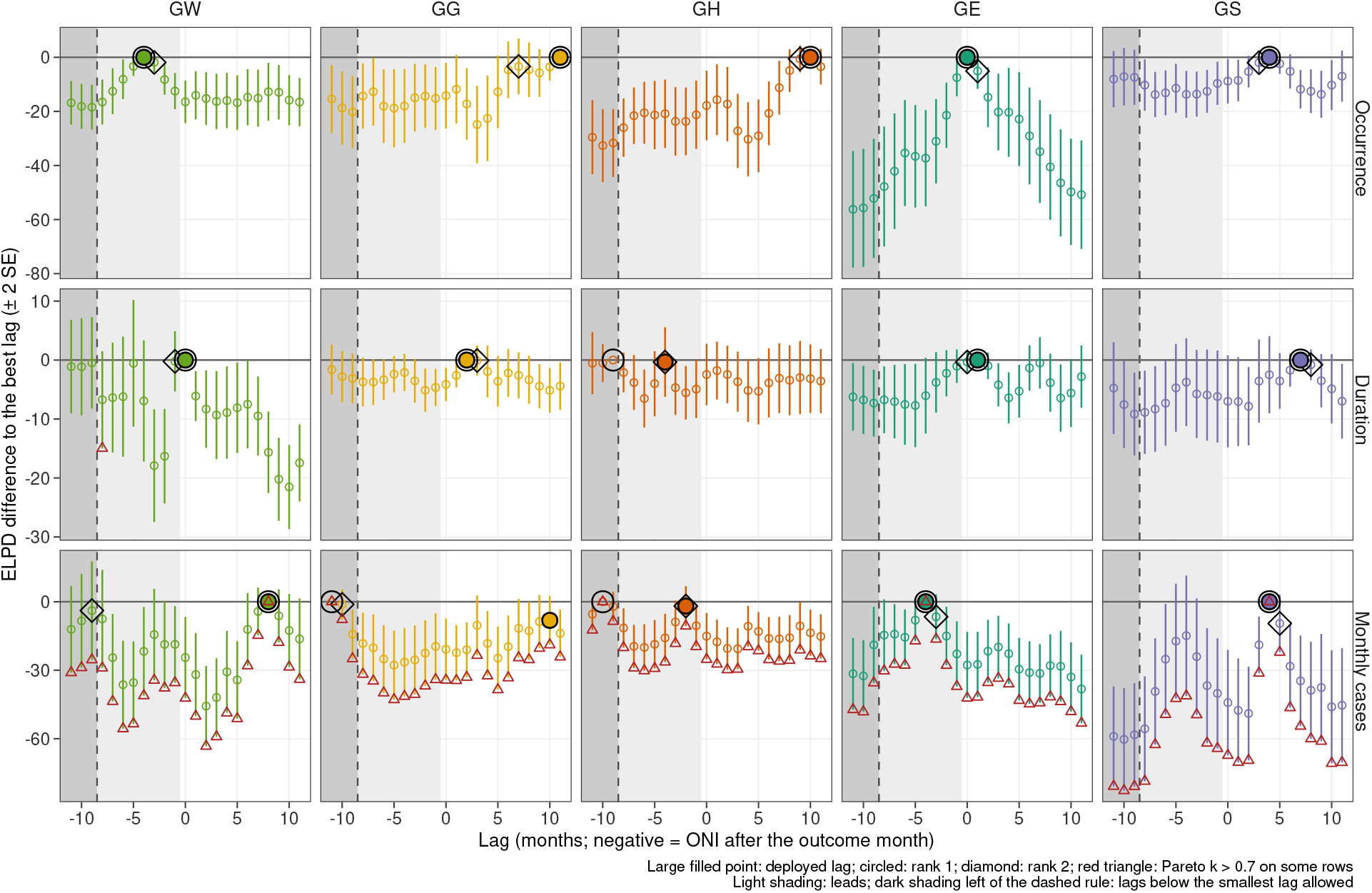
ENSO lag selection. Difference in expected log pointwise predictive density (PSIS-LOO) of each single-lag model relative to the best lag, with two standard errors of the paired difference, for lags *−*11 to 11, by analysis (rows) and ENSO group (columns). Negative lags read the index after the outcome month (light shading); lags below the selection floor are shaded darker, with a dashed line at the floor. Filled point: selected lag; circle: best lag; diamond: second-best lag; red triangle: lag with Pareto-*k >* 0.7 on some observations.

**Fig. S8.**
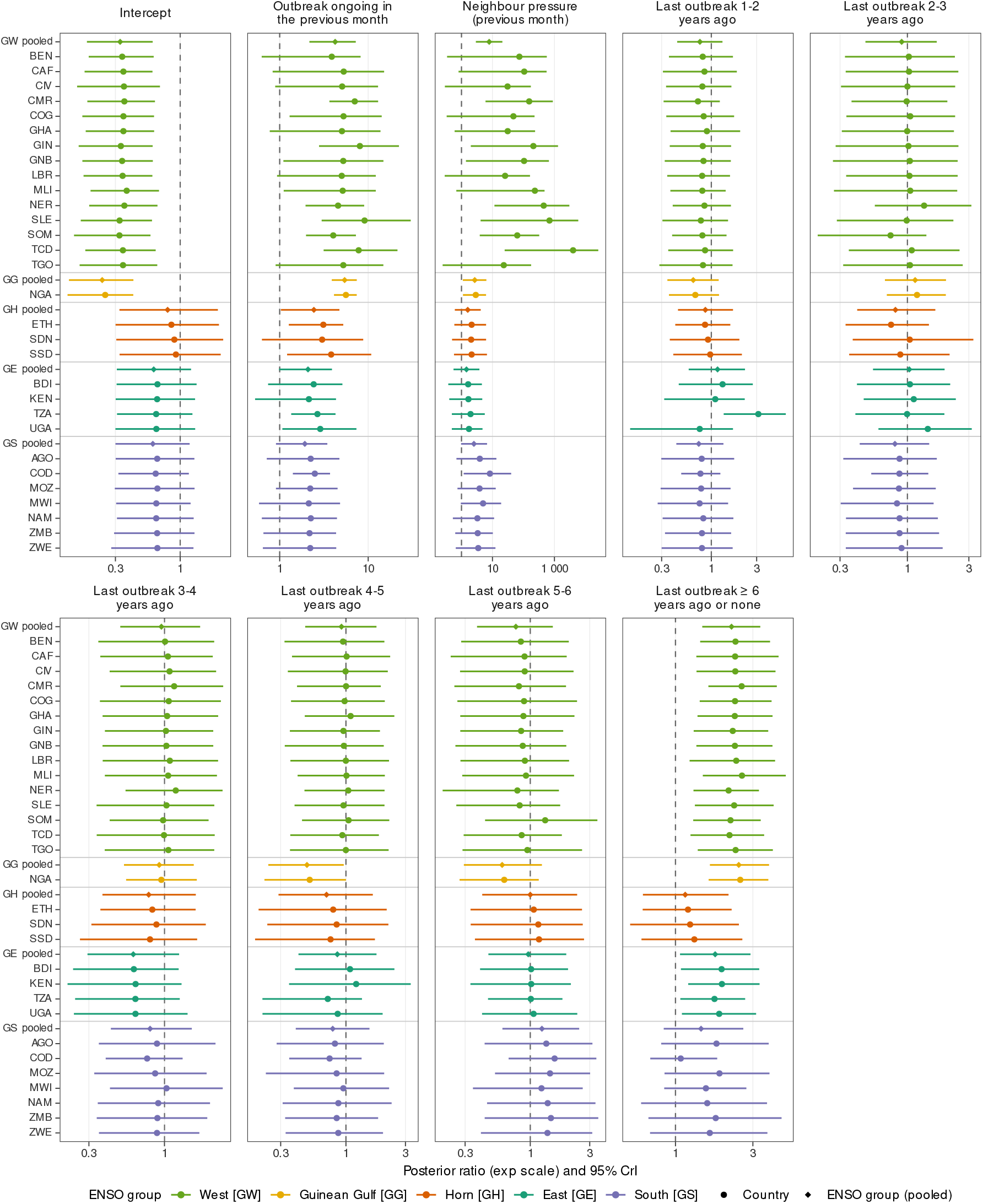
Non-ENSO terms of the occurrence model. Posterior ratio (exponentiated coefficient; point and 95% credible interval, log scale) by ENSO group and term. Intercept, within-unit contagion (outbreak ongoing in the previous month), neighbour pressure and time since the last outbreak (reference: less than one year). Circles are country-level values (population effect plus the country’s random effect), diamonds the pooled population effect of each ENSO group; countries are listed under their group’s pooled row.

**Fig. S9.**
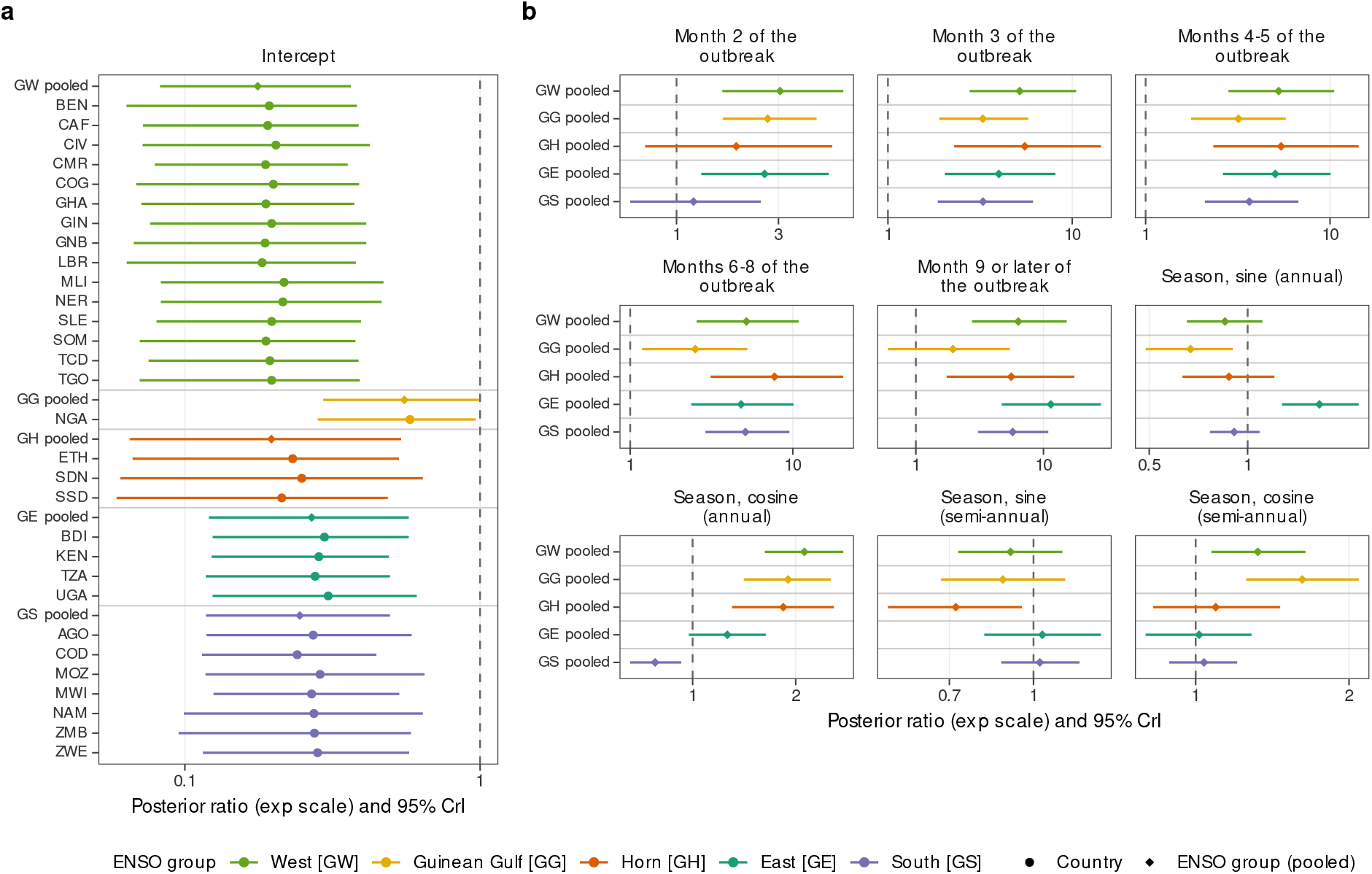
Non-ENSO terms of the duration model. Posterior ratio (exponentiated coefficient; point and 95% credible interval, log scale) by ENSO group and term. (**a**) Intercept, with a country random effect. (**b**) Episode-age bands of the cessation hazard (reference: month 1 of the outbreak) and the calendar-month harmonics of its seasonal term (annual and semi-annual sine and cosine), which have no country random effect and are shown for the ENSO groups only. Circles are country-level values (population effect plus the country’s random effect), diamonds the pooled population effect of each ENSO group; countries are listed under their group’s pooled row.

**Fig. S10.**
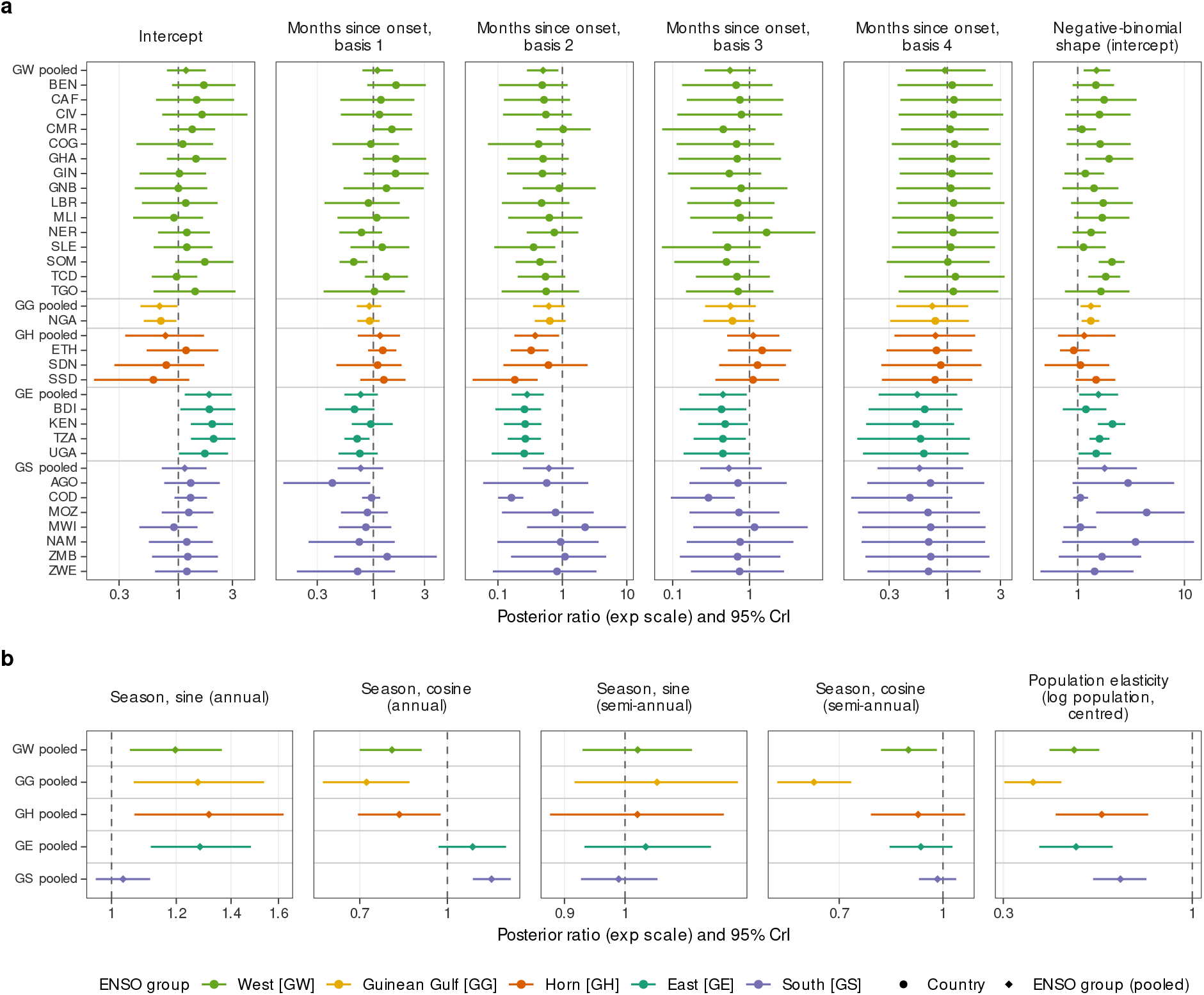
Non-ENSO terms of the monthly case-count model. Posterior ratio (exponentiated coefficient; point and 95% credible interval, log scale) by ENSO group and term. (**a**) Terms with a country random effect: intercept, the months-since-onset basis and the dispersion (shape) intercept. (**b**) Terms without one, shown for the ENSO groups only: seasonal harmonics and population elasticity. Circles are country-level values (population effect plus the country’s random effect), diamonds the pooled population effect of each ENSO group; countries are listed under their group’s pooled row.

**Fig. S11.**
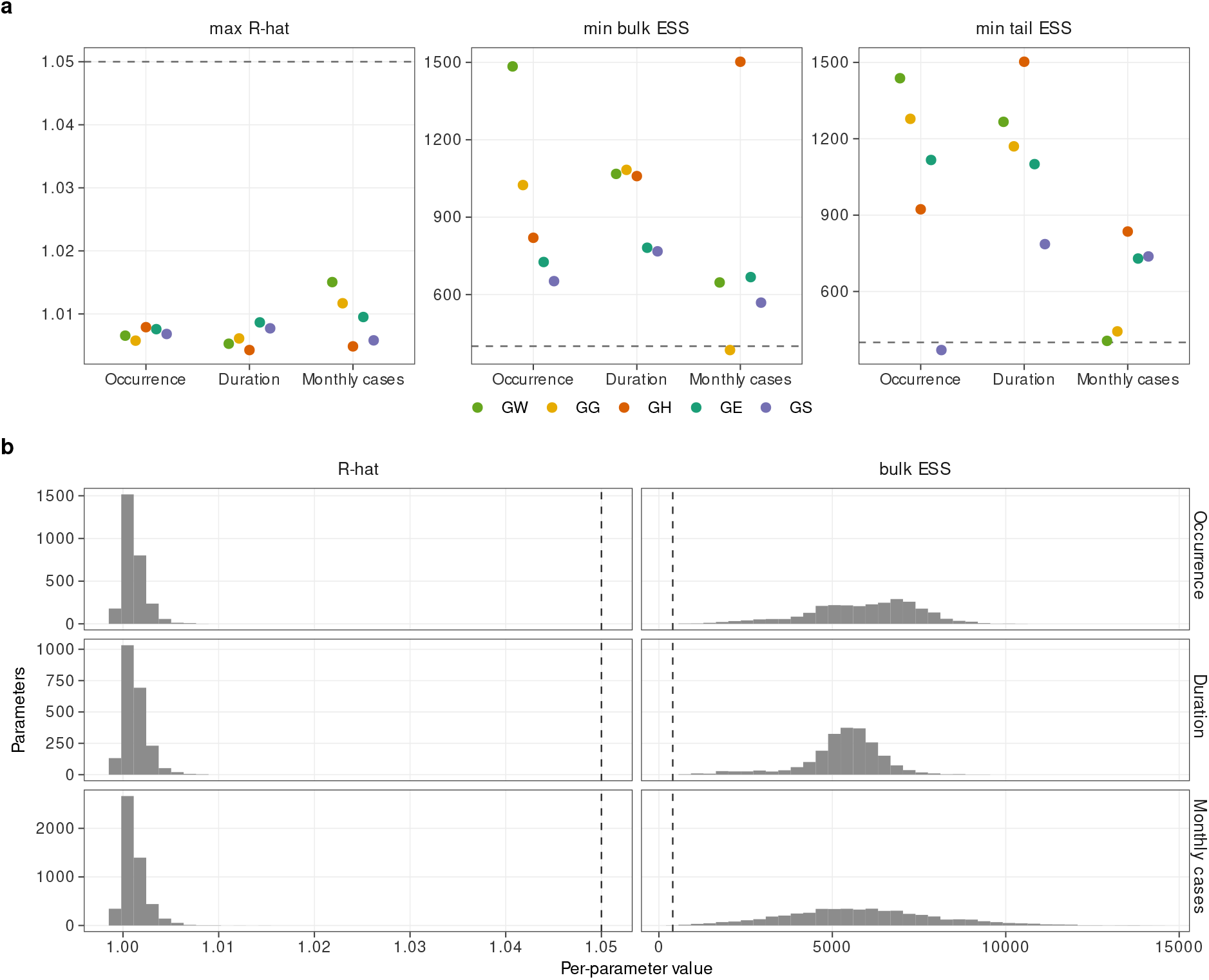
Sampler diagnostics. (**a**) Maximum *R̂* and minimum bulk and tail effective sample sizes (ESS) over the parameters of each final fit, by analysis and ENSO group. (**b**) Distribution of the per-parameter *R̂* and bulk ESS across all parameters of the final fits, by analysis. Dashed lines: the thresholds of the convergence check (*R̂* = 1.05, ESS = 400; Section S1.3.9); no fit exceeds *R̂* = 1.05; the fits that fail the check on effective sample size or divergent transitions are listed in Section S1.3.9. Bulk ESS can exceed the 4,000 draws of a fit, up to the estimator’s cap of 4,000 log_10_ 4,000 *≈* 14,400, when the sampler’s draws are anticorrelated, as for most standardised random effects.

**Fig. S12.**
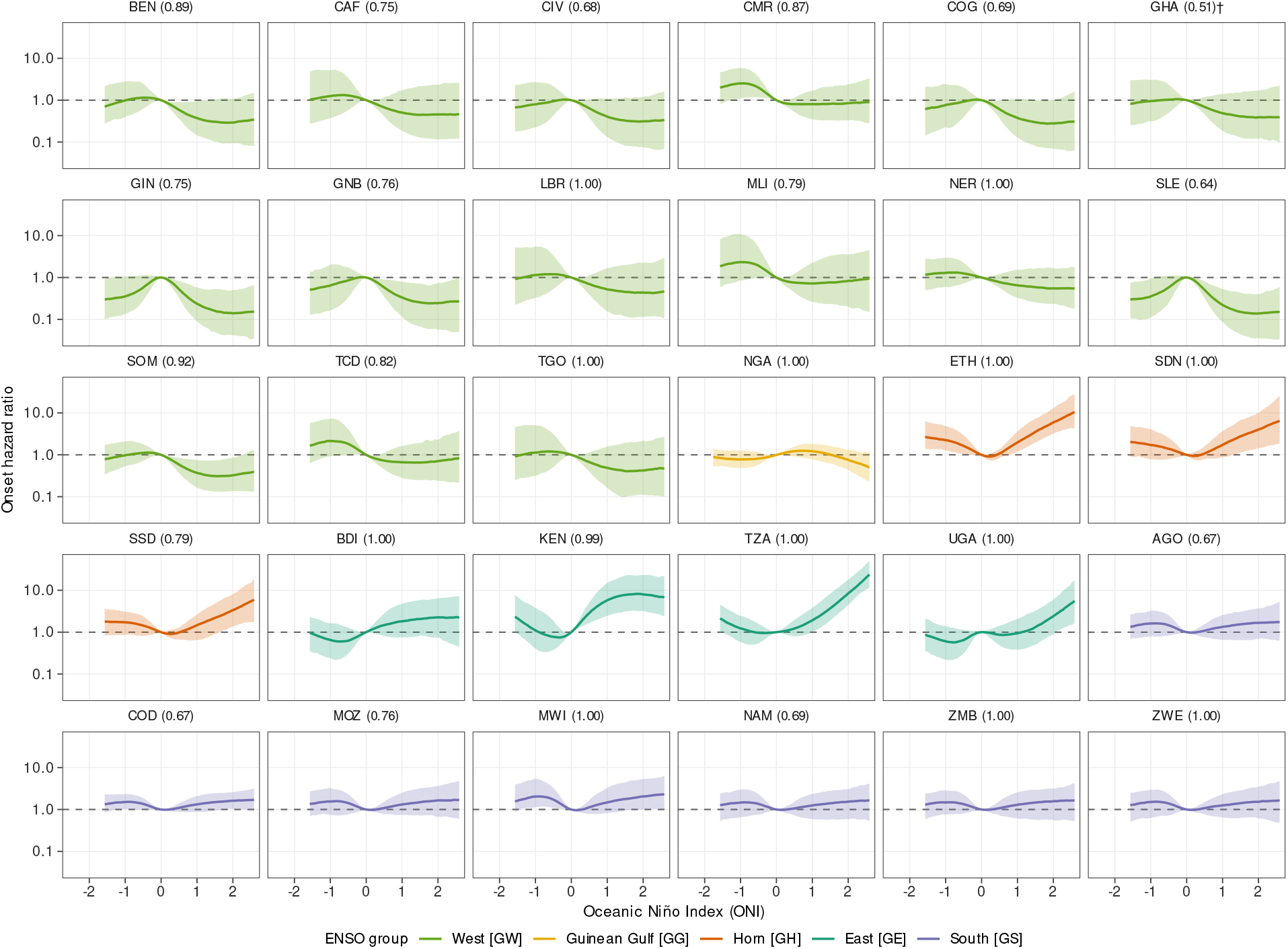
Country-level ENSO exposure–response, occurrence. Incidence rate ratio of outbreak onset against the ONI at the selected lag of the country’s ENSO group, relative to ONI = 0: posterior median (line) and 95% credible interval (band) of the population ENSO effect plus the country’s ENSO random slope, coloured by ENSO group, log scale. Each panel is labelled with the country’s largest class-membership probability *p*_max_ (**^†^** below 0.6). Conditional on the other model terms; a direct effect given outbreak history and contagion.

**Fig. S13.**
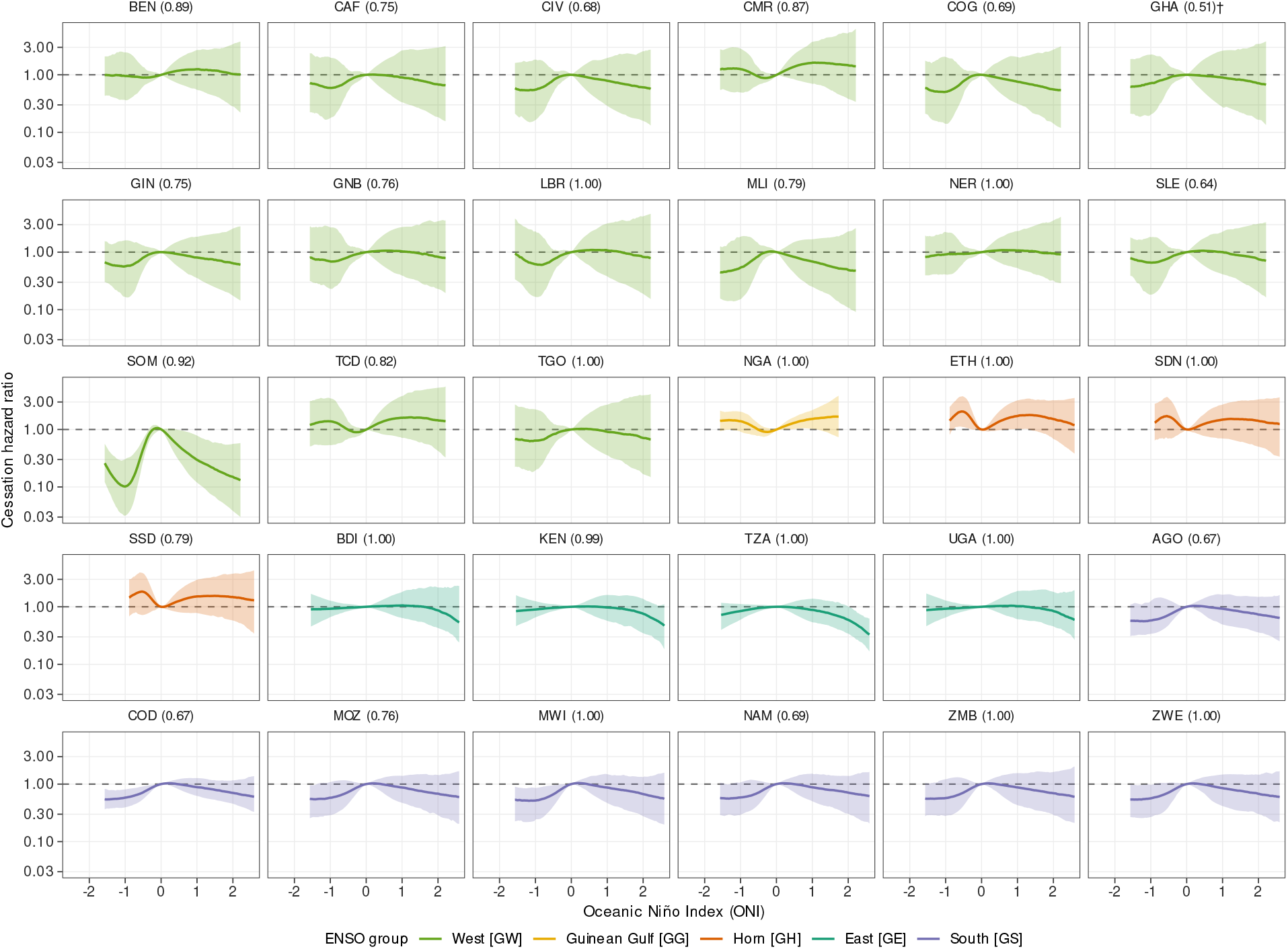
Country-level ENSO exposure–response, duration. Cessation hazard ratio against the ONI at the selected lag of the country’s ENSO group, relative to ONI = 0 (a ratio above 1 means outbreaks end sooner): posterior median (line) and 95% credible interval (band) of the population ENSO effect plus the country’s ENSO random slope, coloured by ENSO group, log scale; panels labelled with *p*_max_ as in fig. S12. Conditional on the other model terms.

**Fig. S14.**
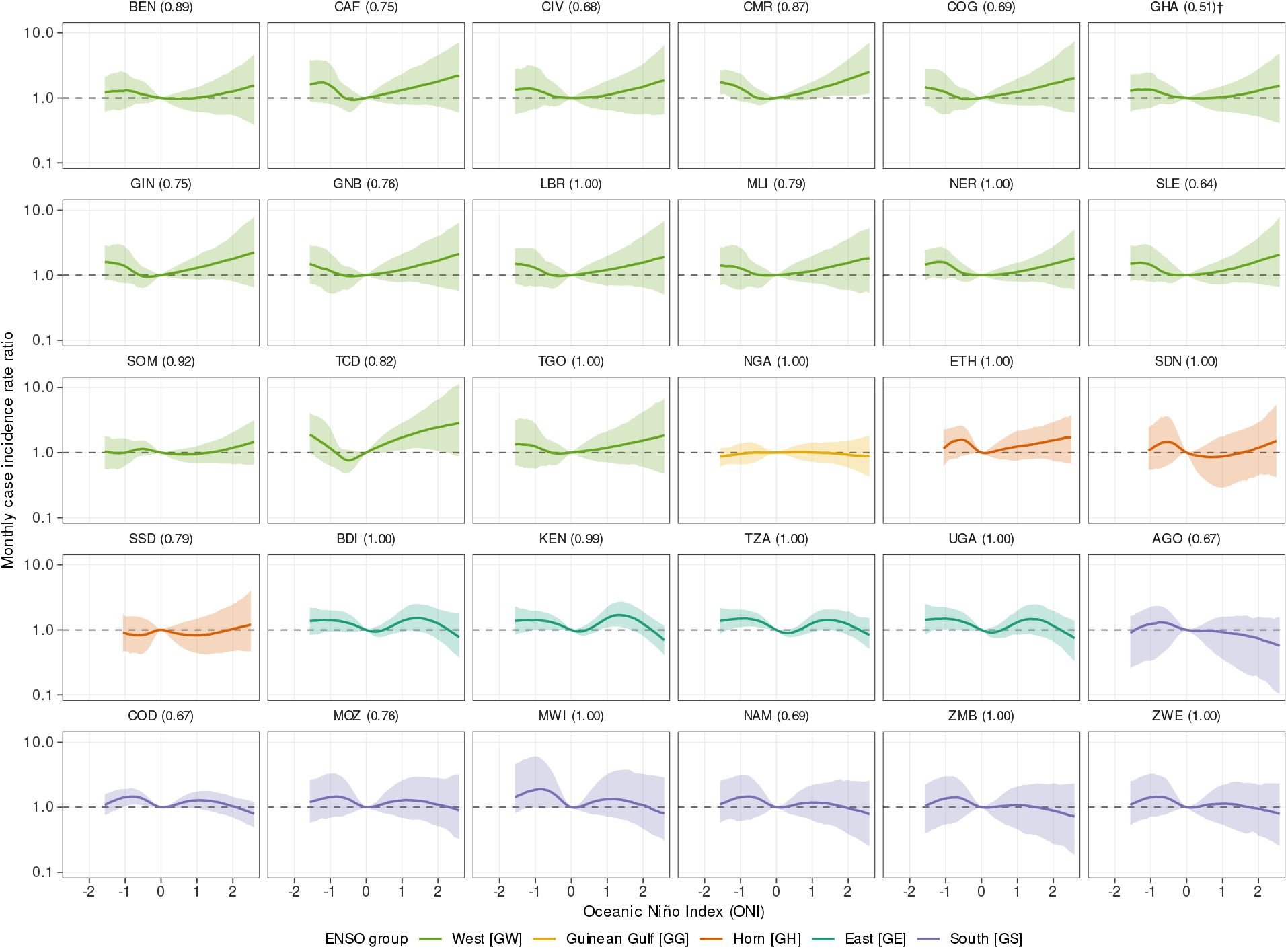
Country-level ENSO exposure–response, monthly case counts. Rate ratio of monthly suspected cases in an active episode against the ONI at the selected lag of the country’s ENSO group, relative to ONI = 0: posterior median (line) and 95% credible interval (band) of the population ENSO effect plus the country’s ENSO random slope, coloured by ENSO group, log scale; panels labelled with *p*_max_ as in fig. S12. Conditional on the other model terms.

**Fig. S15.**
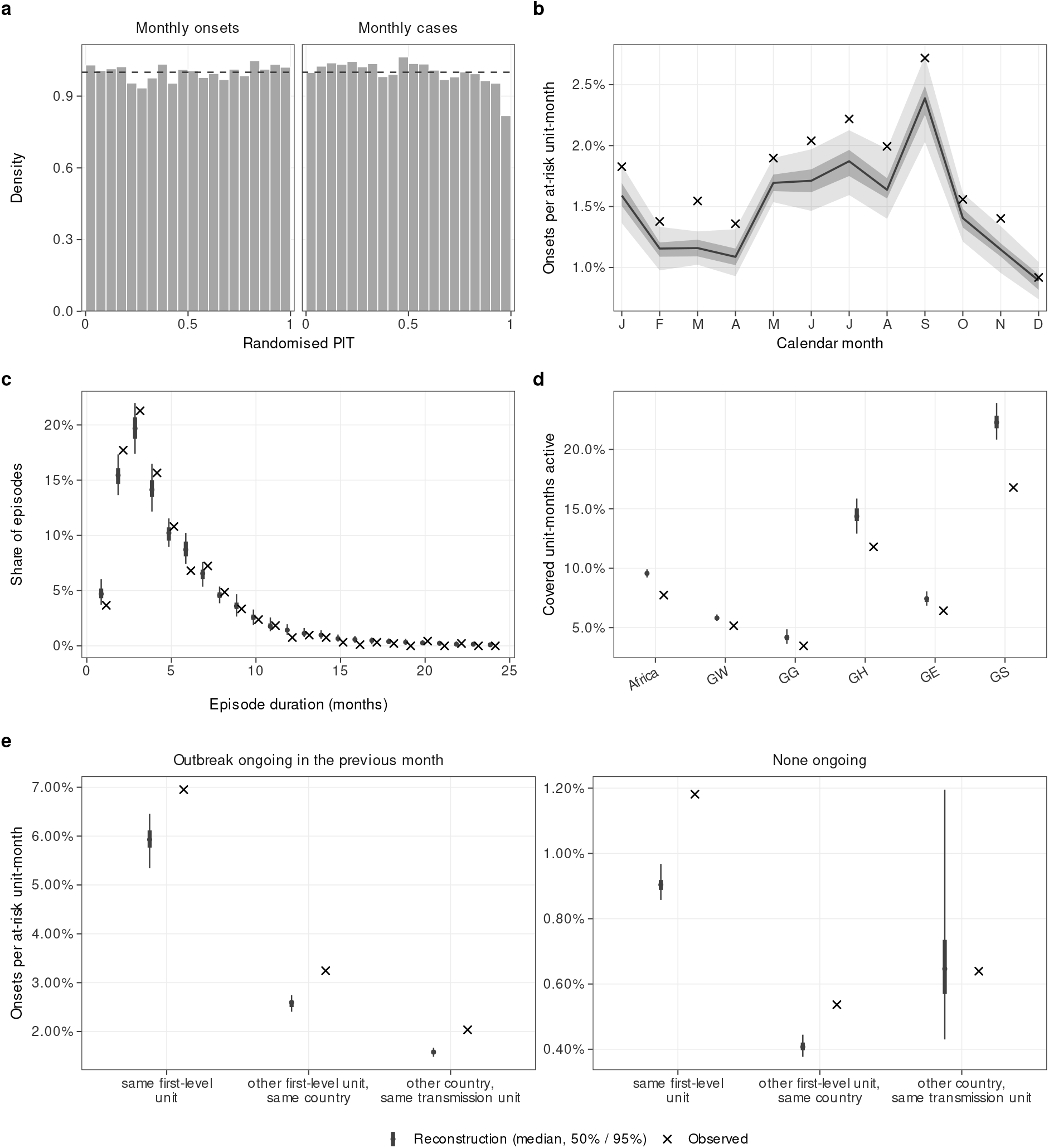
Historical reconstruction against observations. (**a**) Randomised probability integral transform (PIT) histograms of the observed monthly outbreak onsets (unit-months at risk) and monthly suspected cases (surveillance-covered unit-months) under the particle-filter reconstruction; the dashed line is the U(0, 1) reference of a calibrated reconstruction. (**b**) Onsets per at-risk unit-month by calendar month. (**c**) Distribution of episode durations (consecutive active months in a unit). (**d**) Fraction of surveillance-covered unit-months in an active episode, Africa and each ENSO group. (**e**) Onsets per at-risk unit-month among units not active themselves in the previous month, at three scopes (horizontal axis: the same first-level unit, another first-level unit of the same country, another country of the same transmission unit), when an outbreak was ongoing at that scope in the previous month (left) or none was (right). Reconstruction: median (point or line) with 50% and 95% intervals across replicates; observed: crosses. Panels a, b, c and e pool all modelled units in Africa. In-sample: the reconstruction is conditioned on the observations it is compared with (Section S1.6.9).

**Fig. S16.**
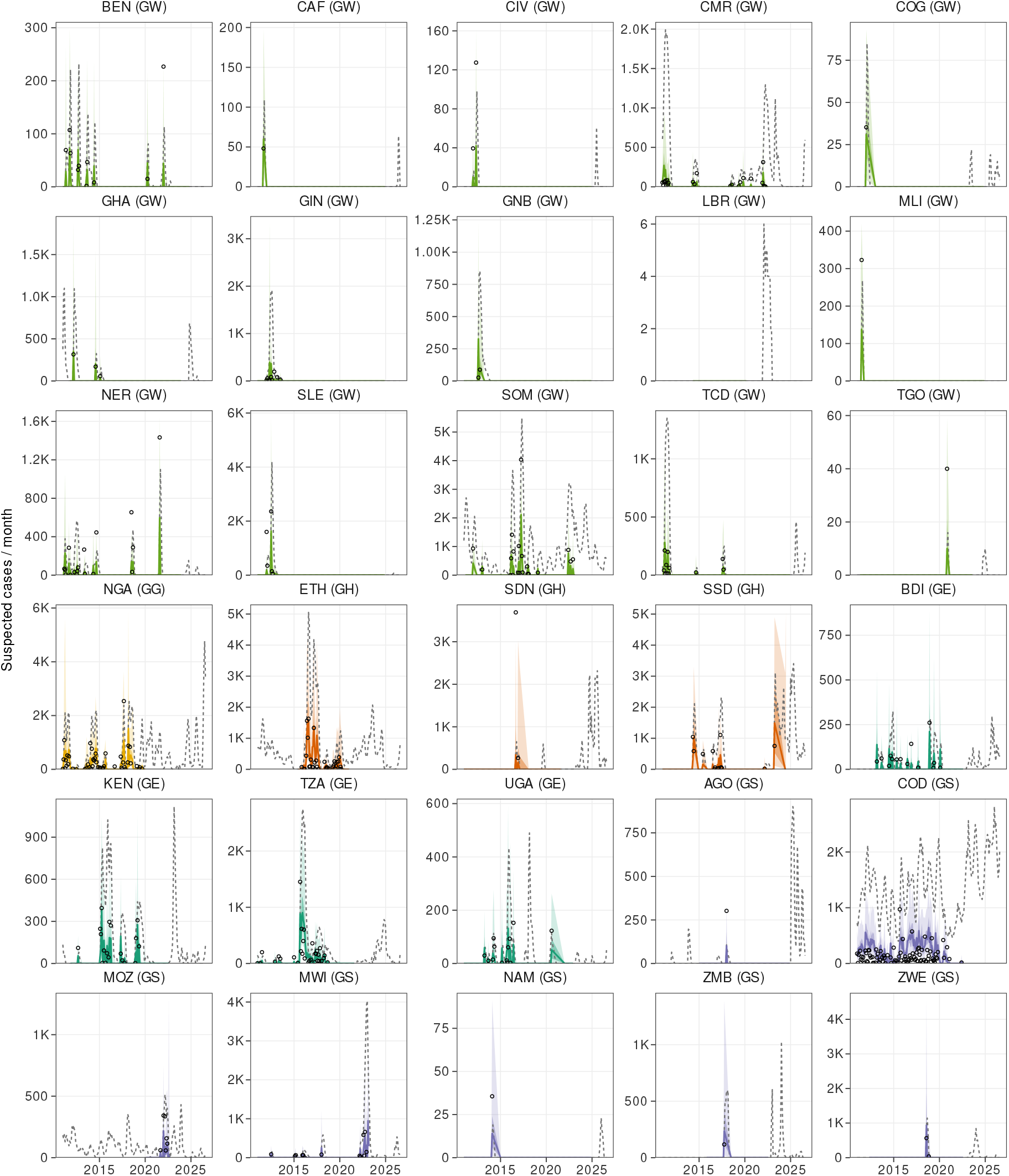
Historical reconstruction by country, suspected cases. Monthly suspected cases per country in the particle-filter reconstruction: median (line) with 50% and 95% bands over the surveillance-covered unit-months, median over all modelled units (grey, dashed), and recorded monthly cases (circles; months with at least one case). Panels are labelled with the country’s ENSO group and ordered by group; the vertical scale differs between panels.

**Fig. S17.**
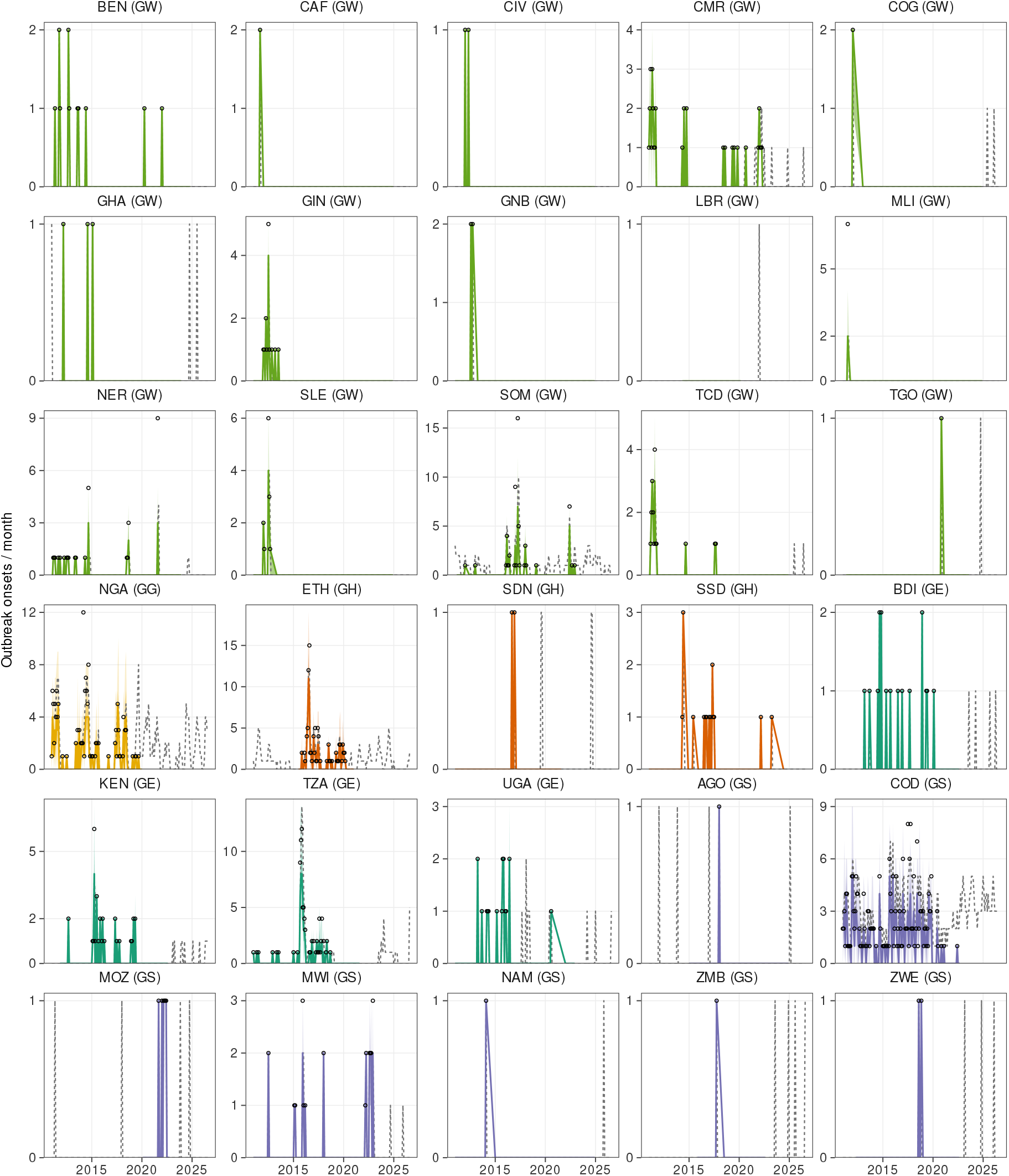
Historical reconstruction by country, outbreak onsets. Monthly outbreak onsets per country in the particle-filter reconstruction: median (line) with 50% and 95% bands over the surveillance-covered unit-months, median over all modelled units (grey, dashed), and recorded onsets (circles; months with at least one onset). Panels are labelled with the country’s ENSO group and ordered by group; the vertical scale differs between panels.

**Fig. S18.**
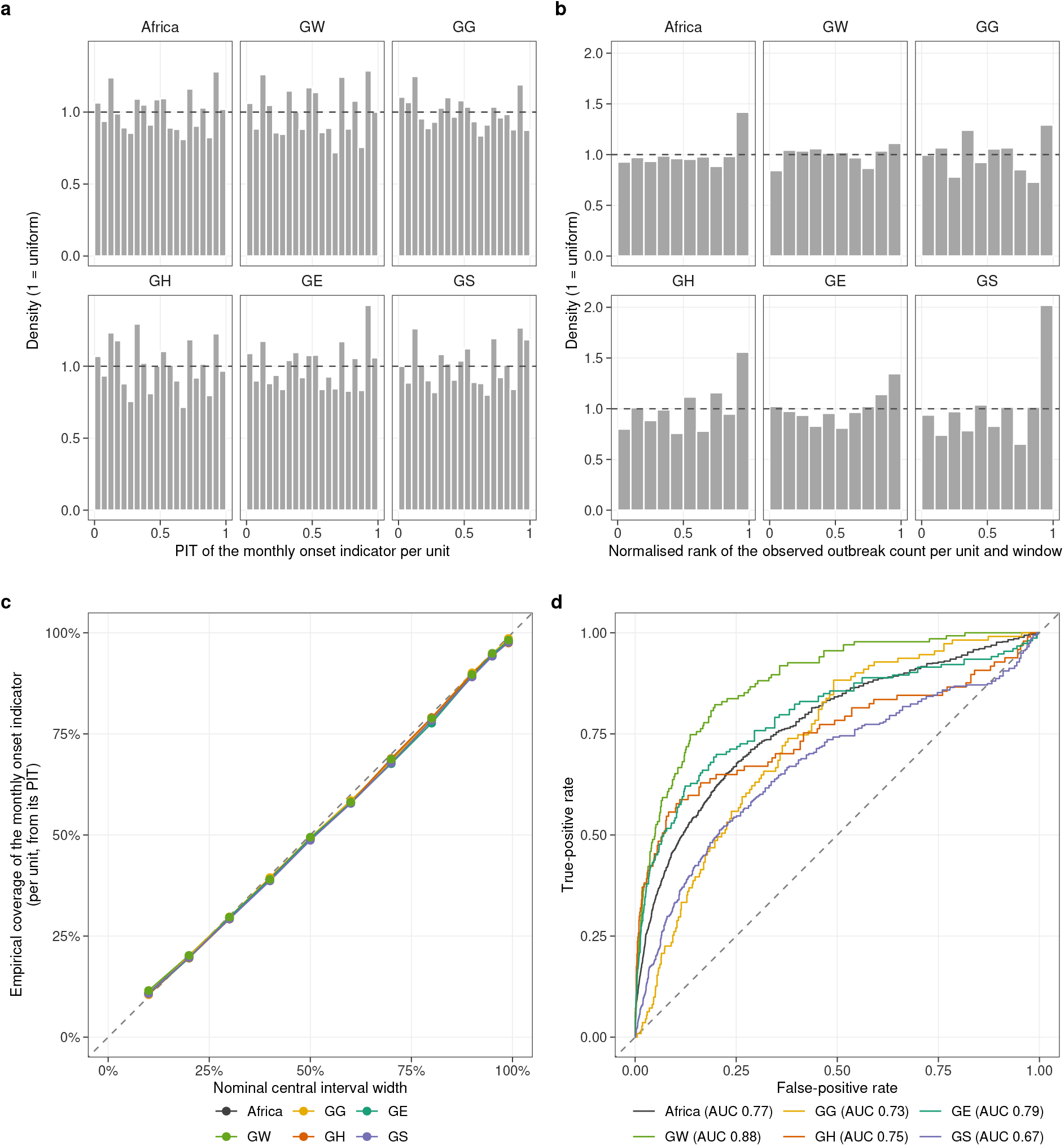
Hindcast calibration. (**a**) Histogram of the probability integral transform of the observed onsets of every unit and hindcast window, pooled by ENSO group and for Africa. (**b**) Randomised rank of the observed window total among the hindcast replicates, all leads. A calibrated forecast gives flat histograms (dashed line: density 1). (**c**) Empirical against nominal coverage of central prediction intervals of the monthly onset indicator per unit, from its probability integral transform (dashed: perfect coverage). (**d**) Receiver operating characteristic curves of the hindcast onset probabilities, with the area under the curve in the legend.

**Fig. S19.**
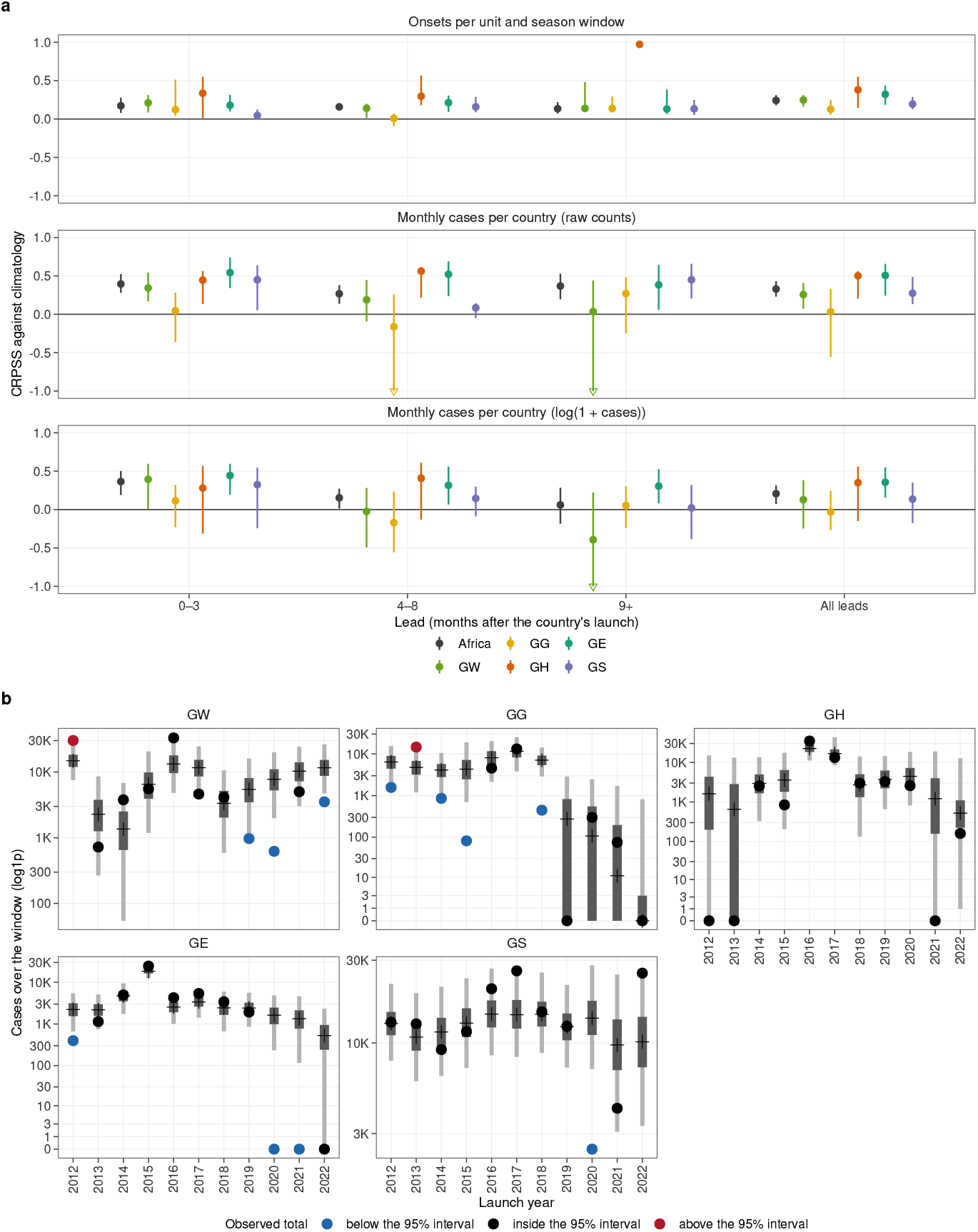
Hindcast skill. (**a**) Continuous ranked probability skill score (CRPSS) of the hindcasts relative to a climatological reference, for Africa and each ENSO group, by lead class (months after each country’s launch: early 0–3, peak season 4–8, late 9 or more, and all leads pooled). Top row: onsets per unit and season window, against a binomial climatology with the unit’s own onset rate outside the scored season. Middle and bottom rows: monthly suspected cases per country on the at-risk units, on raw counts and on log(1 + *y*), against the pooled calendar-month climatology that leaves out the scored season. Points: CRPSS (positive values mean the hindcast beats climatology); bars: 95% intervals from 1,000 bootstrap resamples of whole cholera years. Intervals that extend below *−*1 are cut at *−*1 and marked with an open triangle. A cell whose climatology is exact on every scored month (zero reference score) has no defined skill and is not shown. Section S1.6.10 describes the scores and references. (**b**) Suspected cases over each hindcast window by launch year, for each ENSO group, all leads. Both totals are taken over the units under surveillance in the window (at least one at-risk month), not only those with observed cases, so simulated outbreaks where none was recorded count against the hindcast; observed first-level totals are apportioned to these units by population share. Hindcast median (cross) with 50% (dark bar) and 95% (light bar) prediction intervals, and the observed total (point; black when inside the 95% interval, blue below it, red above it). The observed total falls below the 95% interval in 11 of the 55 group windows and above it in two: the hindcasts tend to over-predict case totals. Vertical axis on a log(1 + *y*) scale.

**Fig. S20.**
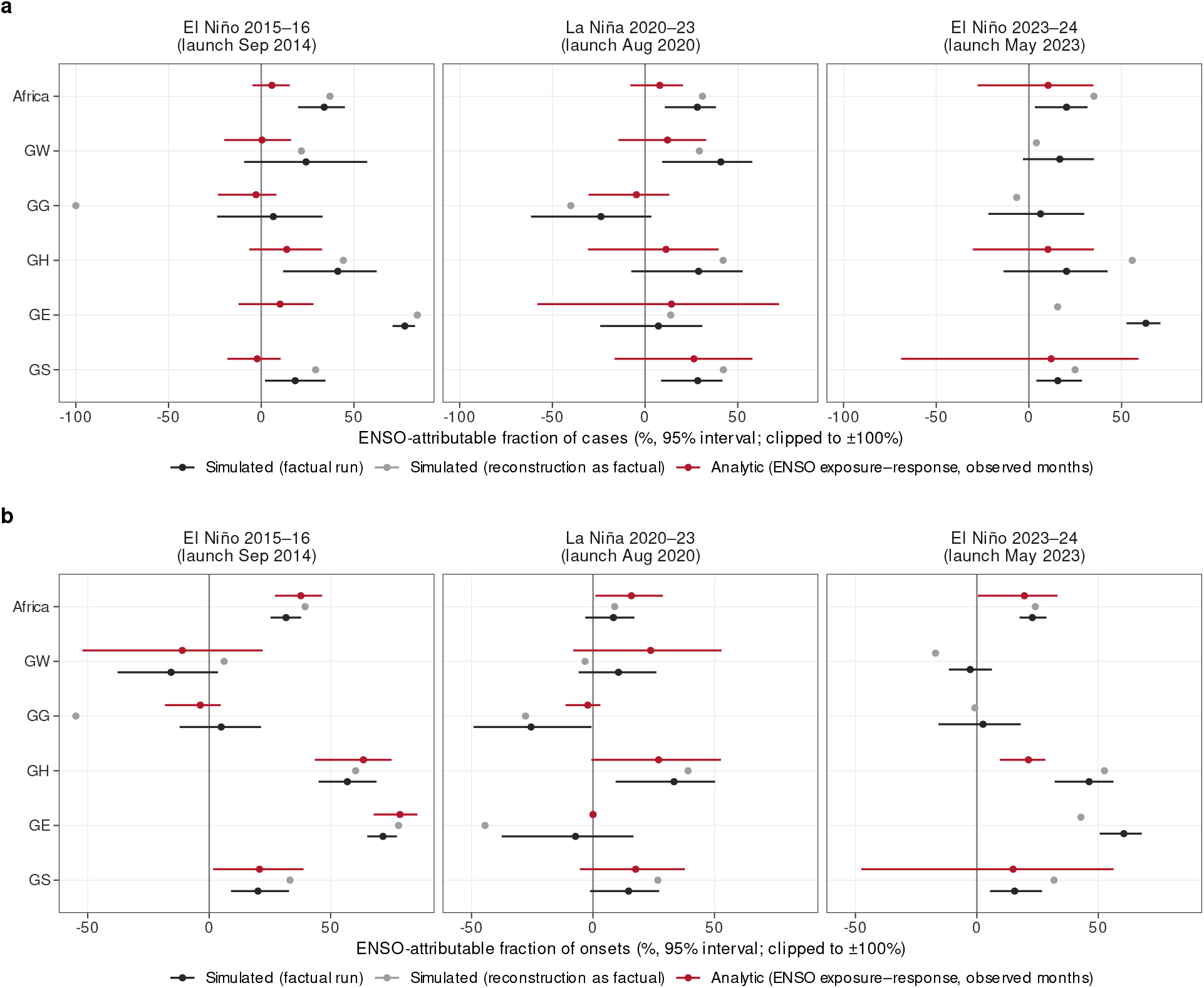
Attribution of past ENSO events. ENSO-attributable fraction of cases (**a**) and outbreak onsets (**b**) per event, for Africa and each ENSO group, against the neutral-ENSO counterfactual (eq. S29), for the free factual simulation, for the historical reconstruction as factual series, and for the analytic calculation on the observed data (eq. S33). Points are point estimates (the ratio of mean factual and counterfactual totals for the simulated readings). Bars: 95% posterior intervals over draws (Section S1.6.14) for the free factual simulation, and over posterior draws and counterfactual trajectories for the analytic reading; the reading with the reconstruction as factual is a point estimate. Values are clipped to *±*100%; the analytic reading is absent where the window has no observed unit-months. Every replicate pair is kept.

**Fig. S21.**
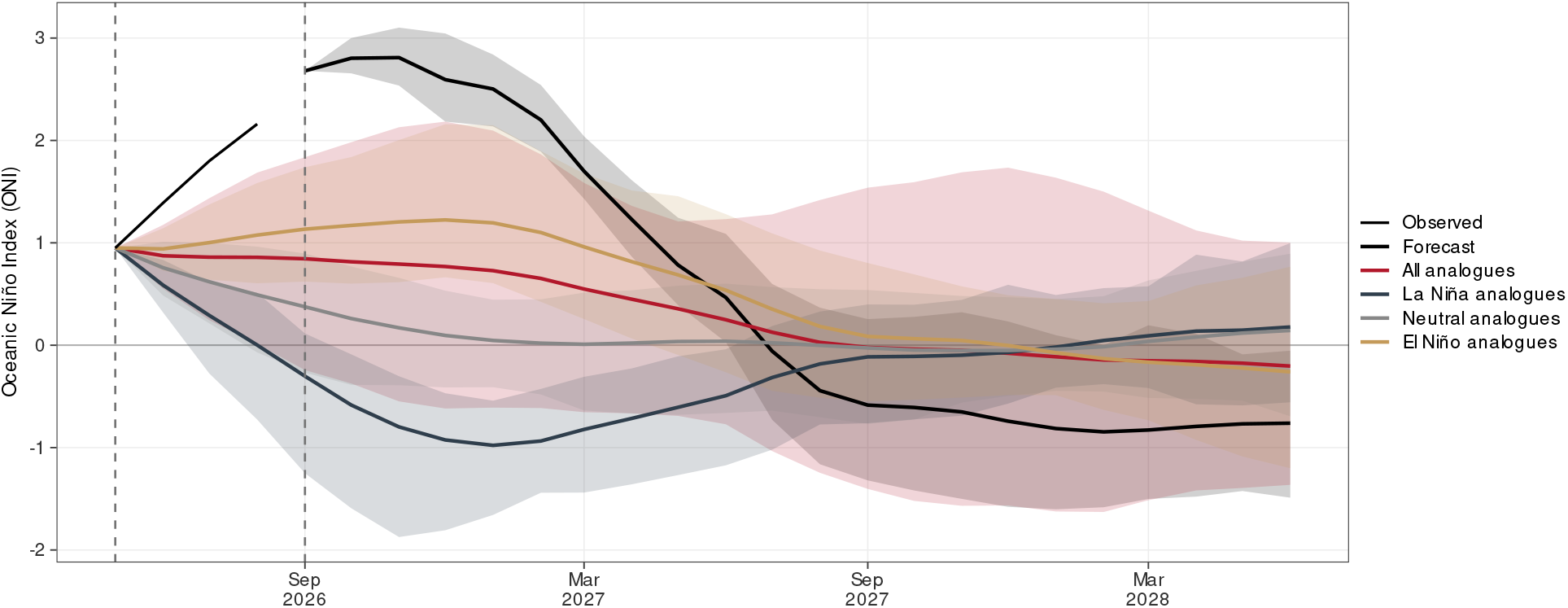
ENSO inputs of the forecast and counterfactual arms. Mean (line) and 90% envelope (5th to 95th percentiles, band) of the ONI trajectories of the forecast arm (CFSv2-based) and of the four pre-industrial analogue sets of the counterfactual arm (all, La Niña, neutral and El Niño analogues), with the observed ONI (black). Dashed vertical lines mark the launches of the two arms.

**Fig. S22.**
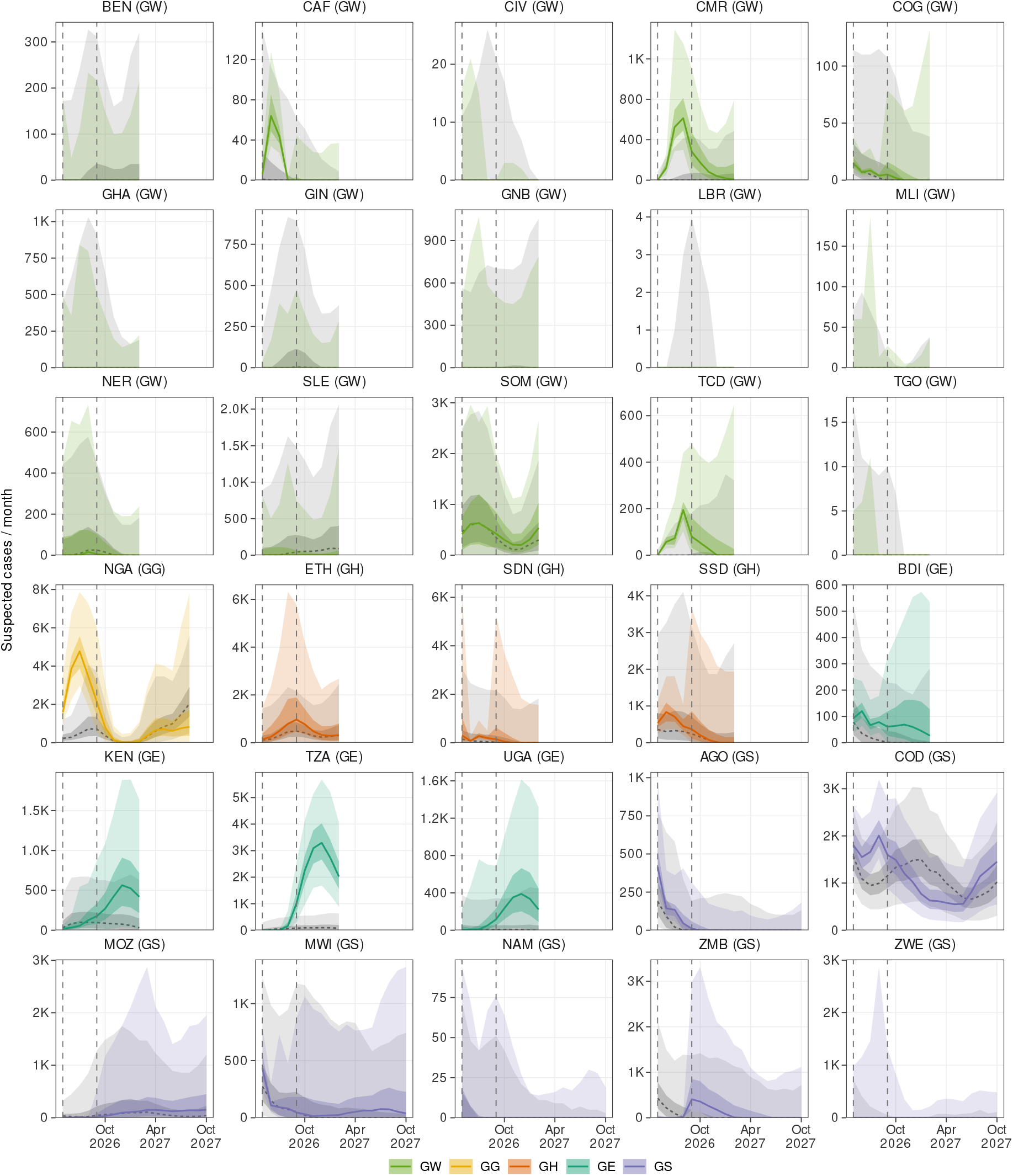
Forecast and neutral-ENSO counterfactual by country, suspected cases. Monthly suspected cases per country under the ENSO-informed forecast (ENSO-group colour) and the neutral-ENSO counterfactual (grey, dashed): median with 50% and 95% bands across paired replicates, each country to the firm horizon of its ENSO group. Dashed vertical lines mark the counterfactual and factual launches. Panels are labelled with the country’s ENSO group; the vertical scale differs between panels.

**Fig. S23.**
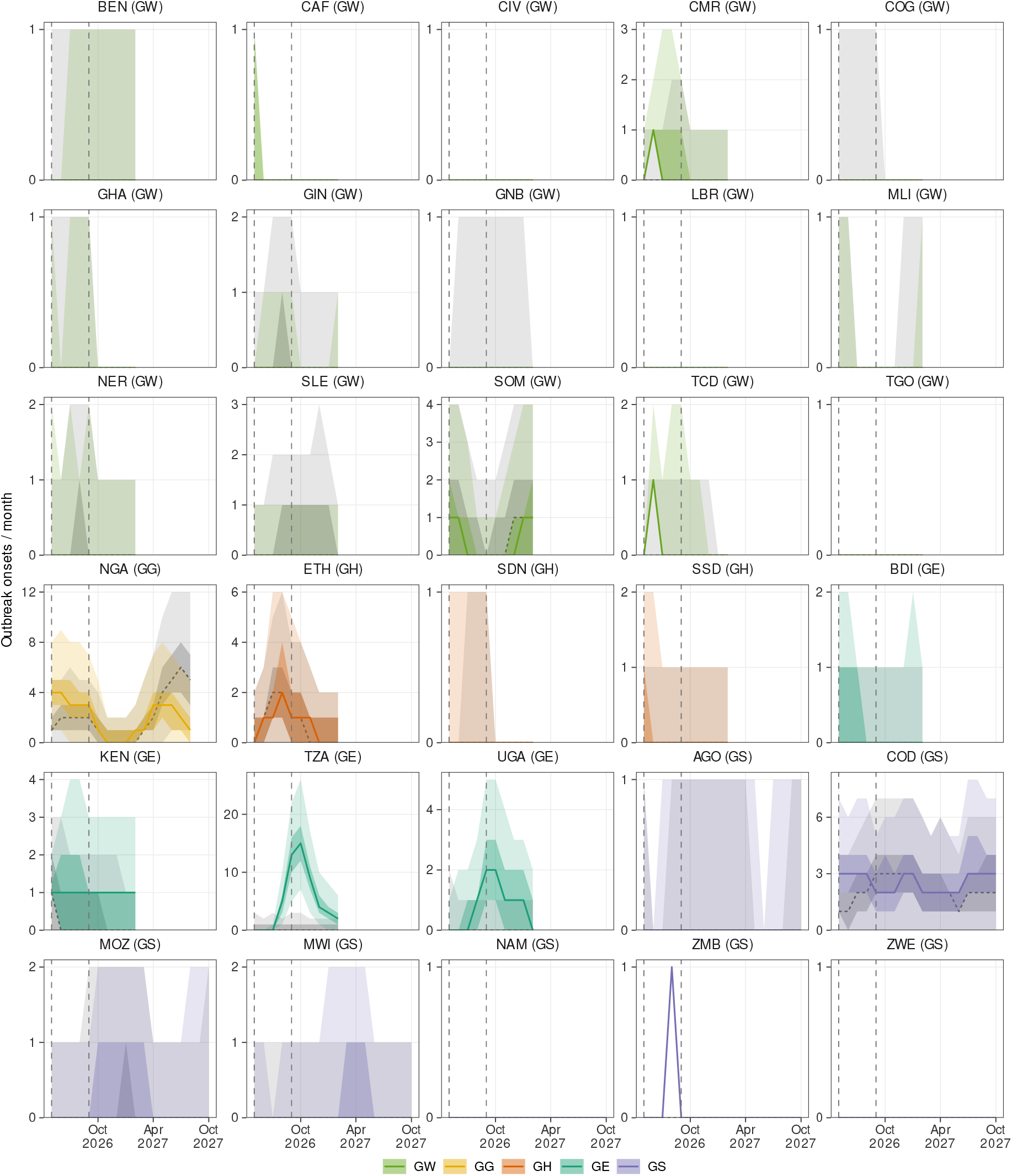
Forecast and neutral-ENSO counterfactual by country, outbreak onsets. Monthly outbreak onsets per country under the ENSO-informed forecast (ENSO-group colour) and the neutral-ENSO counterfactual (grey, dashed): median with 50% and 95% bands across paired replicates, each country to the firm horizon of its ENSO group. Dashed vertical lines mark the counterfactual and factual launches. Panels are labelled with the country’s ENSO group; the vertical scale differs between panels.

**Fig. S24.**
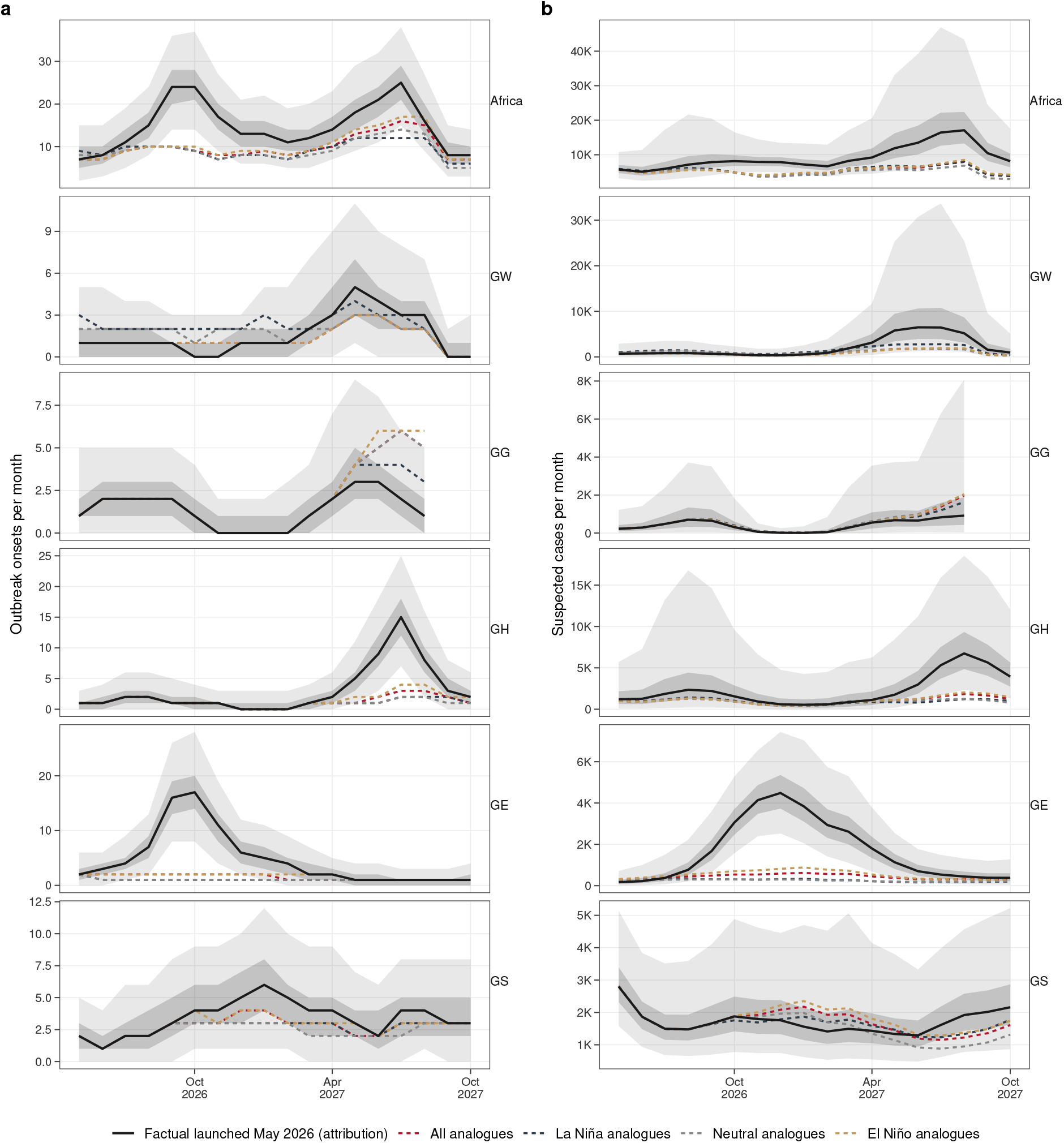
Forecast against counterfactual, by analogue set. Monthly outbreak onsets (**a**) and suspected cases (**b**) in Africa (top row) and in each ENSO group, from the start of the attribution window. Black: the factual arm of the attribution, median (line) with 50% and 95% bands across replicates. Dashed: the median of the counterfactual arm driven by each set of pre-industrial ENSO analogues (all, La Niña, neutral and El Niño analogues); the neutral set is the counterfactual of the main text. Vertical scales differ between rows.

**Fig. S25.**
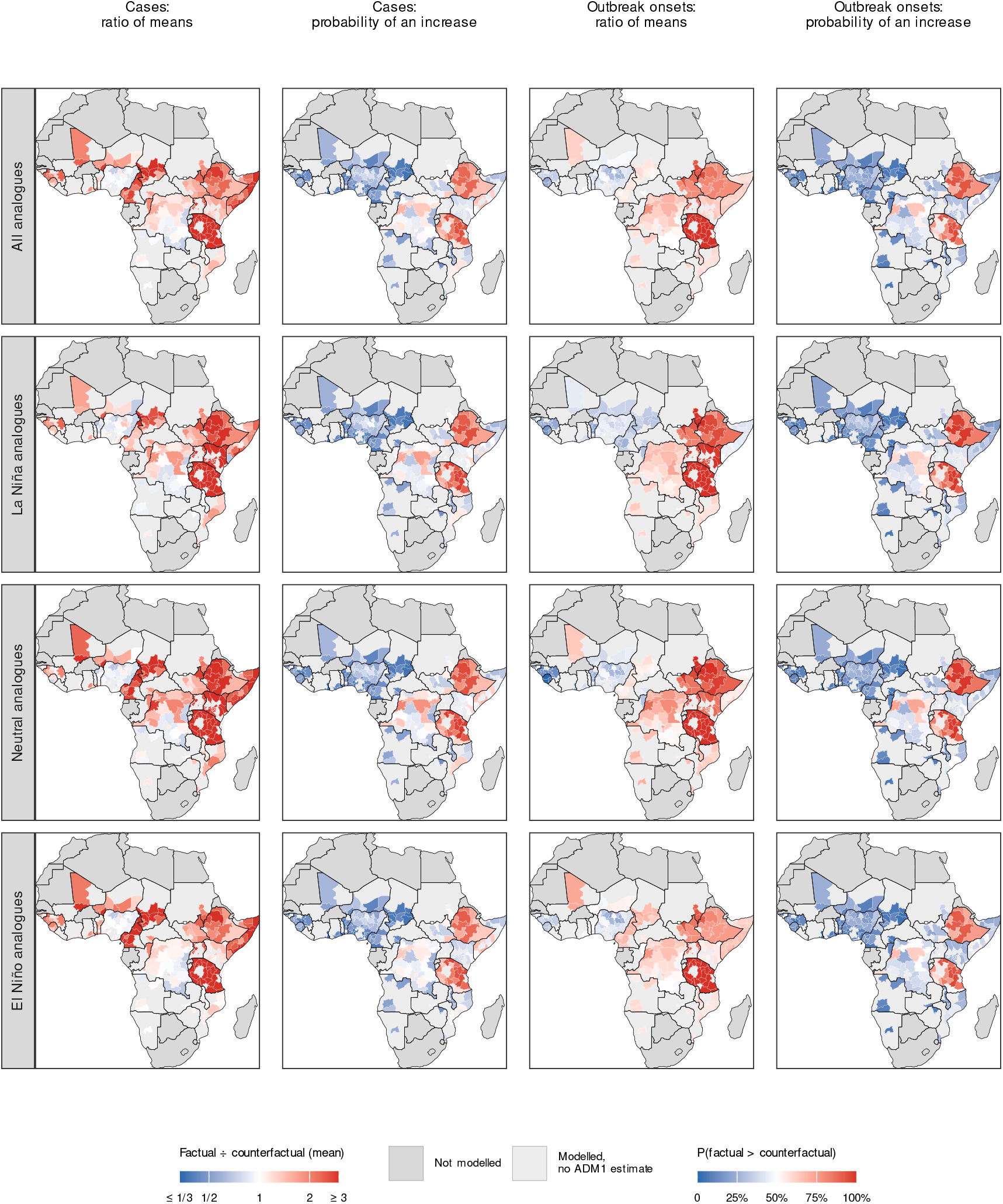
Attributable burden, by analogue set. For suspected cases (left pair of columns) and outbreak onsets (right pair), by first-level unit: ratio of the mean factual to the mean counterfactual total over the attribution window (log scale, capped at 1/3 and 3; a unit whose counterfactual total is zero is drawn at the cap), and probability that the factual total exceeds the counterfactual one across paired replicates. Rows: counterfactuals driven by all, La Niña, neutral and El Niño pre-industrial ENSO analogues (the neutral row is the counterfactual of the main text). Same attribution window, and the same rule for removing replicate pairs, as the main-text attribution map. Units with a zero total in both arms, and units outside the modelled countries, are grey.

**Fig. S26.**
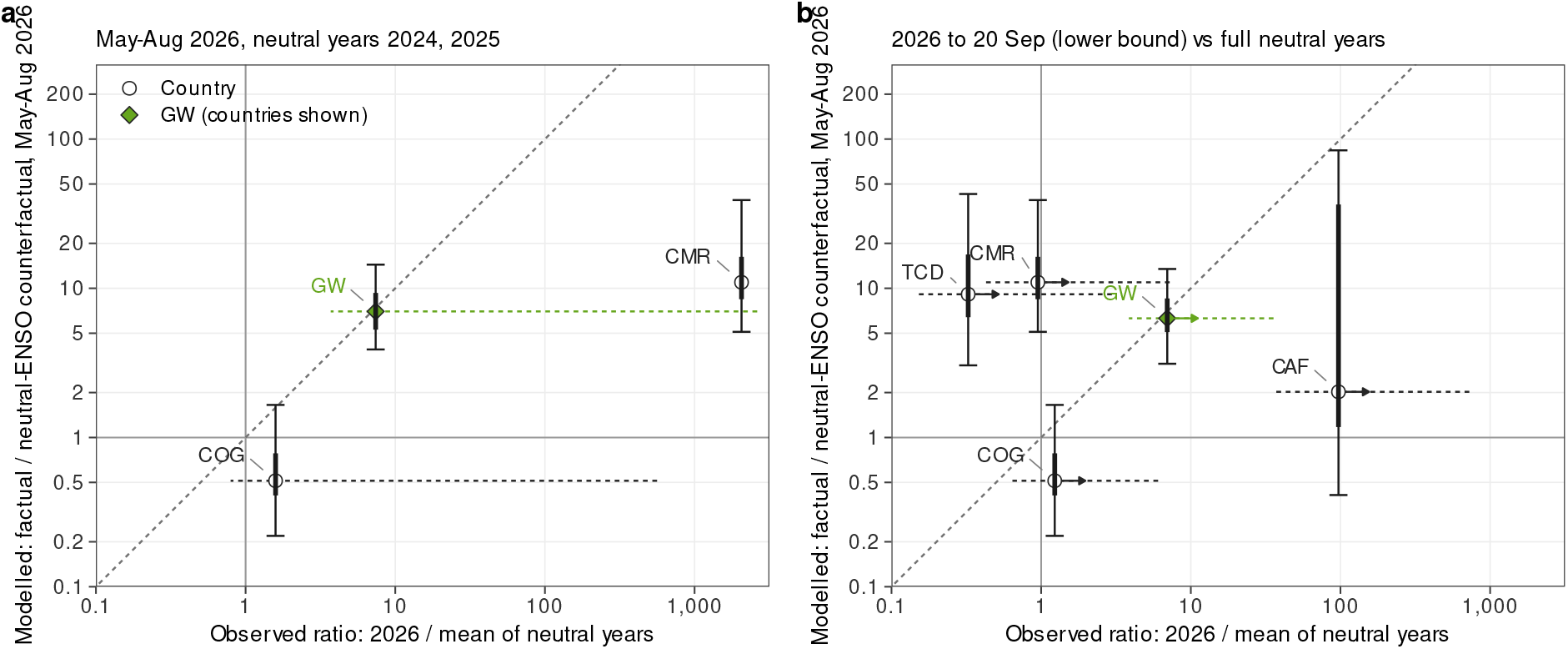
Observed and modelled ENSO ratio of 2026, West group. Horizontal axis: observed ratio of 2026 WHO suspected cases to the mean of the ENSO-neutral years (annual-mean ONI within *±*0.5), population-standardised to 2026, with one pseudo-case on each side; dotted bar, 95% bootstrap interval over neutral years. Vertical axis: modelled ratio of the historical reconstruction to its neutral-ENSO counterfactual over May–August 2026, each counterfactual replicate paired with the reconstruction replicate it was launched from; bars, 50% (thick) and 95% (thin) intervals over reconstruction replicates. Circles: countries; diamond: the West group (GW) over the countries shown. Dashed diagonal: equality. (**a**) Same calendar months, May–August 2026 against the neutral years 2024 and 2025 of the WHO monthly series. (**b**) 2026 to 20 September against full neutral years since 2010, a lower bound (arrows). Countries without a WHO series are not shown; Somalia, whose 2026 series stops in January, is censored. Cameroon’s May–August neutral years are both zero, so its observed ratio in panel a reflects the pseudo-count and has no interval. The reconstruction is filtered on the same WHO series (Section S1.6.8).

## Supplementary Tables

**Table S1.**
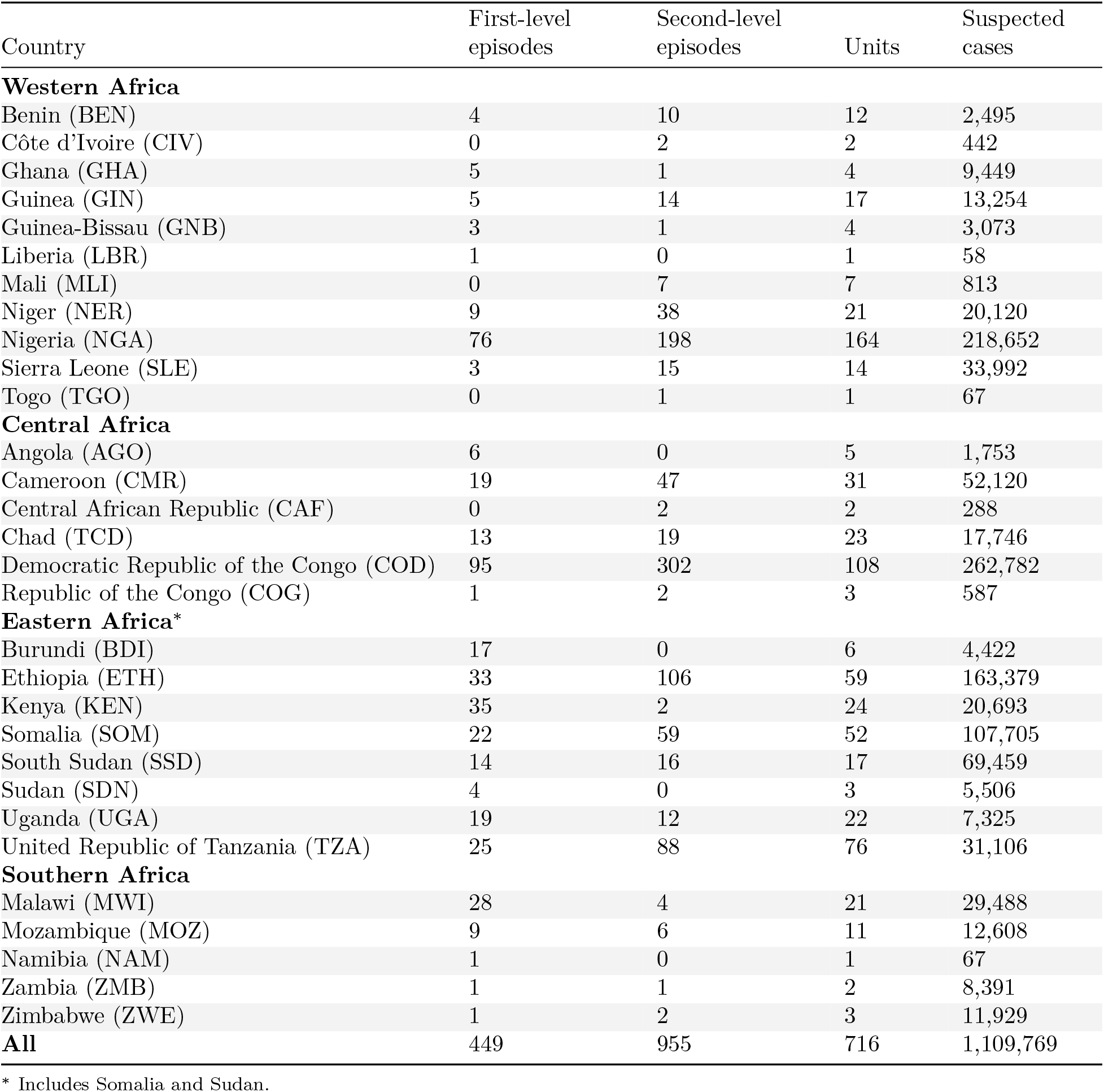
Episodes by country and administrative level. Number of canonical episodes per country at each administrative level, administrative units with an episode, and suspected cases in the episodes; countries grouped by World Health Organization African Region (WHO-AFRO) subregion (bold rows). National-level (ADM0) records are excluded.

**Table S2.** The analysis set by administrative level, Africa and WHO-AFRO subregion. Canonical episodes of the analysis set by the administrative level of their unit, in the layout of Zheng et al. [36], for Africa and each World Health Organization African Region (WHO-AFRO) subregion; national-level (ADM0) records are excluded. Units: administrative units with an episode; cholera years: number of cholera years with an episode. Cases, duration (weeks from the first to the last week of the episode, closing run excluded), population of the unit and attack rate (suspected cases per 1,000 population) are given as median [interquartile range] over episodes. Cholera years start at each location’s seasonal trough (Section S1.1.6), so the January 2010 to November 2024 record spans 16 cholera years.

| Statistic | First level | Second level | All |
|---|---|---|---|
| **Africa** |  |  |  |
| Episodes | 449 | 955 | 1,404 |
| Units | 201 | 515 | 716 |
| Cholera years | 16 | 16 | 16 |
| Suspected cases | 570,313 | 539,456 | 1,109,769 |
| Cases per episode | 405 [149–1,337] | 211 [103–528] | 257 [115–709] |
| Duration (weeks) | 14.9 [7.9–24.9] | 12.9 [7.9–22.9] | 13.9 [7.9–23.9] |
| Population (thousands) | 930 [408–2,716] | 257 [140–472] | 357 [171–780] |
| Attack rate (per 1,000) | 0.46 [0.16–1.41] | 0.96 [0.42–2.18] | 0.78 [0.30–1.95] |
| **Western Africa** |  |  |  |
| Episodes | 106 | 287 | 393 |
| Units | 51 | 196 | 247 |
| Cholera years | 14 | 13 | 14 |
| Suspected cases | 160,419 | 141,995 | 302,414 |
| Cases per episode | 365 [126–1,384] | 160 [90–336] | 182 [95–481] |
| Duration (weeks) | 11.4 [6.9–22.6] | 8.9 [4.9–13.9] | 8.9 [5.9–16.9] |
| Population (thousands) | 2,398 [509–4,667] | 267 [160–450] | 347 [188–675] |
| Attack rate (per 1,000) | 0.32 [0.14–0.78] | 0.70 [0.33–1.53] | 0.61 [0.24–1.34] |
| **Central Africa** |  |  |  |
| Episodes | 134 | 372 | 506 |
| Units | 41 | 131 | 172 |
| Cholera years | 14 | 14 | 14 |
| Suspected cases | 146,081 | 189,194 | 335,275 |
| Cases per episode | 582 [207–1,325] | 247 [102–618] | 305 [118–750] |
| Duration (weeks) | 22.4 [11.9–37.6] | 19.9 [11.6–31.9] | 19.9 [11.9–32.9] |
| Population (thousands) | 789 [338–2,573] | 181 [109–374] | 259 [129–550] |
| Attack rate (per 1,000) | 0.83 [0.19–1.81] | 1.36 [0.67–2.76] | 1.19 [0.56–2.64] |
| **Eastern Africa**<sup>\*</sup> |  |  |  |
| Episodes | 169 | 283 | 452 |
| Units | 84 | 175 | 259 |
| Cholera years | 15 | 16 | 16 |
| Suspected cases | 212,072 | 197,523 | 409,595 |
| Cases per episode | 334 [144–1,333] | 285 [126–625] | 295 [130–758] |
| Duration (weeks) | 12.9 [6.9–21.9] | 12.9 [7.9–20.9] | 12.9 [6.9–21.9] |
| Population (thousands) | 846 [490–1,696] | 338 [189–724] | 476 [242–1,127] |
| Attack rate (per 1,000) | 0.36 [0.16–1.35] | 0.72 [0.29–2.00] | 0.57 [0.22–1.74] |
| **Southern Africa** |  |  |  |
| Episodes | 40 | 13 | 53 |
| Units | 25 | 13 | 38 |
| Cholera years | 11 | 5 | 11 |
| Suspected cases | 51,741 | 10,743 | 62,484 |
| Cases per episode | 367 [130–1,398] | 166 [94–523] | 317 [122–1,291] |
| Duration (weeks) | 11.4 [7.9–18.1] | 9.9 [6.9–12.9] | 10.9 [7.9–16.9] |
| Population (thousands) | 742 [440–2,434] | 187 [178–243] | 632 [318–1,438] |
| Attack rate (per 1,000) | 0.45 [0.18–1.28] | 1.02 [0.55–2.28] | 0.59 [0.22–1.50] |

<sup>\*</sup> Includes Somalia and Sudan.

**Table S3.** Outbreaks by ENSO state of the cholera year, Africa and WHO-AFRO subregion. Each canonical episode takes the ENSO state of its cholera year: the December–February Oceanic Niño Index of that year, El Niño at or above +0.5 and La Niña at or below *−*0.5, with no persistence requirement. Outbreaks: number and share of the block’s episodes; outbreaks per year: episodes divided by the number of cholera years in that state over the record, the same for every block; cholera years (Africa): number of cholera years with an episode. Cases, duration (weeks) and attack rate (suspected cases per 1,000 population): median [interquartile range] over episodes; p: Kruskal–Wallis test across the three states, not adjusted for location, season or year, which the models account for; these summaries do not enter the model fits. Blocks: Africa and the World Health Organization African Region (WHO-AFRO) subregions; national-level (ADM0) records are excluded. Cholera years start at each location’s seasonal trough (Section S1.1.6), so the January 2010 to November 2024 record spans 16 cholera years.

| Statistic | La Niña | Neutral | El Niño | All | p |
|---|---|---|---|---|---|
| **Africa** |  |  |  |  |  |
| Outbreaks, n (%) | 577 (41.1%) | 333 (23.7%) | 494 (35.2%) | 1,404 (100.0%) | – |
| Outbreaks per year | 96.2 | 83.2 | 82.3 | 87.8 | – |
| Cholera years | 6 | 4 | 6 | 16 | – |
| Suspected cases | 236 [108–749] | 343 [142–881] | 222 [102–574] | 257 [115–709] | <0.001 |
| Duration (weeks) | 12.9 [6.9–23.9] | 14.9 [8.9–24.9] | 12.9 [6.9–22.9] | 13.9 [7.9–23.9] | 0.006 |
| Attack rate (per 1,000) | 0.79 [0.30–1.92] | 1.00 [0.38–2.81] | 0.67 [0.28–1.54] | 0.78 [0.30–1.95] | <0.001 |
| **Western Africa** |  |  |  |  |  |
| Outbreaks, n (%) | 209 (53.2%) | 75 (19.1%) | 109 (27.7%) | 393 (100.0%) | – |
| Outbreaks per year | 34.8 | 18.8 | 18.2 | 24.6 | – |
| Suspected cases | 195 [92–504] | 225 [113–656] | 156 [96–311] | 182 [95–481] | 0.194 |
| Duration (weeks) | 8.9 [5.9–15.9] | 10.9 [8.4–17.9] | 6.9 [4.9–15.9] | 8.9 [5.9–16.9] | 0.013 |
| Attack rate (per 1,000) | 0.64 [0.26–1.40] | 0.69 [0.25–2.06] | 0.57 [0.24–1.00] | 0.61 [0.24–1.34] | 0.223 |
| **Central Africa** |  |  |  |  |  |
| Outbreaks, n (%) | 232 (45.8%) | 97 (19.2%) | 177 (35.0%) | 506 (100.0%) | – |
| Outbreaks per year | 38.7 | 24.2 | 29.5 | 31.6 | – |
| Suspected cases | 298 [111–840] | 304 [127–798] | 328 [111–632] | 305 [118–750] | 0.464 |
| Duration (weeks) | 19.9 [10.9–30.9] | 24.9 [16.9–37.9] | 18.9 [9.9–33.9] | 19.9 [11.9–32.9] | 0.023 |
| Attack rate (per 1,000) | 1.16 [0.53–2.65] | 1.36 [0.72–2.64] | 1.10 [0.54–2.60] | 1.19 [0.56–2.64] | 0.305 |
| **Eastern Africa**<sup>\*</sup> |  |  |  |  |  |
| Outbreaks, n (%) | 103 (22.8%) | 160 (35.4%) | 189 (41.8%) | 452 (100.0%) | – |
| Outbreaks per year | 17.2 | 40.0 | 31.5 | 28.2 | – |
| Suspected cases | 296 [117–615] | 424 [166–1,248] | 231 [106–582] | 295 [130–758] | <0.001 |
| Duration (weeks) | 10.9 [6.9–22.9] | 13.9 [7.9–21.9] | 11.9 [6.9–19.9] | 12.9 [6.9–21.9] | 0.309 |
| Attack rate (per 1,000) | 0.50 [0.19–1.61] | 0.88 [0.31–3.32] | 0.47 [0.20–1.21] | 0.57 [0.22–1.74] | <0.001 |
| **Southern Africa** |  |  |  |  |  |
| Outbreaks, n (%) | 33 (62.3%) | 1 (1.9%) | 19 (35.8%) | 53 (100.0%) | – |
| Outbreaks per year | 5.5 | 0.2 | 3.2 | 3.3 | – |
| Suspected cases | 391 [132–1,583] | 64 [64–64] | 190 [88–571] | 317 [122–1,291] | 0.108 |
| Duration (weeks) | 13.9 [7.9–18.9] | 10.9 [10.9–10.9] | 8.9 [7.9–11.9] | 10.9 [7.9–16.9] | 0.494 |
| Attack rate (per 1,000) | 0.66 [0.22–2.28] | 0.11 [0.11–0.11] | 0.59 [0.24–0.95] | 0.59 [0.22–1.50] | 0.280 |

<sup>\*</sup> Includes Somalia and Sudan.

**Table S4.** Episodes with a confirmed case against all episodes. Confirmed case counts are reported for 138 of the 1,404 episodes (9.8%) in 13 of the 30 countries (ETH 73, SSD 19, KEN 16, TZA 8, MWI 7, UGA 4, COG 3, GIN 2, SOM 2, AGO 1, GHA 1, NAM 1, TCD 1). Ratio against ENSO-neutral of the onset rate, the median duration and the median monthly suspected cases of El Niño and La Niña onsets (ONI of the onset month beyond *±*0.5), among all episodes (All) and those with a confirmed case (*≥* 1 conf.), onsets February 2010 to February 2020, for Africa and each ENSO group; 95% intervals (Wald for onset rates, location-period bootstrap otherwise); –: fewer than 2 neutral or 5 phase onsets. Bold: the two estimates fall on opposite sides of 1 and at least one interval excludes 1 (1 of 18 computed confirmed contrasts). Last column: within-country ratio, confirmed / other episodes, of the geometric mean of suspected cases (El Niño row) and of duration (La Niña row); over all reporting countries, it is 1.16 [0.77, 1.67] for the attack rate and 1.16 [0.79, 1.70] for the population.

| Set | Phase | Episodes (phase / neutral) | Occurrence: onset rate ratio | Duration: median ratio | Monthly cases: median ratio | Confirmed / other: cases; duration |
|---|---|---|---|---|---|---|
| **Africa** |  |  |  |  |  |  |
| All | El Niño | 306 / 635 | 0.77 [0.57, 1.04] | 0.87 [0.73, 1.00] | 0.83 [0.68, 0.96] |  |
| All | La Niña | 303 / 635 | **1.13 [0.83, 1.53]** | 0.87 [0.73, 1.00] | 1.03 [0.88, 1.21] |  |
| ≥ 1 conf. | El Niño | 46 / 81 | 0.91 [0.50, 1.64] | 1.00 [0.68, 1.34] | 0.78 [0.53, 1.28] | 1.35 [1.02, 1.71] |
| ≥ 1 conf. | La Niña | 11 / 81 | **0.32 [0.11, 0.91]** | 0.82 [0.23, 1.30] | 0.87 [0.39, 2.04] | 1.35 [1.12, 1.60] |
| **GW** |  |  |  |  |  |  |
| All | El Niño | 27 / 137 | 0.31 [0.14, 0.72] | 0.66 [0.46, 0.90] | 1.14 [0.71, 1.65] |  |
| All | La Niña | 97 / 137 | 1.67 [0.99, 2.81] | 1.34 [0.88, 1.91] | 0.88 [0.68, 1.14] |  |
| ≥ 1 conf. | El Niño | 0 / 3 | – | – | – | 0.85 [0.40, 1.76] |
| ≥ 1 conf. | La Niña | 6 / 3 | 4.72 [0.96, 23.21] | 1.20 [0.22, 4.84] | 0.22 [0.03, 1.09] | 0.56 [0.26, 1.16] |
| **GG** |  |  |  |  |  |  |
| All | El Niño | 21 / 122 | 0.27 [0.10, 0.75] | 0.70 [0.55, 1.15] | 0.92 [0.54, 1.21] |  |
| All | La Niña | 103 / 122 | 1.99 [1.13, 3.52] | 0.80 [0.70, 1.00] | 1.09 [0.75, 1.47] |  |
| ≥ 1 conf. | El Niño | 0 / 0 | – | – | – | – |
| ≥ 1 conf. | La Niña | 0 / 0 | – | – | – | – |
| **GH** |  |  |  |  |  |  |
| All | El Niño | 38 / 111 | 0.55 [0.23, 1.27] | 0.76 [0.43, 1.00] | 1.14 [0.62, 2.39] |  |
| All | La Niña | 4 / 111 | 0.09 [0.01, 0.83] | 1.00 [0.56, 2.28] | 2.43 [0.42, 7.65] |  |
| ≥ 1 conf. | El Niño | 20 / 69 | 0.46 [0.19, 1.14] | 0.88 [0.47, 1.68] | 0.86 [0.46, 3.22] | 1.38 [0.96, 1.96] |
| ≥ 1 conf. | La Niña | 3 / 69 | – | – | – | 1.40 [1.14, 1.74] |
| **GE** |  |  |  |  |  |  |
| All | El Niño | 108 / 58 | 2.97 [1.70, 5.17] | 1.30 [0.95, 1.76] | 0.92 [0.69, 1.40] |  |
| All | La Niña | 21 / 58 | 0.85 [0.36, 2.04] | 0.80 [0.39, 1.38] | 1.47 [0.91, 2.34] |  |
| ≥ 1 conf. | El Niño | 20 / 7 | 4.56 [1.51, 13.73] | 1.03 [0.48, 3.25] | 3.61 [0.40, 7.85] | 1.57 [0.89, 2.55] |
| ≥ 1 conf. | La Niña | 1 / 7 | – | – | – | 1.58 [1.09, 2.09] |
| **GS** |  |  |  |  |  |  |
| All | El Niño | 112 / 207 | 0.86 [0.62, 1.21] | 0.77 [0.57, 0.95] | 0.92 [0.68, 1.28] |  |
| All | La Niña | 78 / 207 | 0.89 [0.61, 1.30] | 1.21 [0.82, 1.42] | 1.05 [0.69, 1.35] |  |
| ≥ 1 conf. | El Niño | 6 / 2 | 4.78 [0.79, 28.89] | 0.55 [0.28, 1.83] | 2.14 [0.91, 2.86] | 1.02 [0.60, 1.69] |
| ≥ 1 conf. | La Niña | 1 / 2 | – | – | – | 1.44 [0.74, 2.58] |

**Table S5.** Prior distributions. *N*^+^ denotes a half-normal distribution.

| Parameter | Prior | Analyses |
|---|---|---|
| Intercept α | $\mathcal{N}(0,1)$ | all |
| Fixed effects, including ENSO basis coefficients | $\mathcal{N}(0,0.5)$ | all |
| Episode-age band effects ψ<sub>2</sub>, …, ψ<sub>6</sub> | $\mathcal{N}(0,1.5)$ | duration |
| Random-effect standard deviations, country level | $\mathcal{N}^{+}(0,0.35)$; $\mathcal{N}^{+}(0,0.05)$ for the single-country group GG | all |
| Random-effect standard deviations, administrative-unit levels | $\mathcal{N}^{+}(0,0.35)$ | all |
| Seasonal spline smoothing standard deviation | $\mathcal{N}^{+}(0,0.35)$ | occurrence |
| Log-dispersion intercept | $\mathcal{N}(1,1)$ | monthly |
| Log-dispersion random-effect standard deviation | $\mathcal{N}^{+}(0,0.5)$; country level $\mathcal{N}^{+}(0,0.05)$ for GG | monthly |

**Table S6.** Transmission units. Pre-specified country clusters within which the neighbour-pressure kernel (eq. (S5)) couples first-level units, and the units of simulation. Countries without outbreak data are in parentheses. Clusters as defined in [48].

| Transmission unit | Countries | Modelled countries |
|---|---|---|
| Central–East | AGO, BDI, COD, ETH, KEN, MOZ, MWI, NAM, SDN, SOM, SSD, TZA, UGA, ZMB, ZWE, (DJI, RWA) | 15 |
| West | BEN, CAF, CIV, CMR, COG, GHA, GIN, GNB, LBR, MLI, NER, NGA, SLE, TCD, TGO, (BFA, GAB, GNQ, MRT, SEN, STP) | 15 |

**Table S7.**
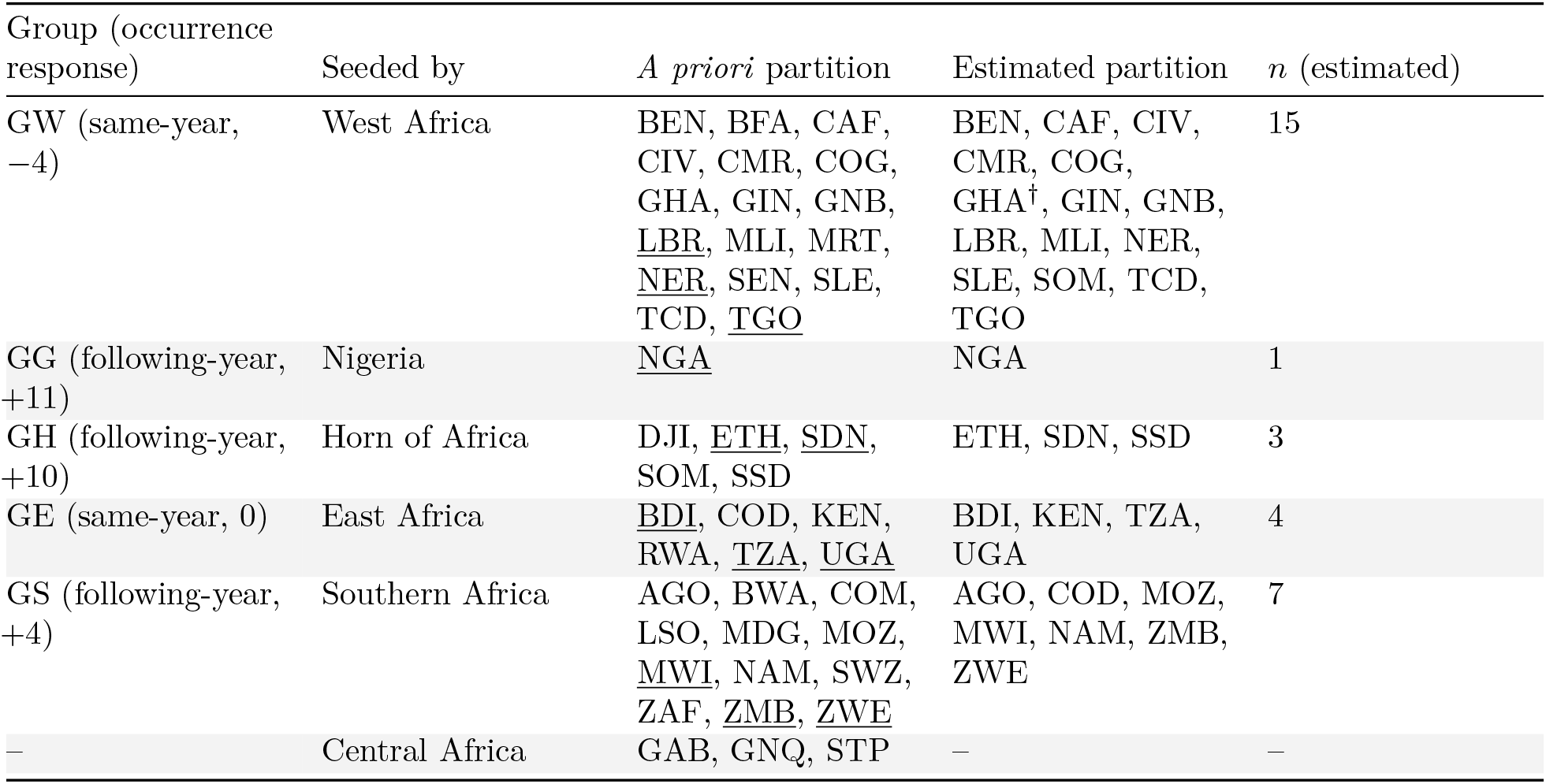
ENSO groups. Countries are given by ISO 3166-1 alpha-3 code (full names in fig. S4). The *a priori* partition is the prior of the latent-class model (eq. S12); the estimated partition (fit with a four-df natural spline and *K* = 5, three agreeing chains) is the one used in all grouped analyses. Groups are named after the part of the continent in which most of their countries lie (GW West, GG Guinean Gulf, which holds Nigeria alone, GH Horn, GE East, GS South), in the order of all figures and tables, with the lag class and selected lag of their occurrence model (Section S1.2.1); “Seeded by” names the *a priori* group whose seed countries fix the class. Seed countries, fixed to their *a priori* group, are underlined. Countries with *p*_max_ *<* 0.6 are marked **^†^** (fig. S6b). Countries without outbreak data appear only in the *a priori* partition.

**Table S8.**
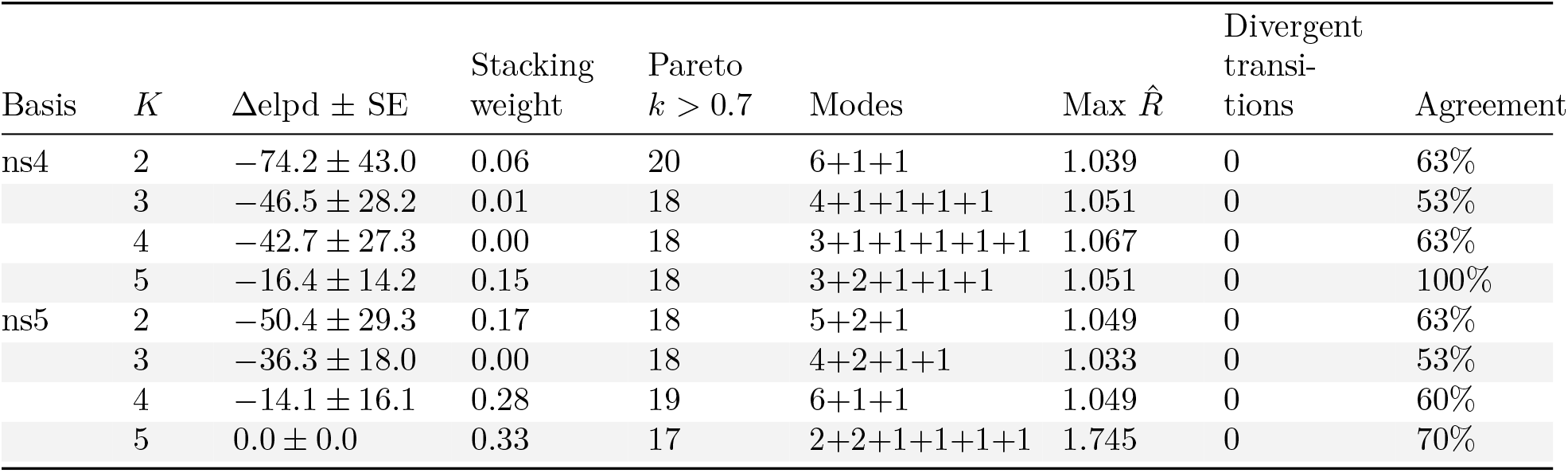
Grouping-model comparison. Country-block PSIS-LOO of the latent-class ENSO lag-kernel occurrence model by ENSO exposure basis and number of classes *K*: difference in expected log predictive density to the best fit with its standard error, stacking weight, number of countries with Pareto *k >* 0.7, divergent transitions and the share of countries whose most probable class agrees with the final partition, on the three agreeing chains of each fit; sizes of the chain modes; and maximum *R̂*, over the two chains of each fit that agree best within its dominant mode, as the posterior of the latent-class model is multimodal (chains 2 and 3 of the final fit). One block per country makes Pareto *k* high by construction, so the ranking is descriptive.

| Basis | *K* | Δelpd ± SE | Stacking weight | Pareto *k* > 0.7 | Modes | Max *R̂* | Divergent transi-tions | Agreement |
|---|---|---|---|---|---|---|---|---|
| ns4 | 2 | −74.2 ± 43.0 | 0.06 | 20 | 6+1+1 | 1.039 | 0 | 63% |
|  | 3 | −46.5 ± 28.2 | 0.01 | 18 | 4+1+1+1+1 | 1.051 | 0 | 53% |
|  | 4 | −42.7 ± 27.3 | 0.00 | 18 | 3+1+1+1+1+1 | 1.067 | 0 | 63% |
|  | 5 | −16.4 ± 14.2 | 0.15 | 18 | 3+2+1+1+1 | 1.051 | 0 | 100% |
| ns5 | 2 | −50.4 ± 29.3 | 0.17 | 18 | 5+2+1 | 1.049 | 0 | 63% |
|  | 3 | −36.3 ± 18.0 | 0.00 | 18 | 4+2+1+1 | 1.033 | 0 | 53% |
|  | 4 | −14.1 ± 16.1 | 0.28 | 19 | 6+1+1 | 1.049 | 0 | 60% |
|  | 5 | 0.0 ± 0.0 | 0.33 | 17 | 2+2+1+1+1+1 | 1.745 | 0 | 70% |

**Table S9.** Selected ENSO lags. Lag in months of the ONI, by analysis and ENSO group, with its class (same-year or following-year; Section S1.2.1); a negative lag reads the index after the outcome month. Groups are labelled as in fig. S6. The firm forecast horizon is the last month whose ONI is a full three-month CFSv2 mean (June 2027, the month before the last CFSv2 target month, July 2027) plus the smallest lag of the group across the three analyses (Section S1.6.12).

| Analysis | GW | GG | GH | GE | GS |
|---|---|---|---|---|---|
| Occurrence | Same-year (−4) | Following-year (+11) | Following-year (+10) | Same-year (0) | Following-year (+4) |
| Duration | Same-year (0) | Following-year (+2) | Same-year (−4) | Following-year (+1) | Following-year (+7) |
| Monthly case counts | Following-year (+8) | Following-year (+10) | Same-year (−2) | Same-year (−4) | Following-year (+4) |
| Firm forecast horizon | February 2027 | August 2027 | February 2027 | February 2027 | October 2027 |

**Table S10.** Past ENSO events attributed. Launch month of both arms (the month before the ONI crossed *±*0.5 into the event), month of the event’s peak (for the 2020–23 La Niña, the last of its three winter peaks, November 2020, November 2021 and October 2022, which closes its window), last month of the comparison window over Africa (each ENSO group’s window ends at the cholera year containing the peak plus the group’s largest positive selected lag; Section S1.6.16), and number of pre-industrial analogues in the counterfactual set. Attributable fractions per event: data S1, files `T4a_event_af_cases and T4b_event af_onsets`.

| Event | Launch | Peak | Window end (Africa) | Analogues |
|---|---|---|---|---|
| El Niño 2015–16 | September 2014 | December 2015 | September 2017 | 86 |
| La Niña 2020–23 | August 2020 | October 2022 | May 2024 | 83 |
| El Niño 2023–24 | May 2023 | December 2023 | September 2025 | 80 |

**Table S11.** Forecast and neutral-ENSO counterfactual cholera incidence, and population in first-level units with more ENSO-attributable cases. May 2026 to October 2027, every ENSO group over the common Africa window (Section S1.6.11). Incidence: suspected cases per 100,000 per year (window total annualised), factual arm (F, May launch: the attribution factual, launched in May 2026) and neutral-ENSO counterfactual (CF); medians and 95% intervals across paired replicates; the denominator is the population of the simulated units, averaged over the window’s months; GG’s runs end in August 2027, so its rate covers 16 months and Africa includes it to then. Over the window, Africa’s F arm totals 173,600 suspected cases (95% interval 119,700–305,000). Attributable fractions of the forecast by window and by analogue set: data S1, files `T5a_forecast_af_windows` and `T5b_forecast_af_sets`. F, September launch: the displayed forecast, launched at the September 2026 handoff of the historical reconstruction (which supplies the months before), each group over its own window from May 2026 to its firm forecast horizon (GW, GH and GE February 2027; GG August 2027; GS October 2027; table S9), a country over its group’s, and Africa to October 2027; annualised as F, medians and 95% intervals across replicates. Childs: population of the first-level units (ADM1) whose F total is at least 1.5 times their CF total [72], applied to each posterior draw of the ADM1’s attributable fraction over the ADM1s whose fraction is identified (Section S1.6.14): posterior mean, 95% credible interval and mean share of the scope’s simulated ADM1 population; IPCC robust: ADM1s where at least 66% of the paired replicates have F above 1.5 times CF and at least 80% have F above CF (IPCC AR6 robustness; Section S1.6.14). Population in millions at 2025; IPCC robust with its share of the scope’s simulated ADM1 population and the number of ADM1s. Groups (bold) as in table S7, with their countries (ISO 3166-1 alpha-3); country rows use the replicates of their group.

| Scope | F, May launch | CF | F, September launch | Childs, population (95% CrI; share) | IPCC robust, population (share; ADM1) |
|---|---|---|---|---|---|
| **Africa** | 22.0 (15.2–38.6) | 11.9 (8.2–17.7) | 24.0 (17.1–40.6) | 271.3 (234.8–302.0; 54%) | 105.3 (21%; 19) |
| **GW** | 25.1 (7.7–81.4) | 12.7 (5.1–25.6) | 11.8 (4.5–24.8) | 39.6 (20.8–53.7; 36%) | 0.0 (0%; 0) |
| BEN | 1.0 (0.0–26.4) | 2.3 (0.0–19.5) | 0.0 (0.0–11.5) | 0.7 (0.0–3.5; 9%) | 0.0 (0%; 0) |
| CAF | 1.4 (0.0–55.6) | 1.4 (0.0–36.2) | 14.4 (8.4–31.6) | 0.6 (0.0–1.1; 54%) | 0.0 (0%; 0) |
| CIV | 0.0 (0.0–1.7) | 0.0 (0.0–1.6) | 0.0 (0.0–1.6) | 0.0 (0.0–0.0; 0%) | 0.0 (0%; 0) |
| CMR | 6.3 (0.0–100.1) | 2.2 (0.0–16.0) | 12.1 (6.0–24.8) | 15.6 (4.8–20.5; 73%) | 0.0 (0%; 0) |
| COG | 15.0 (0.2–455.6) | 14.4 (0.2–162.9) | 20.6 (9.1–158.7) | 0.2 (0.0–0.3; 47%) | 0.0 (0%; 0) |
| GHA | 0.0 (0.0–39.8) | 0.1 (0.0–32.1) | 0.0 (0.0–24.6) | 0.0 (0.0–0.0; 0%) | 0.0 (0%; 0) |
| GIN | 2.0 (0.0–132.7) | 6.4 (0.0–62.0) | 0.0 (0.0–41.0) | 1.7 (0.0–4.8; 23%) | 0.0 (0%; 0) |
| GNB | 0.0 (0.0–672.1) | 1.6 (0.0–477.7) | 0.0 (0.0–544.1) | 0.1 (0.0–0.3; 7%) | 0.0 (0%; 0) |
| LBR | 0.0 (0.0–23.5) | 0.0 (0.0–15.9) | 0.0 (0.0–11.7) | 0.0 (0.0–0.0; 0%) | 0.0 (0%; 0) |
| MLI | 0.0 (0.0–72.0) | 0.0 (0.0–28.1) | 0.0 (0.0–28.5) | 1.5 (0.0–2.0; 78%) | 0.0 (0%; 0) |
| NER | 3.4 (0.0–31.1) | 2.9 (0.0–13.6) | 2.0 (0.0–19.2) | 5.8 (0.0–11.9; 39%) | 0.0 (0%; 0) |
| SLE | 6.1 (0.0–233.0) | 22.0 (0.0–129.9) | 5.2 (0.0–97.5) | 1.0 (0.0–6.0; 14%) | 0.0 (0%; 0) |
| SOM | 120.6 (23.8–430.4) | 49.4 (12.6–117.3) | 40.9 (2.1–123.7) | 9.7 (3.7–14.0; 68%) | 0.0 (0%; 0) |
| TCD | 1.8 (0.0–67.0) | 0.8 (0.0–17.5) | 7.3 (2.8–28.9) | 2.7 (0.5–3.6; 27%) | 0.0 (0%; 0) |
| TGO | 0.0 (0.0–14.6) | 0.0 (0.0–12.5) | 0.0 (0.0–7.2) | 0.0 (0.0–0.0; 0%) | 0.0 (0%; 0) |
| **GG** | 8.0 (2.5–23.8) | 10.0 (4.0–20.5) | 20.4 (13.2–38.4) | 3.7 (0.0–14.4; 5%) | 0.0 (0%; 0) |
| NGA | 8.0 (2.5–23.8) | 10.0 (4.0–20.5) | 20.4 (13.2–38.4) | 3.7 (0.0–14.4; 5%) | 0.0 (0%; 0) |
| **GH** | 26.2 (12.7–62.7) | 11.6 (3.9–30.7) | 13.7 (5.4–40.4) | 111.0 (89.3–120.8; 92%) | 76.4 (63%; 3) |
| ETH | 16.1 (7.6–32.1) | 6.0 (1.4–16.7) | 6.8 (1.0–29.9) | 101.5 (84.7–107.6; 94%) | 76.4 (71%; 3) |
| SDN | 35.0 (0.0–250.3) | 13.5 (0.0–142.0) | 24.0 (4.9–175.3) | 7.4 (0.6–9.7; 76%) | 0.0 (0%; 0) |
| SSD | 233.0 (30.9–1,048.4) | 114.4 (8.2–580.2) | 136.9 (71.8–444.2) | 2.2 (0.8–3.6; 61%) | 0.0 (0%; 0) |
| **GE** | 22.0 (14.9–31.0) | 3.8 (1.7–7.6) | 27.7 (18.4–39.9) | 85.1 (76.7–90.1; 94%) | 27.9 (31%; 15) |
| BDI | 10.2 (0.8–34.7) | 6.6 (0.3–27.4) | 18.0 (6.6–48.1) | 3.1 (0.0–5.6; 55%) | 0.0 (0%; 0) |
| KEN | 11.0 (3.9–22.5) | 3.5 (0.4–10.7) | 10.6 (2.4–27.4) | 31.5 (24.3–33.4; 94%) | 0.0 (0%; 0) |
| TZA | 39.0 (24.9–58.0) | 3.3 (0.7–9.1) | 50.5 (30.6–75.9) | 34.2 (34.2–34.2; 100%) | 27.9 (82%; 15) |
| UGA | 10.2 (2.7–26.3) | 2.6 (0.1–11.6) | 14.4 (3.1–40.3) | 16.3 (11.7–17.4; 94%) | 0.0 (0%; 0) |
| **GS** | 20.6 (13.6–34.2) | 18.4 (11.8–30.9) | 20.8 (14.3–35.6) | 31.8 (9.9–52.3; 31%) | 1.0 (1%; 1) |
| AGO | 3.6 (0.3–15.8) | 3.5 (0.3–18.1) | 6.1 (3.1–15.9) | 1.3 (0.0–5.7; 14%) | 0.0 (0%; 0) |
| COD | 20.5 (13.7–29.7) | 19.5 (12.9–28.0) | 20.6 (15.0–28.3) | 21.8 (4.2–37.8; 33%) | 1.0 (1%; 1) |
| MOZ | 31.6 (3.1–165.8) | 19.2 (0.7–113.3) | 23.8 (1.1–150.8) | 3.5 (0.0–7.4; 48%) | 0.0 (0%; 0) |
| MWI | 12.0 (1.5–48.7) | 8.2 (0.8–43.0) | 11.3 (2.7–43.1) | 5.0 (0.8–9.2; 37%) | 0.0 (0%; 0) |
| NAM | 0.6 (0.0–42.6) | 0.4 (0.0–36.3) | 1.2 (0.0–48.8) | 0.0 (0.0–0.0; 0%) | 0.0 (0%; 0) |
| ZMB | 45.8 (2.0–342.5) | 44.9 (2.0–349.1) | 72.5 (1.1–419.9) | 0.1 (0.0–2.3; 4%) | 0.0 (0%; 0) |
| ZWE | 0.0 (0.0–234.2) | 0.0 (0.0–216.0) | 0.0 (0.0–379.7) | 0.0 (0.0–0.0; 0%) | 0.0 (0%; 0) |

## Data S1

**Data S1. (separate file)** Policy-relevant tabulated outputs, one machine-readable CSV file per table. Every other tabulated output cited in the text is in the GitHub data folder of the repository.

- `T3a_effects_country_occurrence.csv:` ENSO effect on outbreak occurrence by country at each ENSO condition against neutral (posterior median, 95% CrI)
- `T3b_effects_country_duration.csv:` ENSO effect on outbreak duration by country at each ENSO condition against neutral (posterior median, 95% CrI)
- `T3c_effects_country_monthly.csv:` ENSO effect on monthly case counts by country at each ENSO condition against neutral (posterior median, 95% CrI)
- `T4a_event_af_cases.csv:` Attributable fraction of suspected cases per past ENSO event, Africa and by ENSO group (95% posterior interval; every replicate kept)
- `T4b_event_af_onsets.csv:` Attributable fraction of outbreak onsets per past ENSO event, Africa and by ENSO group (95% posterior interval; every replicate kept)
- `T5a_forecast_af_windows.csv:` Attributable fraction of the 2026–27 forecast, Africa and by ENSO group, by attribution window
- `T5b_forecast_af_sets.csv:` Attributable fraction of the 2026–27 forecast, Africa and by ENSO group, by counterfactual analogue set
- `D10_enso_exposure_concordant.csv:` Expected share of the population exposed to an ENSO effect of at least *m* in two or more analyses, Africa and by ENSO group (mean, 95% CrI)
- `D10_enso_exposure_exceedance.csv:` Expected share of the population exposed to an ENSO effect of at least *m* in any analysis, Africa and by ENSO group (mean, 95% CrI)
- `D12_enso_exposure_by_country.csv:` Population with a high-certainty adverse ENSO exposure (*P* (effect beyond 25%) *≥* 95% in at least one analysis), by country
- `D18_attrib_exposure_adm1.csv:` ADM1 units with more ENSO-attributable cases or onsets in the 2026–27 forecast, with their population
- `D18_attrib_exposure_by_country.csv:` Population in ADM1 units with more ENSO-attributable cases or onsets in the 2026–27 forecast at *m* = 50%, by country
- `D18_attrib_exposure_scope.csv:` Population in ADM1 units with more ENSO-attributable cases or onsets in the 2026–27 forecast at *m* = 50%, Africa and by ENSO group

